# Uncovering heterogeneous inter-community disease transmission from neutral allele frequency time series

**DOI:** 10.1101/2024.12.02.24318370

**Authors:** Takashi Okada, Giulio Isacchini, QinQin Yu, Oskar Hallatschek

## Abstract

The COVID-19 pandemic has underscored the critical need for accurate epidemic forecasting to predict pathogen spread and evolution, anticipate healthcare challenges, and evaluate intervention strategies. The reliability of these forecasts hinges on detailed knowledge of disease transmission across different population segments, which may be inferred from within-community transmission rates via proxy data, such as contact surveys and mobility data. However, these approaches are indirect, making it difficult to accurately estimate rare transmissions between socially or geographically distant communities. We show that the steep ramp up of genome sequencing surveillance during the pandemic can be leveraged to *directly* identify transmission patterns between communities. Specifically, our approach uses a hidden Markov model to infer the fraction of infections a community imports from other communities based on how rapidly the allele frequencies in the focal community converge to those in the donor communities. Applying this method to SARS-CoV-2 sequencing data from England and the U.S., we uncover networks of inter-community disease transmission that, while broadly reflecting geographical relationships, also expose epidemiologically significant long-range interactions. We provide evidence that transmission between regions can substantially change between waves of variants of concern, both in magnitude and direction, and analyze how the inferred plasticity and heterogeneity in inter-community transmission impact evolutionary forecasts. Overall, our study highlights population genomic time series data as a crucial record of epidemiological interactions, which can be deciphered using tree-free inference methods.

## Introduction

Despite extensive efforts to model epidemiological dynamics, particularly during the COVID-19 pandemic, accurately predicting epidemic trajectories has proven difficult for populations with heterogeneous transmissibility patterns (1–5). Heterogeneities arise from many factors, including varying population densities, mobility patterns, immunity levels, behaviors, and non-pharmaceutical interventions. While the ensuing transmission rate variations are difficult to predict, ignoring them reduces the applicability of model-based forecasts and may result in misguided interventions (6–9).

In principle, metapopulation models can account for known heterogeneities in the host population by dividing it into suitably many subpopulations, which are distinguished by their epidemiological characteristics. A crucial input to these models is a matrix of parameters that represents the rates at which infections are transmitted between subpopulations. Referred to as infectivity matrix (10–12), it encodes how individuals in different population groups acquire infections from other groups. Metapopulation models can be used to predict the impact of heterogeneities on the disease spreading and evolution. By perturbing the transmission parameters, they also allow for the exploration of group-specific non-pharmaceutical interventions, immunity or behavioral changes.

As the number *n* of subpopulations grows, estimating the *n*^2^ parameters of the infectivity matrix becomes increasingly challenging. Valuable clues about the transmission rates have been gleaned from measuring how often members of different groups come in contact. For example, the real-time tracking of cell phones enables estimating the mobility flux between different regions (13–15) and surveys can be used to infer the mixing between different age groups (16, 17). However, converting these fluxes into transmission rates requires additional assumptions about the infection dynamics during contact. For example, how precisely variations in mask-wearing or immunity levels lower transmission rates is difficult to measure directly (18, 19), thus, requiring estimation methods. Furthermore, differences in local interventions and individual behaviors can weaken the relationship between mobility metrics and transmission rates, thereby reducing their predictive power (20).

To transcend these limitations, a direct data-driven approach to infer heterogeneous disease transmission rates is needed. Ground truths would be valuable even retrospectively, as they could be used to falsify transmission rates obtained from indirect methods and, more broadly, to develop improved evidence-based forecasting of epidemic spread and selective sweeps of new variants. Lastly, knowledge of how infections cross population boundaries can also inform phylogenetic approaches to embed genealogies of past outbreaks into geographical space, which are usually based on the assumption that lineages follow unbiased random walks (21, 22).

We argue that the steep ramp up of the surveillance of virus sequence variants during the COVID-19 pandemic offers unprecedented opportunities to use population genetic tools to obtain a direct view of the underlying metapopulation transmission network. Numerous studies have already demonstrated that the related effort of molecular source attribution (23–25) substantially gains in precision by the abundance of data. For example, embedding the phylogenetic tree of the sampled viruses within the geographical landscape of England has allowed for the reconstruction of detailed spatio-temporal infection processes for different variants of con-cern (26, 27). Nonetheless, inferring actual infection processes necessitates making significant assumptions about the dispersal of lineages (21, 22). Furthermore, phylogenetic methods require constructing genealogical trees, which is computationally challenging for large datasets like the complete set of sequenced SARS-CoV-2 samples.

Our objective is to create a computationally efficient, tree-free approach to infer infection matrices directly from neutral allele frequency time series. We show that, with adequate data, it is possible to map entire networks of disease transmission between communities. By analyzing genomic SARS-CoV-2 data from England, obtained from the COVID-19 Genomics UK Consortium (COG-UK) (28), and data from the USA obtained from GISAID (29), we highlight differences across variants of concern, examine the statistical characteristics of the resulting transmission networks, explore the significance of long- and short-range connections, and assess their impact on the spread of new variants.

## Results

### The Basic Idea

We can illustrate the core of our method by examining how the importation of cases affects allele frequency differences between communities. Imagine two populations, A and B, that are initially epidemiologically isolated due to a complete traffic lockdown, which is later lifted. The population genetic simulations in Fig. 1A show that, during the isolation period, allele frequencies fluctuate independently in each population since there is no interaction between both populations. However, once the lockdown ends, both communities begin exchanging infections, causing their allele frequencies to gradually converge. The higher the transmission rate between the two populations, the faster this frequency alignment occurs. Our method leverages this phenomenon, computationally linking the convergence of allele frequencies to inter-community transmission.

**Fig. 1.**
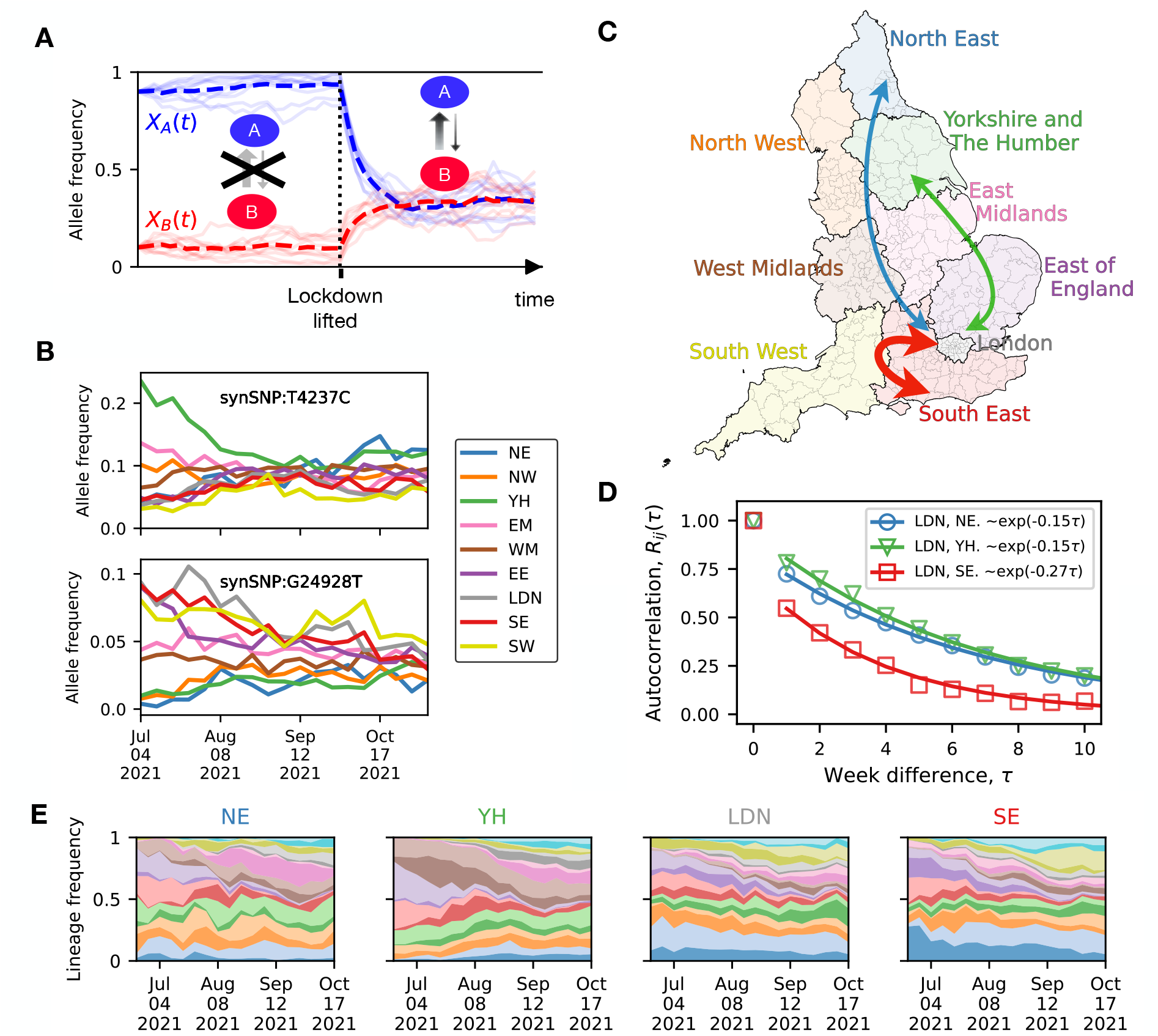
Inter-community transmission promotes allele frequency convergence. (**A**) Simulated dynamics of a neutral variant to illustrate the effect of inter-community disease transmission between two communities, A and B. Ten simulated trajectories are shown with their average as a dashed line. When the two communities are isolated by a traffic lockdown, their allele frequencies fluctuate independently due to genetic drift acting independently in both populations. After the lockdown is lifted (dotted vertical line), the allele frequencies tend to converge due to the exchange of infections between the communities. The rate of convergence reflects the rate of case importations. (**B**) Time series of allele frequencies for two prevalent SARS-CoV-2 alleles (top and bottom graphs), exemplifying allele frequency convergence during the Delta wave in England. Each color corresponds to one of the nine regions of England shown in (**C**). (**D**) Averaged across approximately independent alleles in our data set, the rate of convergence substantially differs for different region pairs. The markers show the autocorrelation function *R*_*ij*_ (*τ*) of the allele frequency differences between regions *i* and *j*. We focus on the three region pairs indicated by arrows in Fig. **C**: London and North East (blue); London and Yorkshire and the Humber (green); and London and South East (red) (see SI Sec. S.1 for the procedure for selecting the alleles over which we have averaged and for the mathematical definition of *R*_*ij*_ (*τ*)). The solid lines represent exponential fits. (**E**) The relative abundance of the top 20 Delta variant lineages over time in North East, Yorkshire and The Humber, London, and South East. Decay rates similar to those in Fig. D are obtained from lineage-frequency data (Fig. S5).

Before delving into our method, it is useful to first verify that the expected alignment of frequency trajectories is indeed observable in SARS-CoV-2 data. Focusing for clarity on just two alleles of the Delta variant, Fig. 1B demonstrates that allele frequencies in different regions in England tend to converge over time. Moreover, Fig. 1D demonstrates that, on average, allele frequency mismatches between nearby regions, such as London and the South East, diminish faster than those between distant regions, like London and the North East, in line with an isolation-by-distance expectation (32) (see SI Fig. S5 for additional comparisons between region pairs). The Muller plots in Fig. 1E illustrate how the sample frequencies of the top 20 Delta variant lineages in the COG-UK phylogenetic tree fluctuated across regions in England during the 2021 Delta wave (SI Sec. S.1).

### Overview of the inference approach

To mathematize the above idea, we resort to the principles by which lineage frequencies evolve under neutrality.

Consider a population composed of *n* sub-populations, distinguished by location (different cities or districts), age, ethnicity, or any other feature that might influence the epidemic characteristics of its members. Under neutrality, we can assume that, up to random fluctuations, the frequency *X*_*i*_(*t*) of a particular lineage in population *i* at time *t* depends linearly on the lineage frequencies {*X*_*j*_(*τ*)}_*j*=1…*n*_ at some earlier time *τ < t*,

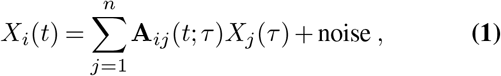

where **A** is a right-stochastic *n* × *n* matrix: its eleI:ments are non-negative and, within each row, sum up to one, 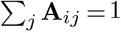. For now, we do not need to know anything about the noise term, except that its expectation vanishes (See Sec. 1.2 for a derivation of Eq. 1).

The coefficient **A**_*ij*_ represents the proportion of infections that population *i* imports from population *j*, thus capturing cross-infection rates between regions. Therefore, we refer to **A** as the *importation-rate matrix*. Yet, an equally important dual interpretation of the matrix **A** arises when one tries to model *backward* processes, where one starts from a given sampled genome and follows its lineage of ancestors backward in time. For this process, **A**_*ij*_(*t*; *τ*) describes the probability that the lineage jumps from population *i* to population *j* as time is run backward from *t* to *τ*. This backward-in-time interpretation is needed, for example, when one tries to embed phylogenetic trees within a metapopulation, because **A**_*ij*_(*t*; *τ*) provides the probabilistic weight for assigning the branch of a phylogenetic tree to a transition from population *i* to *j*. We will frequently return to the interpretation of the rows of **A** as backward transition probabilities, as it helps develop intuition and hypotheses for the structure of **A**. For example, the constraint Σ_*j*_**A**_*ij*_ = 1 is obviously required for the interpretation of the rows of **A** as probability distributions.

The most straightforward way to estimate importation rates, **A**_*ij*_, using the model described above, is to minimize the least squares difference between predicted and observed lineage frequencies over all right stochastic matrices. This method is most effective when the true lineage frequencies are known; however, in practice, these frequencies are derived from a random sample of infected individuals. As a result, the observed frequencies are affected by sampling noise, introducing biases into the least squares estimation, as demonstrated in SI Sec. S.2.4.

To avoid sampling biases, we employ a hidden Markov model (HMM, sketched in Fig. 2A). The HMM analyzes the entire trajectory of the time series, treating true frequencies as hidden states and incorporating genetic drift and sampling error as Gaussian noise processes. Posterior distributions of the importation rates and the noise strengths are inferred using a Markov Chain Monte Carlo (MCMC) method. To speed up training, we have also implemented an Expectation-Maximization (EM) algorithm. For tracking neutral lineages, mutations are used as a tree-free alternative, with clustering techniques employed to ensure neutrality and reduce statistical errors (33). Further details about the inference method and the data processing are reported in Sec. 1.1–1.3 and SI Sec. S.2.

**Fig. 2.**
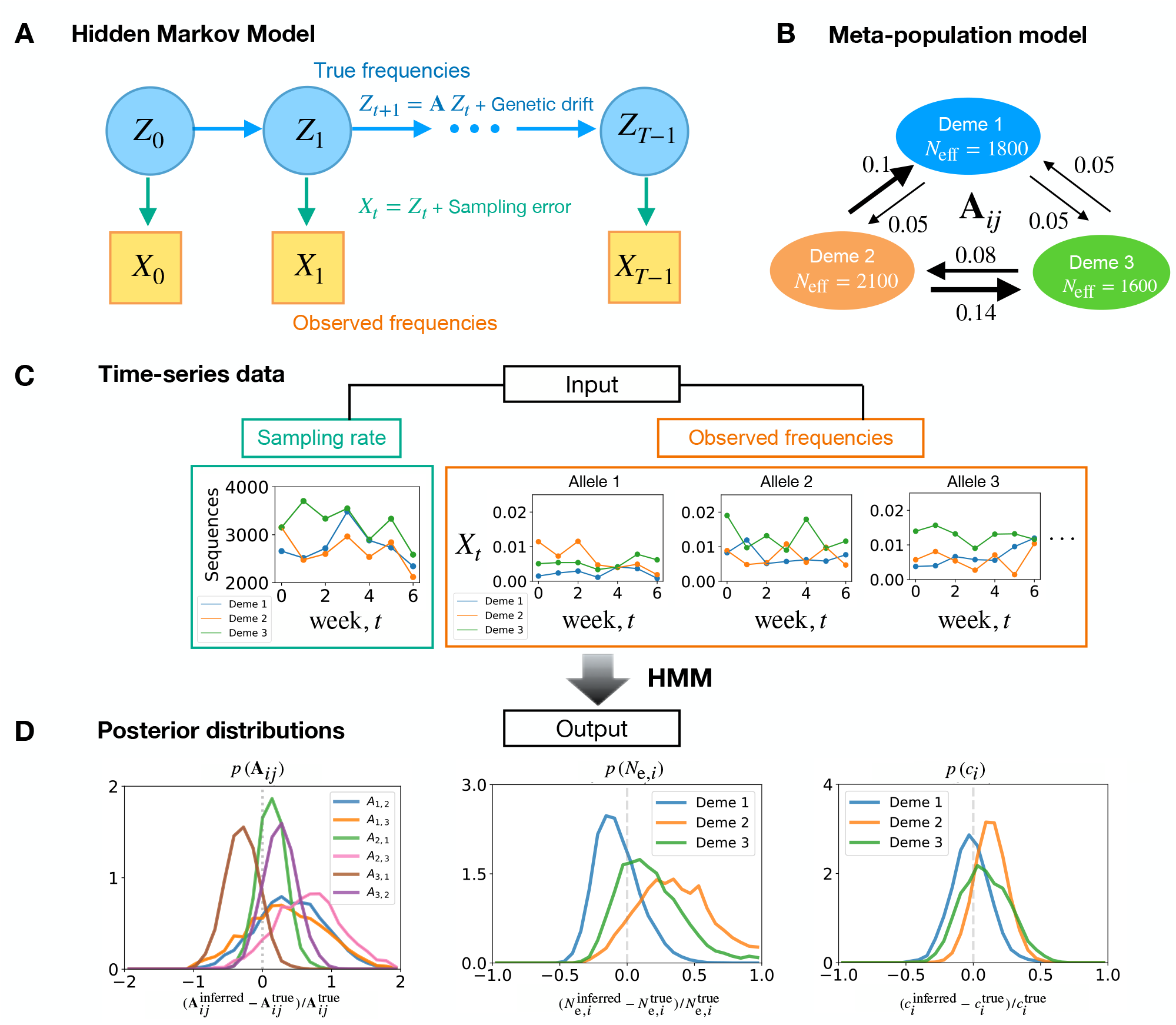
Inference method overview. We use a hidden Markov model (HMM) with continuous hidden and observed states (a Kalman filter (30)) to infer transmission networks from allele frequency time series. (**A**) A schematic of the HMM for the frequency dynamics. (**B**) To demonstrate the utility of our approach, we here use the depicted 3-deme model to generate allele frequency time series as input for the inference of the importation rates (arrows). In these simulations, the frequencies are evolved according to Eq. 5, with their initial frequencies being the same as those observed in EE, LDN, and SE of England during the week of June 20-26, 2021. We used effective population sizes comparable to those measured for England regions during the Delta wave period (31). The measurement noise overdispersion parameter (defined in the Methods section) is set higher than the value actually inferred, for illustrative purposes. (**C**) The input for the inference comprises weekly sampled sequences (left) and observed frequencies of lineages (right) within a focal variant. (**D**) The output is posterior distributions of the 6 importation rates of the 3 *×* 3 network (left), of the local inferred population size (middle), and of the deviation from uniform sampling (right). Each plot has a vertical dashed line, indicating the true value of the considered observable.

In Figs. 2B-D, we evaluate the effectiveness of our method to infer a known importation-rate matrix from synthetic data simulated under a metapopulation model of three demes interacting through a 3 × 3 matrix. The simulation duration (7 weeks), number of lineages, and effective population size were chosen to reflect the conditions during the Delta plateau period (Aug 2021 – Dec 2021) in England (31).

### Application to SARS-CoV-2 in England

To apply our method to real-world data, we first focus on England due to its large number of sequenced SARS-CoV-2 cases since the early stages of the pandemic (SI Fig. S25). The fall of 2021 constitutes a particularly suitable test case because of its consistently high and relatively stable incident numbers over more than four months (SI Fig. S26). We initially concentrate on this plateau phase of the Delta wave because it offers long allele frequency time series data with relatively low statistical error. In later sections, we also apply our method to the Alpha and Omicron waves in England, as well as the Delta wave in the USA.

Following our observations of regional allele frequency fluctuations in England (Figs. 1C-E), we further subdivided each region to create a total of 50 subunits, which we call demes, to enhance the spatial resolution of our interaction networks. Our subdivision algorithm ensures that each deme contributes a roughly similar number of sequences (see SI Sec. S.9.1). In SI Sec. S.4, we provide evidence that a finer spatial resolution leads to less reliable results, given the data we have.

Since sampling and incidence reporting typically follow a weekly cycle, we run our inference with a time step of one week (Δ*t* = 1). This time step aligns well with the generation time of SARS-CoV-2, which has an average infectious period of approximately 4–6 days depending on the variant (34, 35).

### Transmission networks mirror geography

After inferring the importation-rate matrix **A**, we performed hierarchical clustering to order the 50 demes by the similarity of their transmission networks, as quantified using the Jensen-Shannon divergence between their respective rows (SI Sec. S.7). Remarkably, the resulting matrix exhibits a pronounced block structure (Fig. 3A) with different blocks corresponding to geographically well-connected regions, such as the demes within London or near Liverpool (NE-1) and Manchester (NE-2). When we plot the major infection paths (the largest 5% of the off-diagonal matrix elements) on a map of England (Fig. 3B), a network of predominantly local connections becomes apparent. This aligns with a general decline of importation rates with distance and larger importation rates within than between regions (Fig. 3C).

**Fig. 3.**
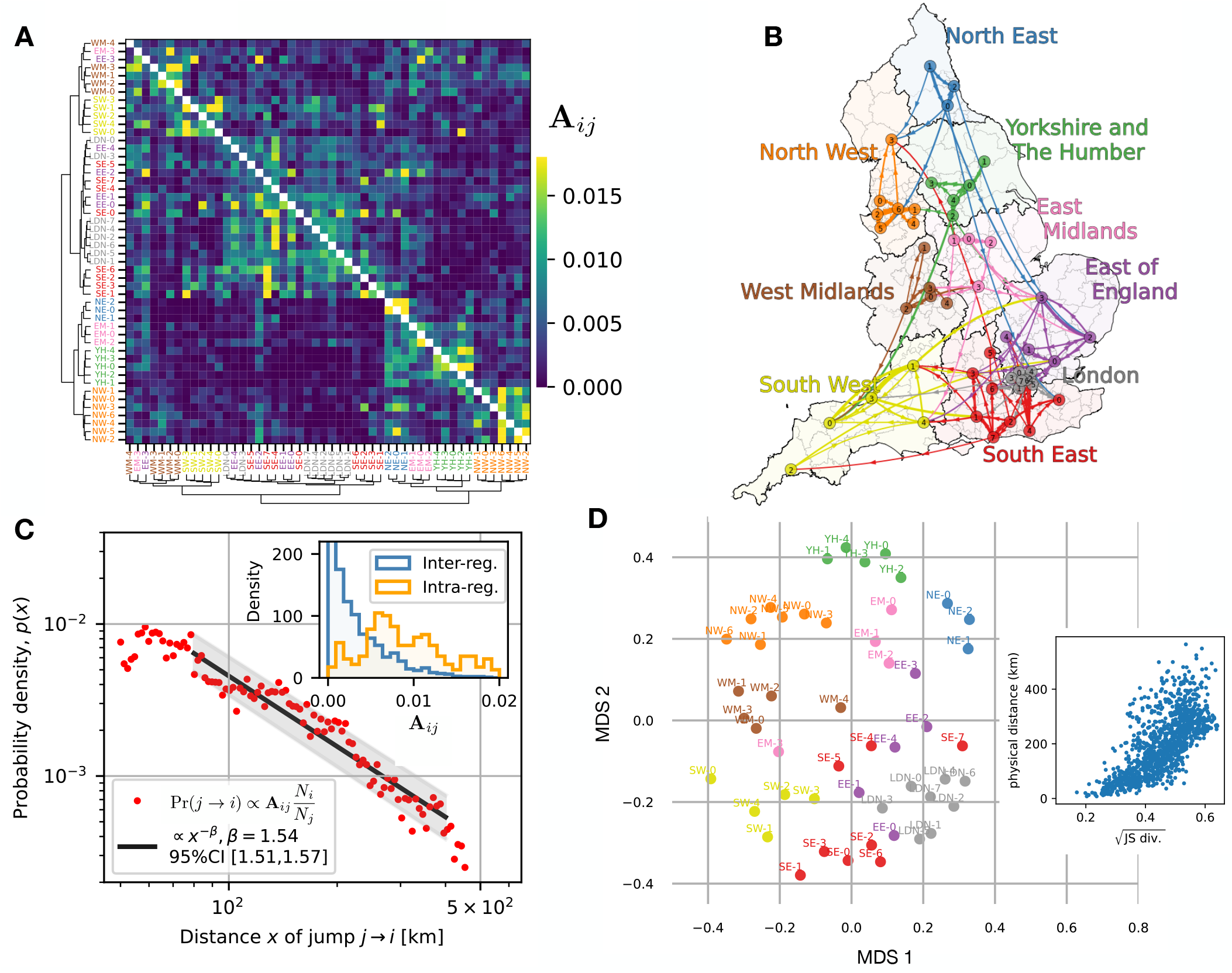
Network inference reveals heterogeneity, distance dependence and stochasticity of disease transmission. (England, Delta wave period, from Jun 20 to Sep 25, 2021). Heat map of the matrix **A**. The matrix coefficient **A**_*ij*_ represents the proportion of infections that population *i* imports from population *j*, thus encoding cross-infection rates between regions. Hierarchical clustering was used to order the 50 demes according to the similarity between rows of the matrix, as quantified by the Jensen-Shannon divergence (see SI Sec. S.6). (**B**) Illustration of the main infection pathways (the largest 5% of the off-diagonal matrix elements). (**C**) Inferred probability distribution of jump distances: A jump from deme *j* to deme *i* is assumed to occur with probability *∝* **A**_*ij*_ *N*_*i*_*/N*_*j*_, where *N*_*i*_ represents the population size of deme *i* (see SI Sec. S.6 for the underlying rationale). Inset: Histogram of matrix elements within and across regions. (**D**) Multidimensional scaling reveals an approximately geographic arrangement of populations on a two-dimensional plane. MDS is a procedure that projects high-dimensional data on a plane so as to maintain pair-wise distances as closely as possible. Here, pairwise distances are taken to be the square roots of Jensen-Shannon divergence between the rows of the matrix presented in Fig. **A**. In the right-side plot, the square root of Jensen-Shannon distance is compared with the physical distance (the Spearman correlation = 0.74).

To further explore how well epidemiological interactions mirror geographic relationships, we sought a two-dimensional representation of the entire network of importation rates. To this end, we performed a multidimensional scaling (MDS) analysis (36) to embed the locations of all the demes in a plane such that the in-plane distance between any two locations measures how different their vectors of importation rates are. The result, properly rotated, crudely resembles the map of England (Fig. 3D). Thus, frequency fluctuations encode geographic relationships even beyond the pairwise level (see SI Sec. S.7 for details of the MDS and further analyses).

### Transmission networks are asymmetric and evolve

The planar MDS representation only provides a time-averaged picture and also ignores any asymmetry in the epidemiological interactions among populations. To investigate the dynamics and asymmetry of infection, we focus on London (LDN) and its neighboring regions East of England (EE) and South East (SE). Applying our method to the Alpha, Delta, and Omicron waves results in the inter-community transmission rate matrices illustrated in Figs. 4A-C. Asymmetry is evident in all cases, with notable changes between waves. We find that the relative influence of different regions shifted across waves. During the Alpha and Delta waves, London generally had a stronger impact on South East and East of England than vice versa. Interestingly, within waves, we detect only relatively modest shifts (Fig. 4D), such as the growing influence of London on South East and East of England from July to October during the Delta wave.

**Fig. 4.**
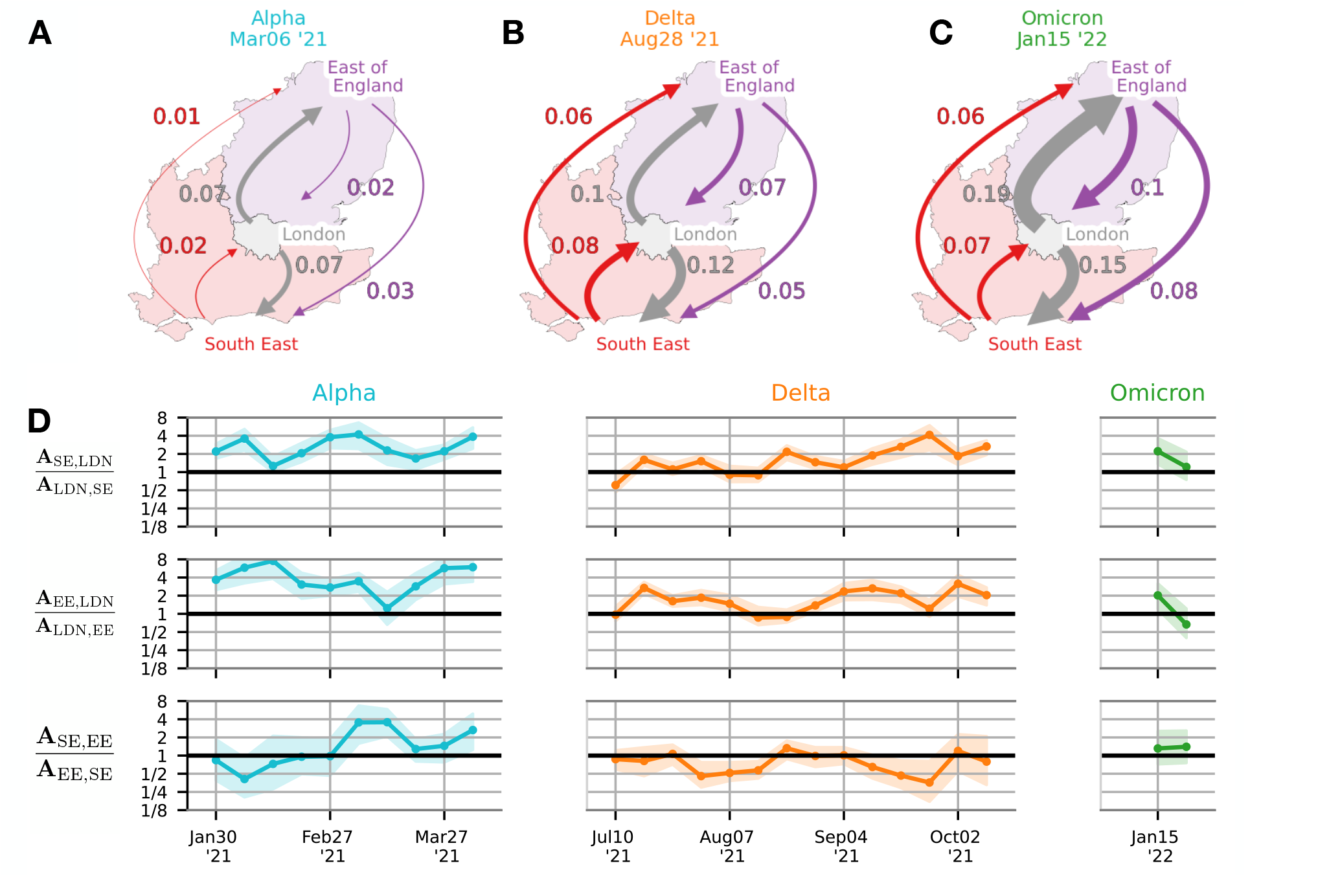
Time-dependence of transmission networks. (**A**-**C**) The 3×3 importation-rate matrix between London and its two neighboring regions, East of England and South of England, is inferred using the HMM-MCMC method. Arrows represent the mean importation rates for the Alpha, Delta, and Omicron variants. (**D**) Time series showing the ratio of importation rates between each pair of regions, with shaded areas indicating the upper and lower quartiles. At each timepoint, the importation-rate matrix is inferred using a time window of 7 weeks centered around that timepoint.

### Inference indicates pronounced genetic drift and uniform sampling noise

Our HMM framework not only estimates importation rates but also quantifies the strength of genetic drift via the effective population size, *N*_e,*i*_, and assesses the impact of sampling noise through the overdispersion parameter, *c*_*i*_. We compared the inferred effective population size with the number of infected individuals estimated by the COVID-19 Infection Survey (37). While there is a strong correlation between the two, the inferred effective population size was consistently lower by factors ranging from 18 to 29 across regions (Fig. 5). This discrepancy between effective and infected population sizes, which aligns with earlier studies aggregating over all England (31), may be explained by factors such as superspreading events and community structure (31, 38–43), as well as potential mutational fitness effects, although significantly non-neutral alleles were removed prior to the inference. In contrast, the overdispersion parameter *c*_*i*_ was close to one (Fig. S18), indicating that sampling error is uniform and of expected magnitude in the COVID-19 Infection Survey.

**Fig. 5.**
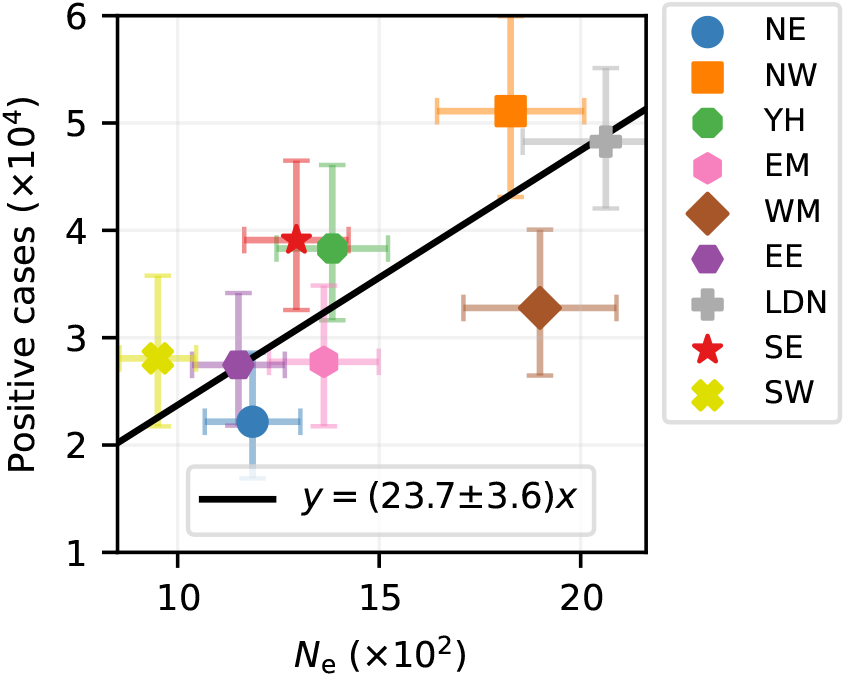
Stochasticity of disease transmission, quantified by the effective population size. For the Delta wave in England, the inferred effective population size at the region level (x-axis) is compared to the estimated number of infected individuals (y-axis). The Pearson’s correlation is 0.73 (p-value = 0.026). The error bars represent the 95% confidence interval (CI), and the solid line indicates the best linear fit.

### Comparison with the USA and mobility proxies

For comparison, we also applied our method to the USA, focusing on the Delta wave (Jul 18 – Oct 30, 2021). We divided the USA into 30 subunits using the same subdivision algorithm employed in the analysis of England (SI Sec. S.9.2). Fig. 6A illustrates major importation pathways. Similar to England, we found that inter-community importation rates in the US mirror geographic relationships (Fig. S24).

**Fig. 6.**
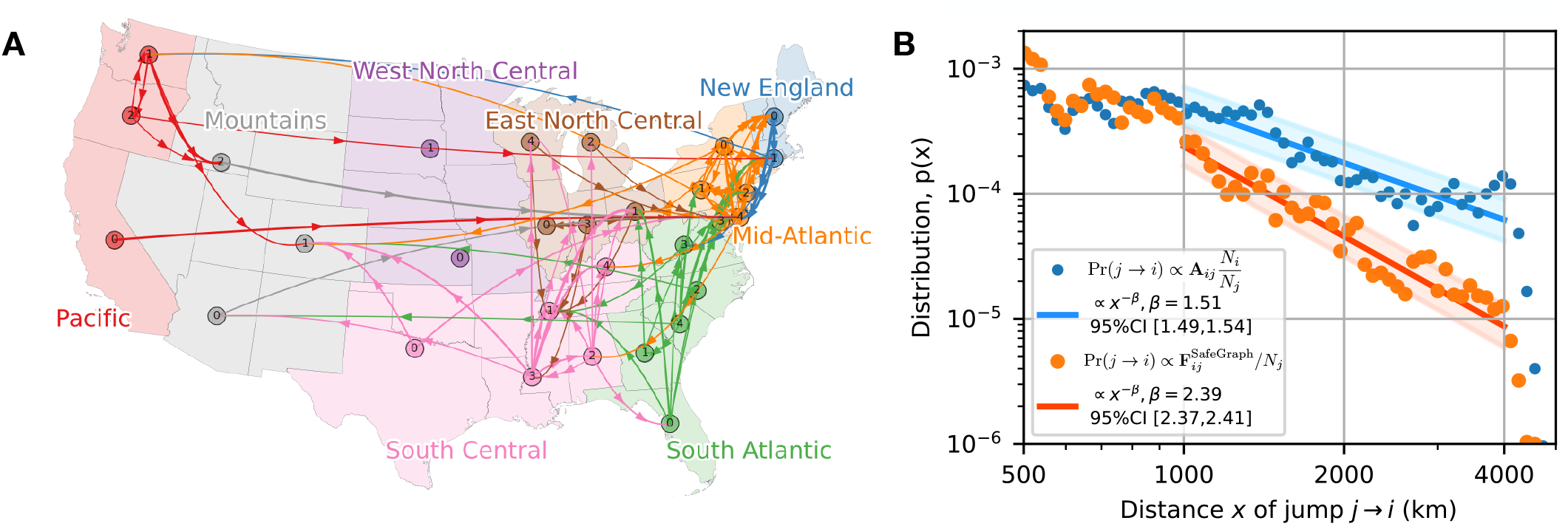
Transmission network in the USA during the Delta wave (30 demes, Jul 18, 2021 – Oct 30, 2021) and comparison to mobility data. (**A**) Illustration of the main infection pathways. Arrows indicate the largest 5% of the off-diagonal matrix elements **A**_*ij*_. See Fig. S23 for the inferred matrix. (**B**) Comparison between the jump kernel inferred from the 30 *×* 30 importation-rate matrix (blue) and indirectly estimated using the SafeGraph data (orange).

The US data also allow us to compare the jump kernel with what would be expected based solely on mobility data collected by SafeGraph during the period of January to May 2020 (44). SafeGraph gathers data from a large panel of anonymous mobile devices in the US, recording an average “home” location over six weeks and all locations where the device pauses for at least one minute. To convert these data to the lineage jump probability, we assume that a lineage jumps from deme *i* to deme *j* with a probability proportional to 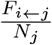, where *F*_*i*←*j*_ is the rate of trips from deme *j* to *i*, and *N*_*j*_ is the population size of source deme *j* (see SI Sec. S.1.4 for details).

Our analysis shows that the jump kernel directly inferred from the sequencing data decays substantially slower with distance than predicted by the mobility proxy data (Fig. 6B). This suggests that deducing epidemiological interactions from mobility data may require a different model or different information than the aggregated cell phone movements provided by SafeGraph.

### Eigenvalue decomposition

Given the potentially large number of inferred importation rates—2,500 in the case of England—it is important to determine which aspects of the transmission network are truly relevant for predicting epidemiological trajectories. Intuitively, what is relevant should depend on the timescale of interest. For instance, if we are concerned with the dynamics from one viral generation to the next, we need a detailed transmission network that captures the strongest importation rates, such as those within a city. Conversely, if we are interested in the dynamics over several weeks, the fine structure is less critical. After a brief relaxation period, the different parts of a city with strong intra-city connections will fluctuate coherently, allowing cities to be effectively represented as single compartments with weaker inter-city links.

Our dynamical model of neutral frequencies enables us to formalize the intuitive concept of timescale-dependent collective variations. First, note from Eq. 1 that our best prediction for the allele frequencies at future time *t* is given by the *t*^th^ power of the matrix **A** applied to the current allele frequencies,

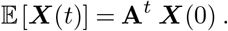

To compute the matrix power on the right-hand side, it is natural to decompose the matrix **A** in terms of its eigenvalues, upon which we get

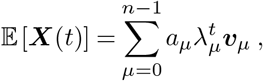

where ***v***_*µ*_ is the right eigenvector of **A** corresponding to the eigenvalue *λ*_*µ*_. The mode amplitudes *a*_*µ*_ are determined by the inner product ⟨***u***_*µ*_ |***X***(0)⟩ between the *µ*^th^ left eigenvector ***u***_*µ*_ and the initial frequency vector ***X***(0). We assume that the eigenvalues are arranged in descending order of magnitude and that the eigenvectors are normalized such that 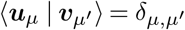.

We expect the dynamics to mix all demes over long times, unless there are isolated pockets, leading to an eventual equalization of the expected allele frequencies across demes. This outcome is guaranteed (i) if **A** has an eigenvalue *λ*_0_ = 1 corresponding to a right eigenvector of constant frequency *X*_*i*_ = 1 and (ii) if the magnitudes of all other eigenvalues are less than 1, |*λ*_*µ>*0_| *<* 1. These conditions are mathematically ensured by the right-stochasticity of **A**, provided that it is also irreducible.

### Relaxation dynamics

Before discussing the long-term stationary state, we first address the relaxation toward stationarity. The amplitude of mode *µ* decays e-fold after a time *τ*_*µ*_ ≡ −ln^−1^(|*λ*_*µ*_|), which diverges as |*λ*_*µ*_| approaches unity. Generally, many eigenmodes influence the relaxation process, as shown in Fig. 7A for the Delta wave in England. However, the total time to reach the steady state is controlled by the longest relaxation time *τ*_1_.

**Fig. 7.**
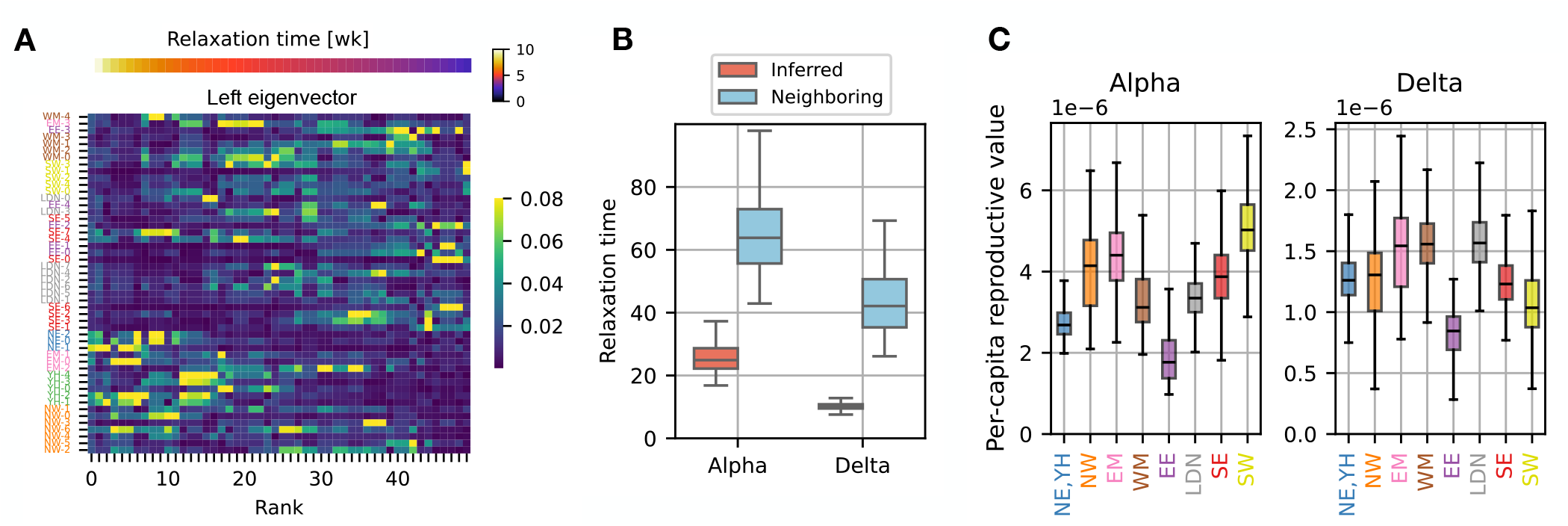
Relaxation dynamics and reproductive values. (**A**) The absolute values of the left eigenvectors ***v***_*n*_ of the 50 *×* 50 matrix during the Delta wave in England, presented in Fig. 3A. Each eigenvector ***v***_*n*_ is normalized such that Σ_*i*_ | (***v***_*n*_)_*i*_|= 1. (**B**) The longest epidemiological relaxation times [week] is calculated for each major variant in England from the regional importation-rate matrix. For comparison, we also show the relaxation times obtained from a modified matrix where transmission between non-neighboring regions is excluded. (**C**) The per-capita reproductive value for the Alpha and Delta variant in England. Here, the reproductive value*π*_*i*_ is evaluated by inferring the importation-rate matrix **A** at the region level, where NE, which has fewer sequences, is combined with YH to obtain reliable results. The values of *π*_*i*_ moderately depend on the choice of time intervals used for the inference (see Fig. S19).

In Fig. 7B, we display *τ*_1_ for the Alpha, Delta, and Omi-cron waves in England. The relaxation times differ significantly between waves; for instance, the relaxation time during the Delta wave is approximately 10 weeks, which is less than half of the relaxation time during the Alpha wave.

What features of the transmission network determine the overall relaxation time? The strongest interactions, typically short-range between neighboring demes, are certainly significant. However, we also identified relatively weak long-distance connections, whose impact is less clear. To assess their effect, we artificially removed all inferred long-distance connections (those spanning non-neighboring regions) and reevaluated the relaxation spectrum. We found that eliminating these long-distance interactions significantly increased *τ*_1_, by more than fourfold during the Delta wave, for example. This finding suggests that even very infrequent longdistance connections must be considered to accurately predict the speed of a pandemic.

### Equilibration and reproductive value

To illuminate the long-term stationary state, let us consider a neutral allele initially fixed in deme *i* and absent in all other demes. Due to case importations, allele frequencies are expected to gradually even out across demes. Consequently, the long-term expected frequency, *π*_*i*_, is the same in all demes and depends only on the identity *i* of the initial deme. The vector ***π*** = (*π*_1_, …, *π*_*n*_)^⊤^, composed of these primary-deme-dependent long-term frequencies, is proportional to the leading left eigenvector previously denoted by ***u***_**0**_ and normalized to sum to 1, Σ_*i*_ *π*_*i*_ = 1. *π*_*i*_ represents the expected contribution of deme *i* to the future gene pool, which is called the class reproductive value (45, 46).

When we normalize the reproductive value of a deme *i* by its number *I*_*i*_ of infected individuals, as measured by the COVID-19 Infection Survey (37), we obtain the per-capita reproductive value *π*_*i*_*/I*_*i*_, which is the probability that a viral genome picked at random from an infected individual in the distant future traces its ancestry back to this individual in the present generation (45, 46). If the infection dynamics were the same everywhere in England, one would expect individual reproductive values to be the same across all regions. Interestingly, we find that per-capita reproductive values vary by about a factor of 2 across England in both the Alpha and the Delta wave, see Fig. 7C.

In the same data set, we find that ratios of reproductive values approximately predict ratios of case importation rates (Sec. 1.4 and SI Sec. S.3),

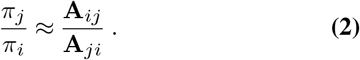

This relation with an equal sign is called detailed balance. Under most epidemiological models, this condition is expected because otherwise the lineage jump process exhibits unwarranted cyclic dynamics in equilibrium. Thus, finding detailed balance is reassuring of basic assumptions in epidemiological modeling.

Additionally, detailed balance, Eq. 2, helps relate heterogeneities in reproductive value to heterogeneities in transmission coefficients. For example, a lower per-capita reproductive value for East of England (EE) is observed both during the Alpha and Delta waves in England (Fig. 7C right), suggesting that the per-capita importation rate from EE to any other region is systematically lower than the other way around. In SI Sec. S.3, we further investigate these importation-rate patterns, demonstrating that the heterogeneities suggested by the reproductive values can be confirmed directly from the importation-rate matrix. We also argue that possible variations in reporting timing could affect heterogeneous patterns of *π*_*i*_, highlighting the importance of accurate reporting dates for precise inference (Fig. S11).

### Selected variants

We have previously emphasized that inferring **A**_*ij*_ sheds light on case importation patterns, their variations across time and space, their potential for control, and their role in embedding phylogenetic trees. But knowing the **A**_*ij*_ of a variant is useful also because it enables predicting its behavior if it is under selection. In the SI Sec. S.5, we show how selective forces modify the allele frequency dynamics in an SIR or SEIR model. In particular, when a variant under positive selection is rare, its dynamics follows the neutral dynamics in Eq. 5, with the modification **A**_*ii*_ → **A**_*ii*_ + *σ*, where *σ* is the selected advantage of the considered variant. Thus, as long as the variant is rare, its expected frequency is computed as:

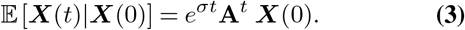

The kymograph in Fig. 8A illustrates the spreading pattern of the Delta variant across England. We conducted simulations of the Delta variant’s frequency dynamics (detailed in SI Sec. S.8), starting from frequencies observed in the week of March 7-13, 2021, when Delta variant sequences were reported solely from the deme labeled “London-0.” Fig. 8B displays the simulated abundance of the Delta variant using the inferred **A**_*ij*_, presented in Fig. 3A, along with fit parameters associated with the relative infectivity of the Delta variant (SI Sec. S.8). To assess the impact of long-range transmission, we artificially removed the inferred connections between non-neighboring regions and then conducted the same simulation (Fig. 8C).

**Fig. 8.**
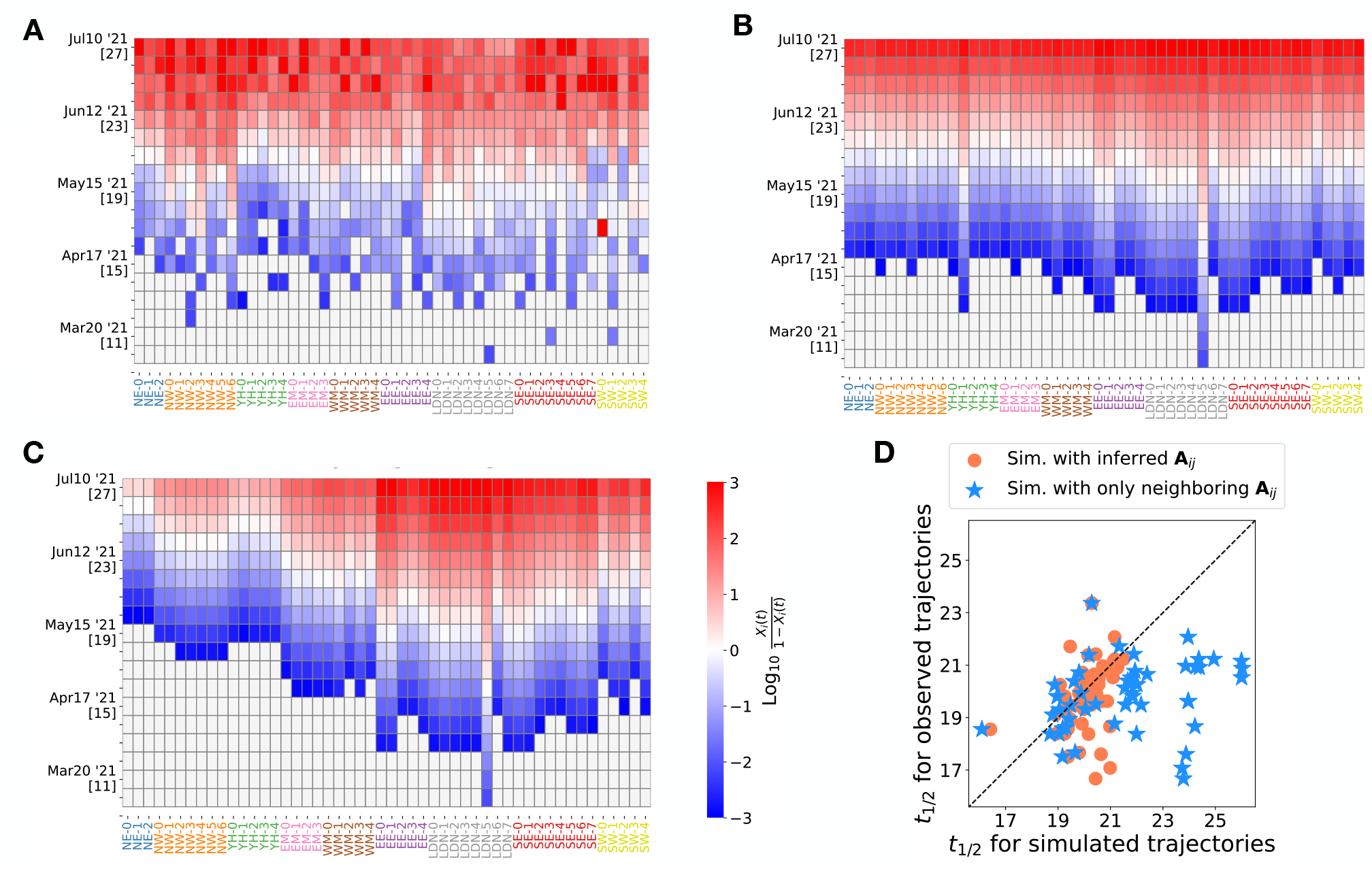
Replaying the dynamics of a selective sweep. (**A**) Observed frequencies of the Delta variant in England, ranging between 10^−3^and 1 − 10^−3^, across *N*_*D*_ = 50 locations in England (movies in SI). The heatmap displays the frequencies on a logit scale. On the vertical axis, epidemiological weeks (epiweeks) in 2021 are indicated below the dates by [*·*]. (**B**) Simulations following the non-neutral transmission model described in SI Sec. S.8, with inter-community importation rates as inferred for the Delta wave (shown in Fig. 3) (movie in SI). The observed frequencies at epiweek 10, when the Delta variant was reported only in deme “London-0”, are used as the initial condition (see SI Sec. S.8 for the fitting process). (**C**) Simulated frequencies, where transmission between non-neighboring regions (such as London and North West) is switched off (movie in SI). (**D**) Comparison of the midpoint epiweek 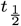—the week when the frequency in a deme reaches 0.5—between observed data and simulated data for each deme (see SI Sec. S.8 for the determination of *t* 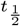). Orange circles represent 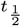 from the simulation with the inferred **A**_*ij*_, while blue stars represent 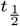 from the simulation without transmission between non-neighboring regions. The horizontal bars indicate the standard deviations obtained from the bootstrapping method.

Figure 8D shows the comparison of the midpoint time, *t*_1*/*2_, at which the abundance of the Delta variant reached 50% in a deme, between the observed and simulated data. The simulation using the inferred matrix roughly reproduces observed *t*_1*/*2_, clustering around epiweeks 18-21. In contrast, without transmission between non-neighboring regions, the simulation show delays in *t*_1*/*2_. In particular, for the NE, NW, and YH demes, which are distant from the initial seeding at “London-0,” the *t*_1*/*2_ values are predicted to be delayed by 2-6 weeks without the non-neighboring transmission. This highlights the significant impact of rate, long-range transmission on the spread of the new variant.

## Discussion

The infection rates between individuals from different population segments are among the many known unknowns of a pandemic. These rates, crucial for forecasting pathogenic spread, depend on complex behavioral patterns, immunity levels, and non-pharmaceutical interventions, rendering them exceptionally challenging to measure directly.

We here proposed a novel approach to infer these elusive rates retrospectively by examining the dynamics of viral ge-netic variation among different population groups. Utilizing this method, we constructed transmission networks during the COVID-19 pandemic in England and the US. Our findings shed light on the heterogeneity and plasticity of transmission networks, which have implications for epidemiological forecasting and intervention strategies.

### Methodological Advances

Our approach hinges on the premise that the observed frequencies of neutral variants are influenced solely by inter-community transmission, random genetic drift, and observational noise. By averaging out these noises, we can isolate the impact of cross-transmission, which tends to reduce regional differences in allele frequencies. This convergence towards a common value is evident in the allele frequency data of SARS-CoV-2 (Fig. 1) and demonstrates a plausible distance dependence. To optimally capture this signal, we have implemented a Kalman filter, which enables us to infer importation rates, local effective population sizes, and the strength of observational noise. A key advantage of this method is that it only relies on allele frequency time series, bypassing the need to construct a phylogenetic tree, which can be computationally demanding, especially when dealing with polytomies. We validated our method using population genetics simulations of a metapopulation with parameters consistent with those of SARS-CoV-2.

### Epidemiological Insights

Our key finding is that, applied to highly sequenced SARS-CoV-2 populations, this method allows us to map an entire interaction network between different populations. Unlike methods based on proxies, this approach quantifies direct epidemiological interactions that can be integrated into models of the disease spread. Our findings reveal substantial heterogeneity and plasticity in disease transmission networks. By applying our method to the SARS-CoV-2 transmission data in England and the U.S., we uncovered several key patterns:

### Geographical Mirroring and Long-Range Interactions

The inferred transmission networks largely reflect geographical proximities. Cross-importation rates are strongest between neighboring regions and gradually weaken with increasing physical distance. Interestingly, when interaction strength is converted into a distance and a map is drawn based on this metric, it roughly represents the geographic layout of England. This suggests that interactions reflect geographic relationships.

The observed gradual weakening of interactions with distance likely reflects the limited traffic flow between distant regions. However, we did not find quantitative agreement between the distance decay of US traffic flows as estimated by SafeGraph and cross-importation rates. We see two potential reasons for the discrepancy: (i) SafeGraph data treats all visited locations equally, regardless of the duration of the visit, which could lead to an underestimation of the impact of long-distance visits. (ii) Long-distance trips may be associated with riskier behaviors, thereby increasing the likelihood of disease transmission. Therefore, the reduction in mobility flux with distance may be partially offset by an increase in infections caused by long-distance travelers. To separate these contributions, differently curated mobility data will be required.

### Dynamic Changes in Transmission Patterns

The crosscommunity importation rates and their directionalities exhibit considerable variation across different waves of variants of concern. These dynamics were evident in the varying importation rates between London, East of England, and South East during different waves, with noticeable shifts in transmission dominance between these regions. Such dynamic changes emphasize the need for adaptive modeling approaches that can accommodate evolving transmission patterns over time.

### Asymmetry in Cross-Importation Rates

Our analysis showed that cross-importation rates are often heterogeneous, indicating that certain regions exert a stronger influence on others. In general, epidemiological “rock-paper-scissor” interactions could exist between different regions. However, we found that such non-transitive interactions are not observed in our inferred importation rates, which approximately satisfy a certain symmetry property (detailed balance) that ensures that the lineage dynamics backward is time reversible (no cyclic fluxes). This also allows using per-capita reproductive values (Fig. 7C) to compare the infectivity across regions.

### Implications for Epidemic Forecasting and Future Directions

The ability to directly infer detailed transmission networks has the potential to improve epidemic forecasting. We found that weak long-range interactions are crucial for explaining the spreading of beneficial mutations. This highlights the importance of accounting for such interactions in epidemiological models, especially for highly transmissible pathogens like SARS-CoV-2.

However, since importation-rate matrices were also found to change considerably between waves, continuous genomic surveillance will be needed to update cross-importation rates. With accumulating time series data for different waves and different pathogens, future studies might also learn to predict the evolution of cross-importation rates through time. Such progress might come either by identifying interpretable patterns with the help of mechanistic epidemiological models or through machine learning.

The patterns we have identified show that detailed balance is a reasonable assumption for epidemic models, but that different regions have different infectivities. Given detailed balance, one important open challenge is to understand mechanistically what sets the different infectivities of the different regions. Plausible candidate reasons are historical contingencies due to differential exposure to prior waves, inducing behavioral and immunological differences, or policy differences.

Understanding the detailed structure of transmission networks may also allow for the design of targeted interventions. For example, identifying regions with high cross-importation rates can help prioritize areas for vaccination, testing, and other control measures.

### Limitations and Assumptions of the Method

While our allele-frequency-based method offers substantial advancements, several limitations warrant discussion. Our method assumes that the alleles we track are neutral. Including alleles that are spatially sweeping and beneficial can lead to an overestimation of cross-importation rates from the origin to target areas. Therefore, it is crucial to exclude alleles whose selective changes are stronger than genetic drift, as we have done following our precursor work in ref. (31).

The absence of direct measurements for cross-importation rates means we lack a definitive benchmark for comparing our results. As a result, our inferred matrices should be interpreted with some caution. Nonetheless, the observed distance decay in our inferred inter-community transmission networks is a convincingly realistic feature, aligning with expected cross-importation rates. It is reassuring that our method infers a plausible tradeoff between transmission rate and geographic distance based solely on frequency correlations, without requiring geographical information.

Conversely, we have no way of verifying the weak longrange connections we inferred. Simulations with and without these interactions indicate their importance in facilitating rapid epidemic spread. So they need to be taken into account for prediction purposes. But our bootstrapping analysis also suggests that we cannot pinpoint the precise identity of these long-range connections: strong short-range interactions are consistently identified across bootstrap samples, but weak long-range connections vary (see SI Fig. S22). The uncertainty in the identity of these crucial weak links makes targeted intervention difficult, but hardly affect the predicted spreading time scales. Thus, while long-range interactions are significant for prediction purposes, their precise identities are not. Future work may develop a better representation for weak but important connections, for example, through systematic coarse-graining or other forms of regularization.

Our method does not require constructing a genealogical tree; it only relies on standing genetic variation monitored over time. The accuracy of our inferences improves with the amount of time series data available. Therefore, our method is most effective when applied to periods where a particular variant of concern is already prevalent. However, it is less effective in measuring cross-importation rates early on when a variant is just beginning to invade and few samples are available. Phylogeographical inference methods are better suited for source attribution, as they enable tracing the emergence of a new variant (26, 27). Nonetheless, our analysis indicated that cross-importation rates did not change significantly within waves, suggesting that our inferences may serve as a baseline for cross-importation rates in the early stages of a new wave.

## Methods

### 1.1 Inference methods

The simplest way to infer the cross-importation rates **A**_*ij*_(*t* + Δ*t*; *t*) in Eq. 1 for a fixed time difference Δ*t* from time series data is to minimize the least square difference between the predicted and actual lineage frequencies,

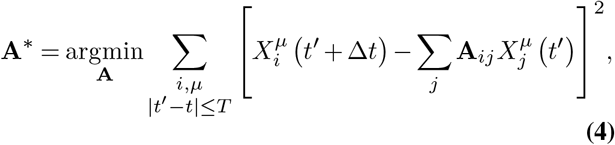

over all right stochastic *n* × *n* matrices, satisfying **A**_*ij*_ *>* 0, Σ_*j*_ **A**_*ij*_ = 1. The summation over time points *t*^*′*^on the right-hand side serves as a regularization step, which ensures that **A** does not vary on time spans smaller than 2*T*. The more sequencing data is available, the smaller *T* can be chosen, which leads to a better resolution of the temporal variations in **A**. Standard errors of the matrix coefficients are obtained by bootstrapping over the available lineages *µ*.

The linear regression approach in Eq. 4 is computationally efficient but requires as input the true lineage frequencies, which are never known exactly. Instead, one can only measure the frequencies within the sequenced sample, which represent the true frequencies distorted by sampling noise.

Accordingly, we have adopted a HMM, as depicted in Fig. 2, which treats the frequencies as hidden states. By modeling genetic drift and sampling noise as Gaussian distributions, the HMM effectively transforms into a computationally efficient Kalman filter (30) with a likelihood function that can be calculated analytically. The per-generation variance due to genetic drift is inversely proportional to the effective population size *N*_e_, which is usually smaller than the actual number of infected individuals. Specifically, for SARS-CoV-2 in England, the ratio between actual and effective population size was found to range from tens to hundreds (31). Likewise, sampling noise can be larger than expected based on random sampling, if sampling is not random but entails correlations. We therefore set the sampling variance to be inversely proportional to 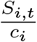, where *S*_*i,t*_ is the number of sequences sampled from population *i* at time *t*, and *c*_*i*_ measures the deviation from random sampling. To infer the strength of genetic drift (*N*_*e*_) and sampling noise (*c*_*i*_) from our HMM, we have implemented an MCMC algorithm, which yields posterior distributions for these parameters (see Sec. S.2 in SI).

Our simulation results in SI Fig. S7 show that, while both the least squares method and the HMM method with the MCMC algorithm retrieve importation-rate matrices close to the ground truth, the least squares estimation tends to overestimate small interactions. Such a bias appears from the fact that the solution of Eq. 4 tends toward the uniform matrix when noise levels are high.

We thus focused on the HMM method to minimize these biases. To reduce computational cost of the MCMC, we have also implemented an EM algorithm, which provides the maximum likelihood estimate of all relevant parameters. The inference error of the EM algorithm was assessed by using a bootstrapping approach, where the parameters were inferred multiple times from randomly created sets of alleles, each set maintaining the same size as the original set.

### 1.2 Neutral evolution in a metapopulation

Here, we provide additional mathematical rationale for why Eq. 1 describes the frequency dynamics of a neutral allele in a metapopulation. First, the deterministic terms in Eq. 1 are linear in the frequencies because, under neutrality, the frequency of any union of lineages must obey the same stochastic evolution equation. Additionally, neutrality implies that frequencies do not change in expectation if the frequency is the same in all populations, necessitating that each row of **A** sums up to 1. (If *X*_*i*_(*τ*) = *x* for all *i*, Eq. 1 implies 𝔼 [*X*_*i*_(*t*) | **X**(*τ*)] = *x* Σ_*j*_ **A**_*ij*_, which equals *x* only if Σ_*j*_ **A**_*ij*_ = 1.) Finally, negative matrix elements are excluded because they can generate negative frequencies.

Note that an alternative way of writing Eq. 1 is

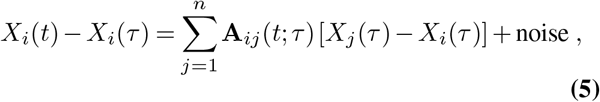

which explicitly shows (i) that the frequency in *j* only influences the frequency in *i* if *X*_*i*_ *≠ X*_*j*_, and (ii) that a larger value of **A**_*ij*_ *>* 0 leads to a faster convergence of *X*_*i*_ to *X*_*j*_.

Each coefficient **A**_*ij*_(*t*; *τ*) for *j ≠ i* denotes the proportion of infections that population *i* receives from population *j* during the interval from *τ* to *t*. Meanwhile, **A**_*ii*_(*t*; *τ*) = 1 − Σ_*j≠ i*_ **A**_*ij*_(*t*; *τ*) represents the proportion of infections that are not imported, i.e., “homegrown” infections.

From this perspective, it is straightforward to see how Eq. 5 arises. During the period from *τ* to *t*, population *i* imports a total of *I*^*i*^(*t*)**A**_*ij*_(*t*; *τ*) infections from population *j*, with a fraction *X*_*j*_(*τ*) of these infections carrying the focal allele. Thus, at time *t*, the expected total number of infections carrying the focal allele in population *i* is *I*_*i*_(*t*) _*j*_ **A**_*ij*_(*t*; *τ*)*X*_*j*_(*τ*). Consequently, the updated allele frequency *X*(*t*) is given by Σ_*j*_ **A**_*ij*_(*t*; *τ*)*X*_*j*_(*τ*). By substi-tuting **A**_*ii*_(*t*; *τ*) = 1 − Σ _*j≠i*_ **A**_*ij*_(*t*; *τ*), we obtain Eq. 5.

### 1.3 From lineages to alleles

Our inference technique is fueled by data collected through counting the prevalence of *independent* lineages across locations and times. Identifying these independent lineages is straightforward when a complete and accurate phylogenetic tree is available, as it simply involves segmenting the tree into monophyletic groups. But constructing a large phylogenetic tree in the first place—in the case of SARS-CoV-2 for millions of viral genomes—is not only computationally demanding but often results in unresolved polytomies.

A tree-free alternative for creating time series of neutral lineages is provided by tracking the frequency of all preexisting neutral *mutations* in the different sub-populations. This works in principle because the frequency of a neutral allele has to obey Eq. 1, even in the presence of recombination. The downside of this approach is that, due to linkage, different alleles may not be independent. Treating them as independent underestimates statistical errors. Moreover, it is important to ensure that the included alleles are neutral.

Therefore, we first cluster alleles based on their pairwise genetic distances and then select representative alleles for each cluster (33). We then excluded a small fraction of outlier alleles, whose time series are not consistent with neutrality, as measured by a maximum likelihood method described in refs. (31, 47) (also see SI Sec. S.1.3).

In SI Figs. S3 and S20, we demonstrate that, regardless of whether we use tree-based lineages or alleles (including only synonymous mutations or all mutations), the qualitative and coarse-grained outcomes (the largest eigenvalues and reproductive values) of both tree-based and allele-based approaches are consistent and do not significantly differ for the Delta wave in England. Additionally, in SI Fig. S21, we demonstrate that similar importation rates are inferred from a downsampled fraction of the sequences (e.g., 10%).

As an additional consistency check, we provide a theoretical explanation in SI Sec. S.4 on how the importation-rate matrix should change with spatial coarse-graining. We verified that the behavior of the inferred matrix aligns with these theoretical predictions, further supporting the robustness of our inference method.

### 1.4 Detailed balance condition

The inferred infection matrices are typically asymmetrical, which can be checked for example for the Delta wave matrix shown in Fig. 3a. However, one might wonder if a more general symmetry, known as detailed balance, is maintained, which is often assumed in modeling and inference studies. Detailed balance is satisfied when the flux of lineages between two regions is balanced when traced backward in time. This implies that the backward-time lineage jump process is time-reversible. Mathematically, this detailed balance condition can be expressed as

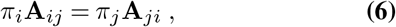

meaning that, in equilibrium, the lineage flux from region *j* to *i* is equal to the reverse flux. In Bayesian phylodynamic inference, detailed balance is often assumed to simplify the learning algorithms (48). Moreover, standard epidemiological models imply detailed balance, because otherwise the lineage jump process exhibits unwarranted cyclic dynamics in equilibrium. We show in SI Sec. S.5 how detailed balance emerges from a standard epidemiological SIR and SEIR models.

It is therefore important to check whether the data justifies the detailed balance premise. We focus on the nine regions in England and plot in Fig. 9 the ratio of left and right-hand sides in Eq. 6 for all possible pairwise interactions. While most long-distance interactions are too weak to test for detailed balance, the strong neighbor-neighbor interactions are largely consistent with detailed balance.

**Fig. 9.**
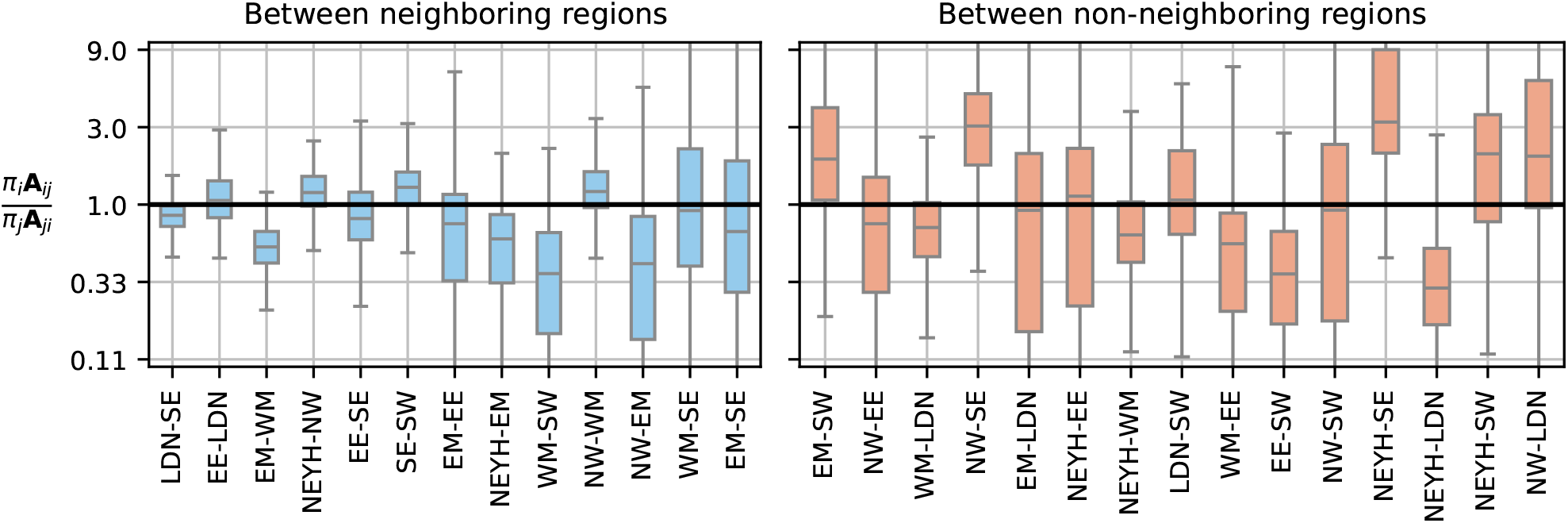
Testing the detailed balance assumption. Under detailed balance, the ratio 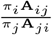 should equal one for all deme pairs (*i, j*), where *π*_*i*_ denotes the class reproductivevalue (satisfying 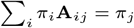 and normalized to 1). The box plots show these ratios for all pairs of regions in England, where YH and NE are combined to a single region, using the 8 *×* 8 importation-rate matrix **A**_*ij*_ inferred from the region-level data of mutations during the Delta wave period (June 20, 2021 - October 31, 2021). Results for neighboring region pairs, which generally exhibit stronger couplings, are shown separately from those for non-neighboring regions, where couplings are typically smaller and exhibit greater inference error in 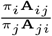.

## Data Availability

All data produced are available online at https://github.com/Hallatscheklab/NetworkInfer

https://github.com/Hallatscheklab/NetworkInfer

## Data Availability

All of the SARS-CoV-2 genomes analyzed in this article are publicly accessible through the GISAID platform and the COG-UK consortium. The GISAID accession identifiers analyzed in this study are provided as part of Supplementary Materials.

## Code Availability

The Python script for the HMM-EM method and the C++ code for the HMM-MCMC method, along with the Python scripts to reproduce the figures in this manuscript, are available at https://github.com/Hallatscheklab/NetworkInfer.

## ACKNOWLEDGMENTS

We are grateful the Hallatschek lab for helpful discussions and feedback. We are grateful to Moritz Kraemer, and Richard Neher for helpful discussions and feedback. TO acknowledges support from JSPS KAKENHI (Grant Numbers JP22K03453 and JP22K06347) and the RIKEN iTHEMS Program. OH acknowledges support by a Humboldt Professorship of the Alexander von Humboldt Foundation. QY acknowledges support from the National Science Foundation Graduate Research Fellowship under Grant No. DGE 1106400. GI acknowledges support of a Humboldt Research Fellowship by the Humboldt Foundation. This research used resources of the National Energy Research Scientific Computing Center (NERSC), a U.S. Department of Energy Office of Science User Facility located at Lawrence Berkeley National Laboratory, operated under Contract No. DE-AC02-05CH11231 using NERSC BER-ERCAP0019907. We gratefully acknowledge all data contributors, i.e., the Authors and their Originating laboratories responsible for obtaining the specimens, and their Submitting laboratories for generating the genetic sequence and metadata and sharing via the GISAID Initiative, on which this research is based. The authors acknowledge use of data generated through the COVID-19 Genomics Programme funded by the UK Department of Health and Social Care (https://www.gov.uk/government/organisations/department-of-health-and-social-care).

We thank the COG-UK consortium and all partners and contributors who are listed at https://webarchive.nationalarchives.gov.uk/ukgwa/20230507113711/https://www.cogconsortium.uk/about/about-us/about-us/.

## Supplementary Information

### S1 Data sources and processing

For the analysis of England, we downloaded the sequence metadata from the COVID-19 Genomics UK Consortium (COG-UK) (28) on March 25, 2022. The metadada include the time and location of sample collection. The number of sequences over time is presented in Fig. S25. For the analysis of the United States, the sequence metadata was obtained from the GISAID database (https://www.gisaid.org/). For the Delta variant analysis, we excluded the AY.4.2 sequences, whose proportion modestly increased in England during the Delta wave (49). For the Omicron variant analysis, we focused on the B.1.1.529 and BA.1 lineages, except for the BA.1 sequences that had any of the mutations S:K417N, S:N440K, or S:G446S (50).

#### S.1.1 Lineage frequency data

While the metadata include the lineage designation using the Pango nomenclature (51, 52), it classifies variants into a limited number of lineages. Therefore, we created our own lineages based on phylogenetic distance using the publicly available COG-UK phylogenetic trees (on March 25, 2022) (31); specifically, we cut the tree at a particular depth to create many subtrees, defining each subtree as a lineage (see Fig. S1A). If any subtrees occupy more than 2.5% of the total sequences, we introduce an additional cut at another position (farther from the root) and divide these subtrees further to create more subtrees (lineages). We continue this process until no subtree occupies more than 2.5% of the total sequences. For the analysis of the Delta variant, the tree was cut at the depths at 50.5*l*_unit_, 56.5*l*_unit_, 59.5*l*_unit_ where *l*_unit_ = 3.34 × 10^−5^. Fig. S1B) shows how the sequences are distributed along the depth of the tree over epiweeks. Note that in this figure and throughout the SI, we adopt epiweeks starting from December 29, 2019, extending their application across multiple years to enable continuous analysis despite the usual yearly reset. The correspondence between the epiweeks and calendar dates is summarized in Sec. S.10.

**Fig. S1.**
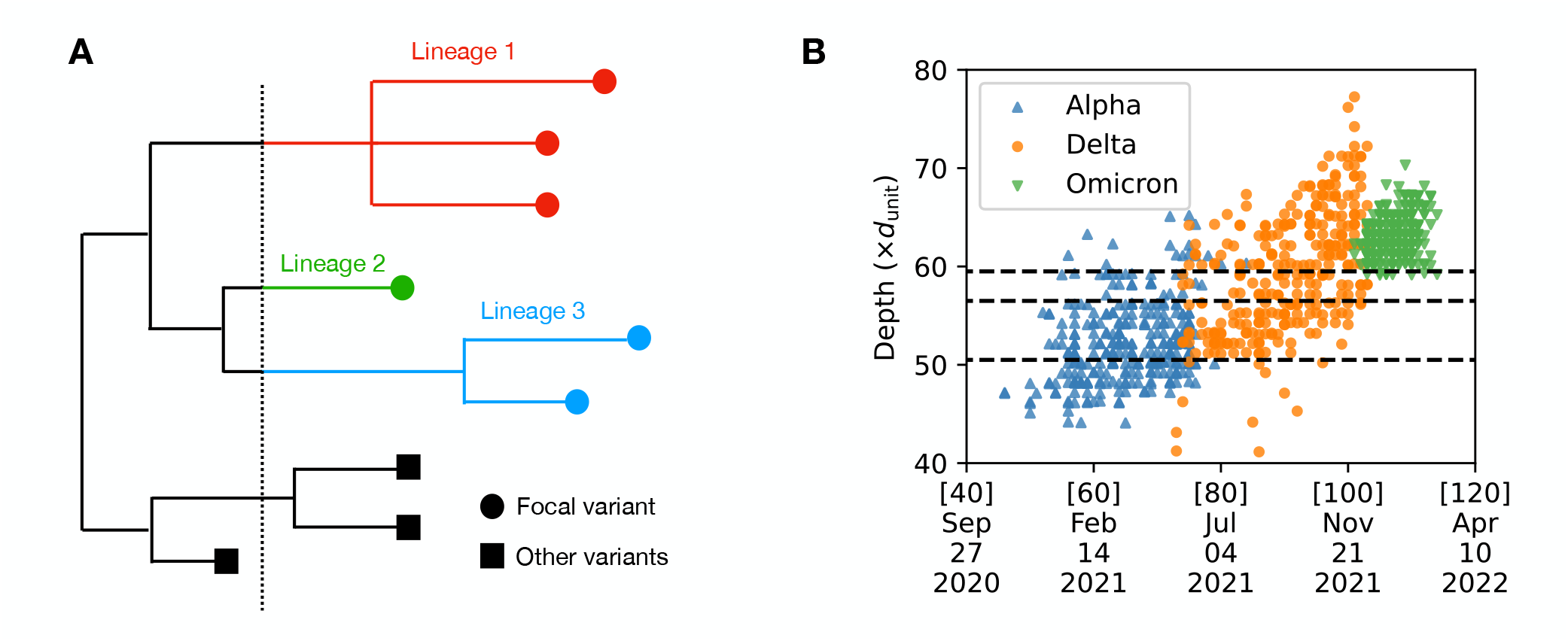
(**A**) Construction of lineages within a variant using the phylogenetic tree: Leaf nodes are represented by circles for sequences of the focal variant and by squares for sequences of other variants. Lineages of the focal variant are defined by cutting the tree at a certain depth, represented by a vertical dashed line. In this illustration, three lineages are obtained through this procedure. (**B**) Collection date versus tree depth for the metadata sequences of the Alpha, Delta, and Omicron variants: For each variant, only 300 sequences are displayed for clearer visualization. The dashed horizontal lines represent the cuts used to define the lineages of the Delta variant in England. The numbers [*·*] on the dates denote epiweeks.

#### S.1.2 Mutation frequency data

Mutation frequency data for England and the USA were generated from mutations listed in the COG-UK and GISAID metadata, respectively. The COG-UK metadata includes both synonymous and non-synonymous mutations, whereas the GISAID metadata includes only non-synonymous mutations. The frequency of each mutation was calculated by counting the number of sequences carrying that mutation and normalizing this by the total number of sequences at each sampled location, with a unit time of one week. For very rare mutations, the effect of sampling noise is significant, making the inference unreliable. Conversely, including very abundant mutations would limit the total number of independent mutations (obtained from a method described below (33)). Therefore, as a compromise, we decided to select mutations whose country-wide frequency, averaged over a focal time window, is moderately low, ranging between 0.003 and 0.05.

The presence of genetic linkage between mutations can create statistical dependencies that can distort our inference. To avoid bias due to linkage-induced correlations, we pruned the set of mutations following ref. (33). The pruning procedure first defines a distance between two mutations *m*_1_ and *m*_2_ as

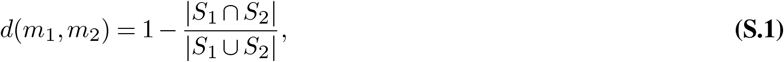

where *S*_*i*_ is the set of sequences carrying the mutation *m*_*i*_ in a country. Next, we constructed a graph where nodes are mutations, and an edge exists between two mutations if *d*(*m*_1_, *m*_2_) *< d*_th_, with *d*_th_∈ [0, 1] as a threshold parameter. We then identified the connected components of the graph, treating each connected component as a cluster of mutations. Finally, from each cluster, we selected the mutation *m*_*i*_ with the largest |*S*_*i*_| as its representative, producing a set of approximately independent mutations.

The higher the threshold value *d*_th_, the more mutation trajectories are grouped into the same cluster, as illustrated for the the Delta variant in England in Figure S2. We confirmed that as *d*_th_ increases, the importation-rate matrix for the Delta variant inferred from the mutation data becomes more similar to the matrix inferred from the lineage data (Figure S3). Based on these results, we decided to use a high value of *d*_th_ = 0.9 for all the inferences presented in this paper (except for Figure S3).

##### S.1.3 Outlier detection

In our inference, we focused on alleles within a particular variant, as our method relies on the neutrality assumption, and mixing alleles from different variants could introduce fitness differences that violate this assumption. However, there is still a possibility that significant differences in fitness exist between alleles even within a variant, which could potentially bias the inference of the importation-rate matrix. To prevent this, we applied the statistical test for neutrality (31, 47), which computes the maximum likelihood estimate of the relative fitness *s* and the *p* value for each allele, from the time-series data of allele counts.

**Fig. S2.**
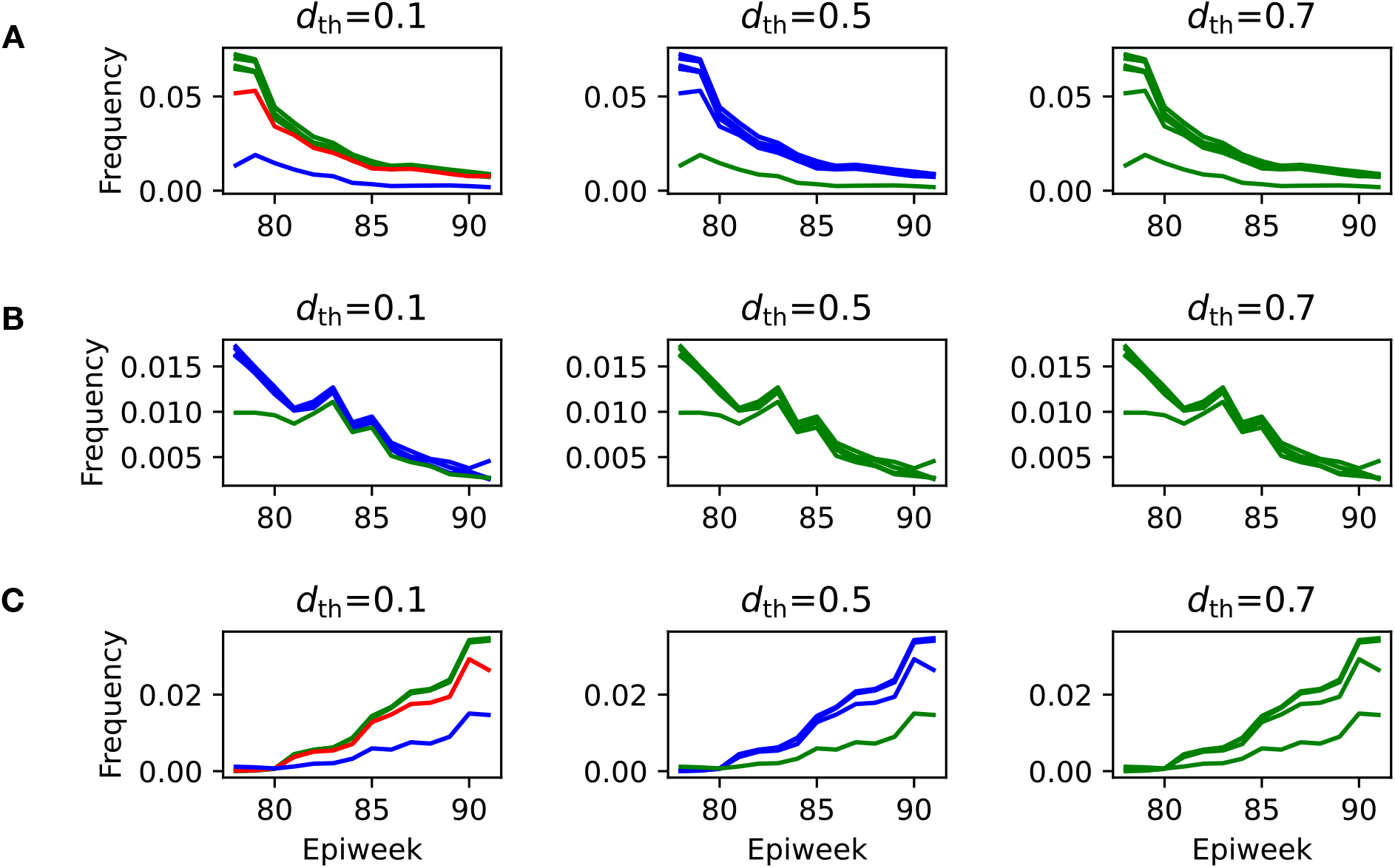
(**A**) The country-wide frequency trajectories of a subset of mutations of the Delta variant in England. For each threshold value *d*_th_, the frequency trajectories of mutations that belong to the same cluster are plotted in the same color. Nine mutations are classified into three, two, and one clusters for *d*_th_ = 0.1, 0.5, and 0.7, respectively. Similarly, Figures (**B**) and (**C**) show the frequency trajectories and the equivalence classes for other subsets of mutations of the Delta variant in England.

**Fig. S3.**
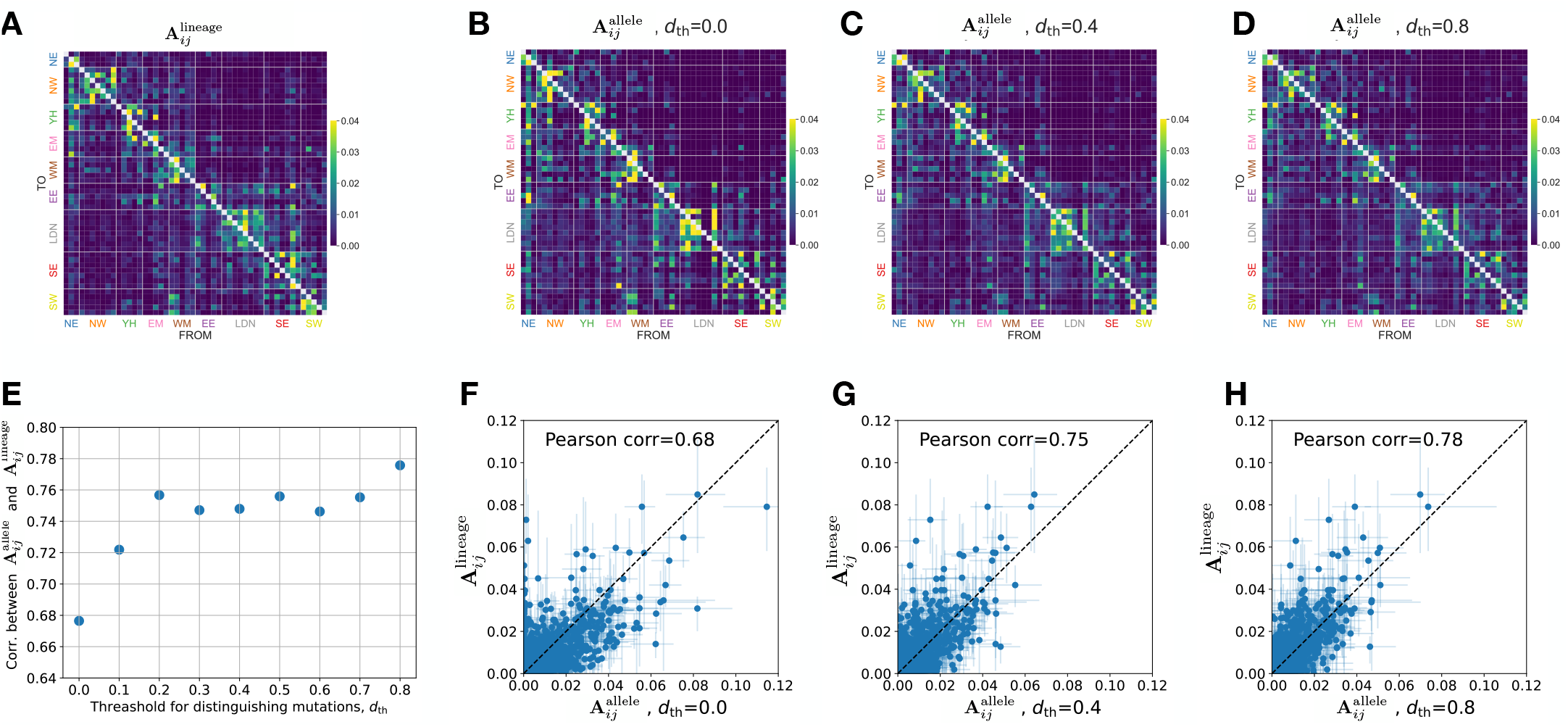
Comparison between the result for the Delta-sublineage data in England and the result for the mutation data, and the dependence on the threshold value *d*_th_ used for clustering mutations. *N*_*D*_ = 50. (**A**) Importation-rate matrix, 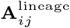, inferred from the lineage data. (**B**-**D**) Importation-rate matrix, 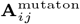, inferred from the mutation data, where *d*_th_ = 0.0, 0.4, 0.8, respectively. (**E**) *d*_th_ vs the Pearson correlation coefficient between 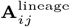 and 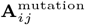. F-H) Element-wise comparison of 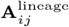 and 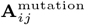 for *d*_th_ = 0.0, 0.4, 0.8, respectively.

For the mutations selected by the clustering described in Sec. S.1.2, we applied this statistical test using their country-wide count data. We then excluded significantly non-neutral mutations (*p <* 0.05) and used only putatively neutral mutations (*p*≥ 0.05) for our inference. Figure S4A displays the results of the statistical test on the Delta variant in England. From left to right, it shows the allele-frequency trajectories for putatively neutral alleles (left), significantly non-neutral alleles (middle), and the *p*-*s* distribution (right). Among the 85 representative mutations identified by the clustering method described in Sec. S.1.2, 68 mutations are identified as putatively neutral. As a control, we also applied the statistical test to the simulated data of neutral allele frequencies generated by the Wright-Fisher model (Fig. S4B). For both the actual and simulated data, the central region of the *p*-*s* distribution exhibits a triangular shape, indicating the validity of the neutrality assumption for the majority of mutations of the Delta variant sitting in this central region of the *p*-*s* distribution.

Similarly, we applied the same filtering to the lineage data and used only putatively neutral lineages for the inference.

**Fig. S4.**
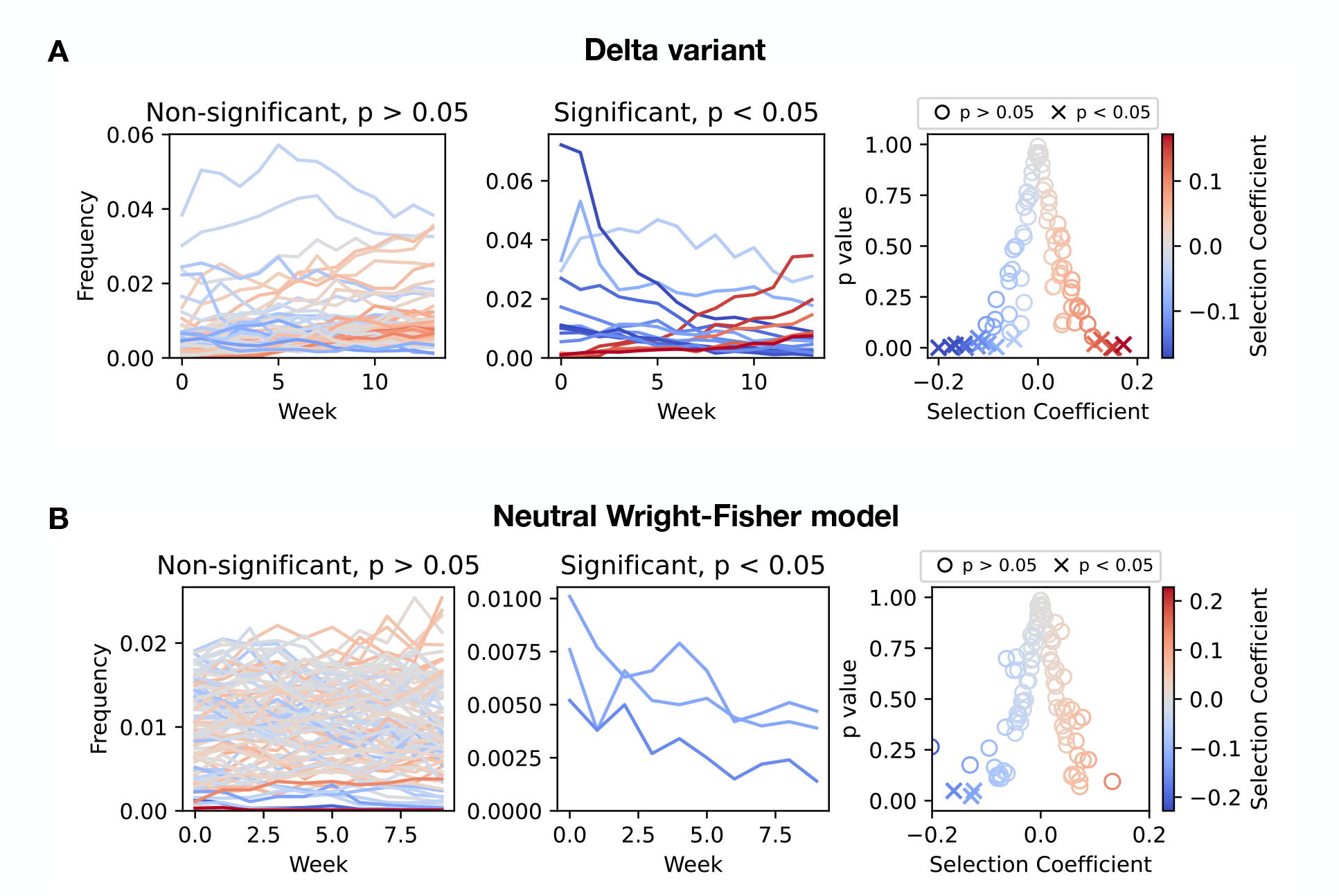
(**A**) Trajectories of mutation frequencies observed in the Delta wave in England (Left: trajectories with *p >* 0.05, Middle: trajectories with *p <* 0.05). Right: Inferred selective coefficient and p-value, where each dot represents these values for a particular mutation. (**B**) Trajectories generated by the neutral Wright-Fisher simulation (Left: trajectories with *p >* 0.05, Middle: trajectories with *p <* 0.05). The effective population size is set to 10,000, the sampling rate per week is set to 10,000, and the number of trajectories is 100, comparable to actual country-wide data in England. Right: Inferred selective coefficient and p-value.

##### S.1.4 Mobility data in the United States

In the analysis of the jump kernel presented in Fig. 6C, we used SafeGraph data (44), which were derived from cell phone tracking. Specifically, we used the county-level dataset processed by the authors of ref. (53) and then aggregated spatial locations to obtain the mobility flux between the 30 demes. Note that while we compared the jump kernel inferred from the sequencing data during the Delta wave with that calculated from the SafeGraph data, the SafeGraph dataset spans March 2020 to February 2021, which predates the surge of the Delta variant.

##### S.1.5 Autocorrelation functions, *R*_*ij*_, in Fig. 1D

In Fig. 1D, we demonstrate the convergence of allele frequencies across regions in England by computing the autocorrelation functions *R*_*ij*_(*τ*), defined as

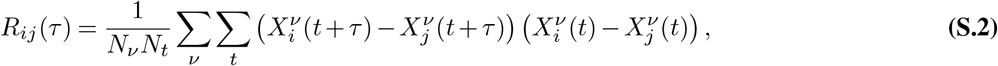

where *ν* and *t* label alleles and timepoints, respectively; *N*_*ν*_ and *N*_*t*_ denote the total numbers of alleles and timepoints, respectively; and 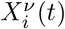 denotes the frequency of allele *µ* in region *i* at week *t*. We also calculated the autocorrelation functions using the lineage frequencies of the Delta variant and confirmed that decay rates similar to those presented in Fig. 1D are obtained from the lineage data (Fig. S5).

**Fig. S5.**
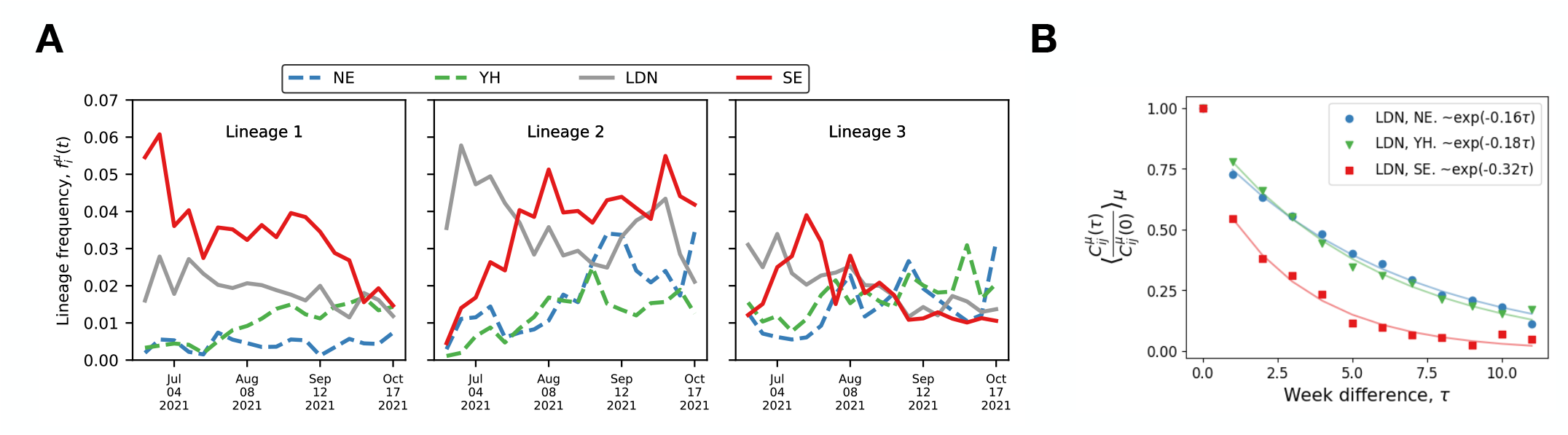
(**A**) The time-series trajectories of lineage frequencies are illustrated for three lineages of the Delta variant. (**B**) The dots show the normalized autocorrelation functions *R*_*ij*_ between London and North East (blue), London and Yorkshire and the Humber (green), London and South East (red), computed from the lineage frequency data. The solid lines show exponential curves fitted to the data. The auto-orrelation functions are computed by averaging over the 40 lineages with the largest sample sizes.

#### S.2 Hidden Markov Model for the neutral frequency dynamics

Our inference method is based on a dynamic linear model (a Kalman filter). We follow the notation and formulation in ref. (54). In the following, we denote the multivariate normal distribution with mean ***µ*** and covariance **Σ** as

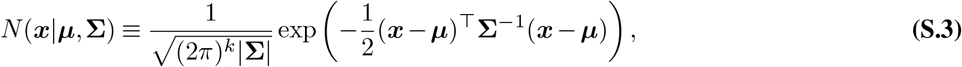

where *k* is the dimensionality of ***x***. We denote the function measuring “heterozygosity”, *x*(1 − *x*), as

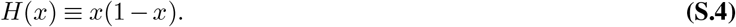

##### S.21 Hidden Markov Model

We consider the neural dynamics of a lineage (or allele) frequency described by the spatial Wright-Fisher model with *N*_*D*_ demes. The dynamics of the true and observed frequencies can be approximately described by the following Markov process (Fig. S6).

- *Transition probability distribution*: The true frequency vector 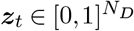 (hidden state) obeys

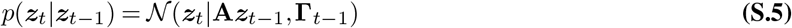

with the importation-rate matrix 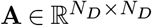 satisfying **A**_*ij*_ *>* 0 (*i*≠ *j*) and Σ_*j*_ **A**_*ij*_ = 1, and

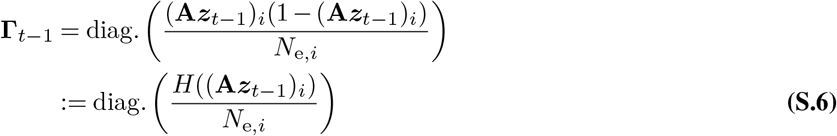 Here (**A*Z***)_*i*_ = Σ_*j*_ **A**_*ij*_*z*_*j*_, and *N*_e,*i*_ is the effective population size of deme *i*, which controls the strength of the genetic drift.
- *Emission probability distribution*: The observed frequency vector 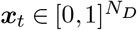 approximately obeys:

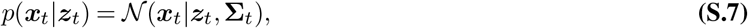

whereΣ_*t*_ characterizes the strength of the measurement error and is assumed to be given by

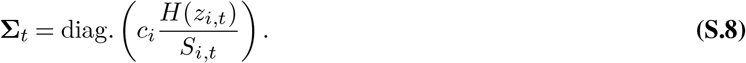 Here *S*_*i,t*_ is the number of sequences at week *t* in deme *i*, and *c*_*i*_ ≥ 1 is a parameter quantifying the deviation from ideal random sampling. The quantity *S*_*i,t*_*/c*_*i*_ can be interpreted as the *effective sampling size*.
- *Initial condition on z*_0_:

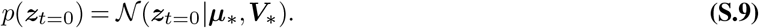 We assume that ***µ***_*_ and ***V***_*_ are given by

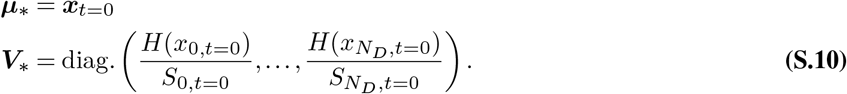

**Fig. S6.**
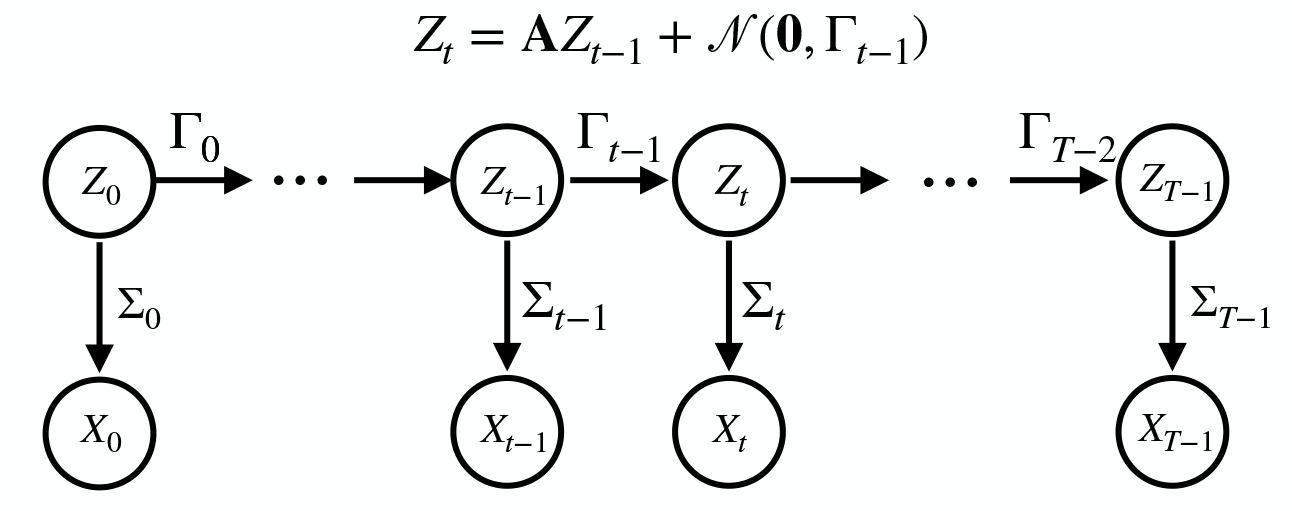
The HMM for the frequency dynamics. *Z*_*t*_ and *X*_*t*_ are true frequencies and observed frequencies, respectively. The transition probability densities (horizontal arrows) and emission probability densities (vertical arrows) are modeled by Gaussian distributions, Eqns. (S.5) and (S.7).

##### S.2.2 Inference

Given the observed frequencies, *x*_*i,t*_, and the number of sampled sequence, *S*_*i,t*_, our goal is to estimate the importation-rate matrix **A**, the effective population sizes *N*_e,*i*_, and the overdispersion of measurement noise *c*_*i*_. To achieve this, we developed two algorithms:

- Inference of the posterior distributions using a Markov Chain Monte Carlo (MCMC) method.
- Maximum likelihood estimation using an EM algorithm.

We employed the MCMC method for the region-level inference in England. Conversely, for deme-resolved matrices (Figs. 3A and 6A), we chose the EM method because of intensive computational requirements of the MCMC approach.

###### S.2.2.1 Computing the likelihood function and the filtered trajectories using the forward algorithm

Here, we describe the forward algorithm that computes the likelihood recursively. We have the identity:

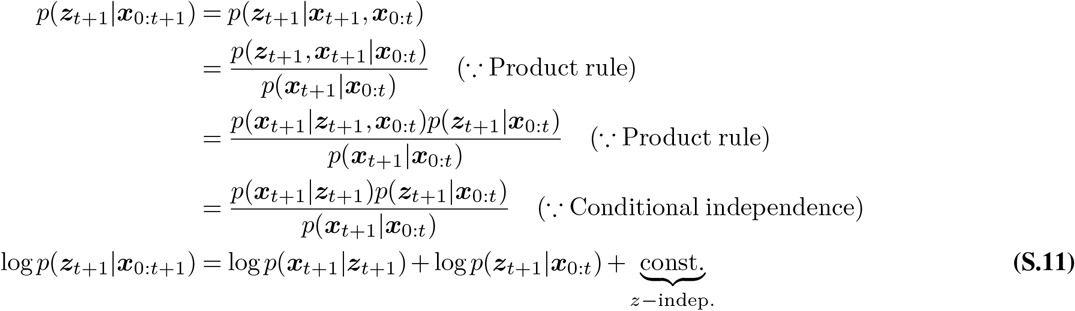

Because the second term *p*(***z***_*t*+1_|***x***_0:*t*_) can be rewritten as

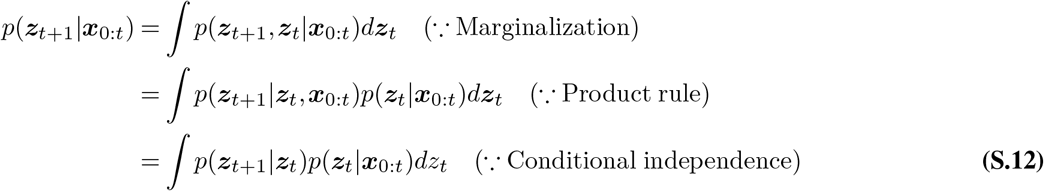

we have

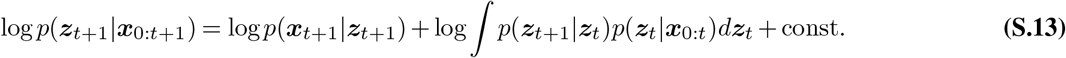

In the standard Kalman filter, a crucial step in deriving the recursion equations is performing Gaussian integration over the hidden state variables ***z***. In our model, the covariance matrices **Γ**_*t*_ and **Σ**_*t*_ are dependent on ***z***, which makes the process non-Gaussian. To utilize the technique of the Kalman filter, we approximate the covariances using matrices that are independent of ***z***;

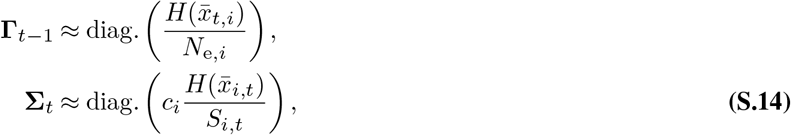

where the hidden frequency *z*_*i,t*_ is replaced by the time-averaged observed frequency 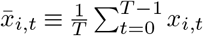. Under this approximation, all quantities appearing in Eq. (S.13) become Gaussian. We note that, unlike the standard Kalman filter, the covariance matrixΣ_*t*_ in our model is time-dependent due to the time-varying sampling rates *S*_*i,t*_. However, we can still perform Gaussian integrations even with the time-dependent covariance, as long as it remains independent of the hidden variables ***z***_*t*_.

After some calculations (as detailed in ref. (54)), it can be shown that the triplet {***µ***_*t*_, ***V***_*t*_, *l*_*t*_}, defined by

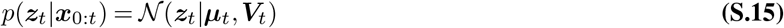

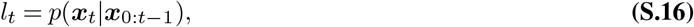

satisfies the following recursive equations:

###### Forward algorithm

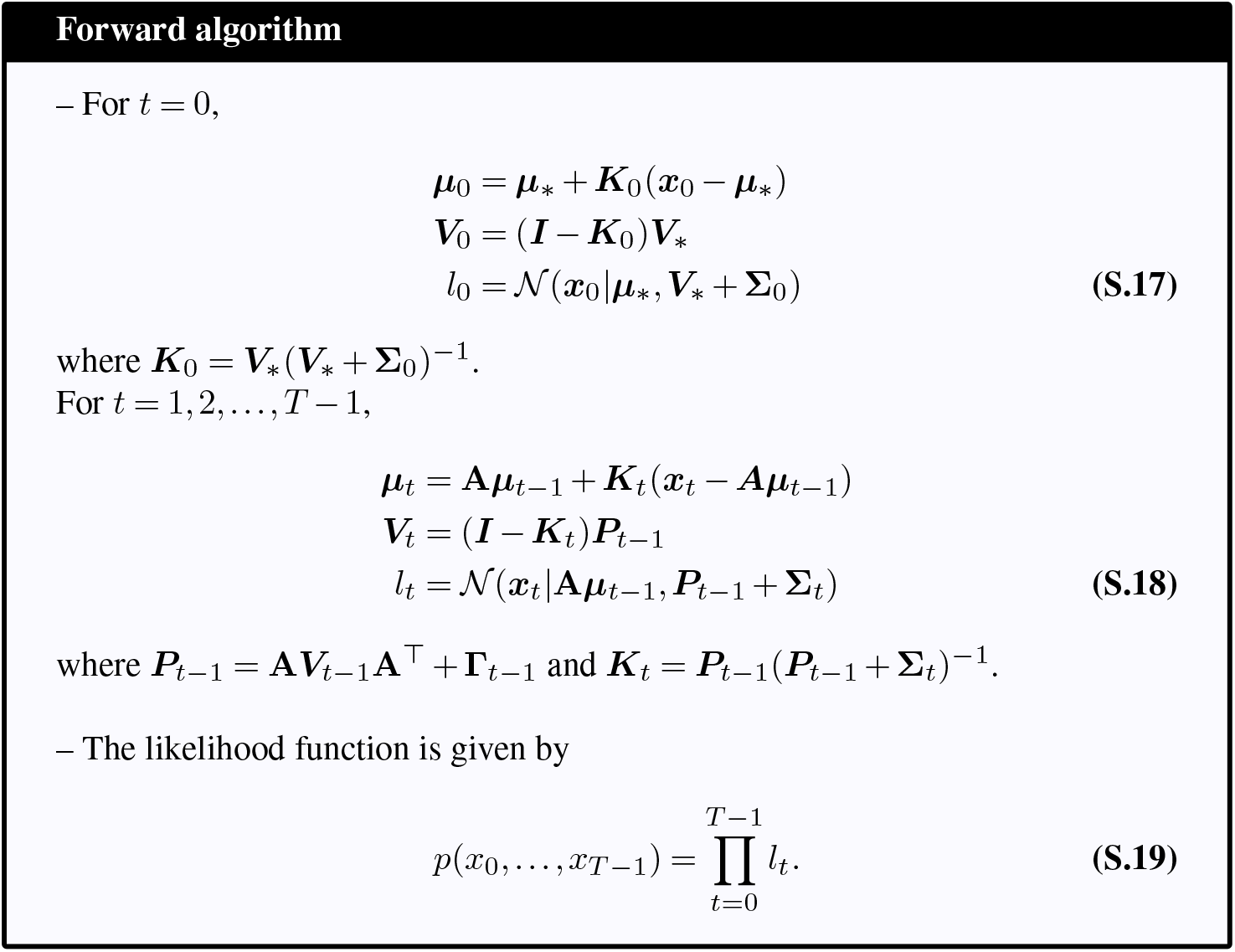

For each lineage (or allele) labeled by *ν*, we compute the triplet 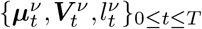 by solving Eqns. (S.17, S.18) recursively, with *µ*_*_ and *V*_*_ in Eq. (S.10) andΣ_*t*_ and Γ_*t*_ in Eq. (S.14). The total log likelihood of the time-series data for all lineages (or alleles) is given by

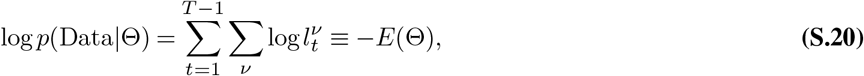

where Θ is the set of the parameters to be inferred, Θ = {**A**_*ij*_, *N*_e,*i*_, *c*_*i*_}.

We note that in Eq. (S.20), the log likelihood at the initial timepoint, 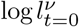, is excluded from the summation (even though *l*_0_ appears in the likelihood in Eq. (S.19)). This exclusion is appropriate due to our assumption about the true initial frequency, ***µ***_*_ = ***x***_0_. Under this assumption, Eq. (S.17) leads to 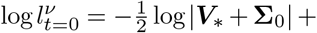 constant, which increases as the overdispersion parameter of measurement noise, *c*_*i*_, decreases (seeΣ_0_ in Eq. (S.14)), regardless of the values of measured frequencies. Therefore, including the term 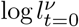 in Eq. (S.20) would introduce a bias that underestimates *c*_*i*_ when maximizing the likelihood.

###### S.2.2.2 Computing the posterior distributions using an MCMC

We consider the flat prior distributions *p*_prior_(Θ) on Θ. In this case, the posterior distribution is proportional to the likelihood,

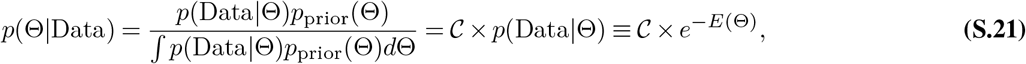

where 𝒞 is a factor independent of Θ.

To obtain the posterior distribution *p*(Θ| Data) ∝*e*^−*E*(Θ)^, we perform an MCMC method, namely, simulate a random walk in the Θ space whose stationary distribution is given by *p*(Θ|Data)∝ *e*^−*E*(Θ)^. Specifically, we implement the Metropolis algorithm:

###### MCMC (Metropolis algorithm)

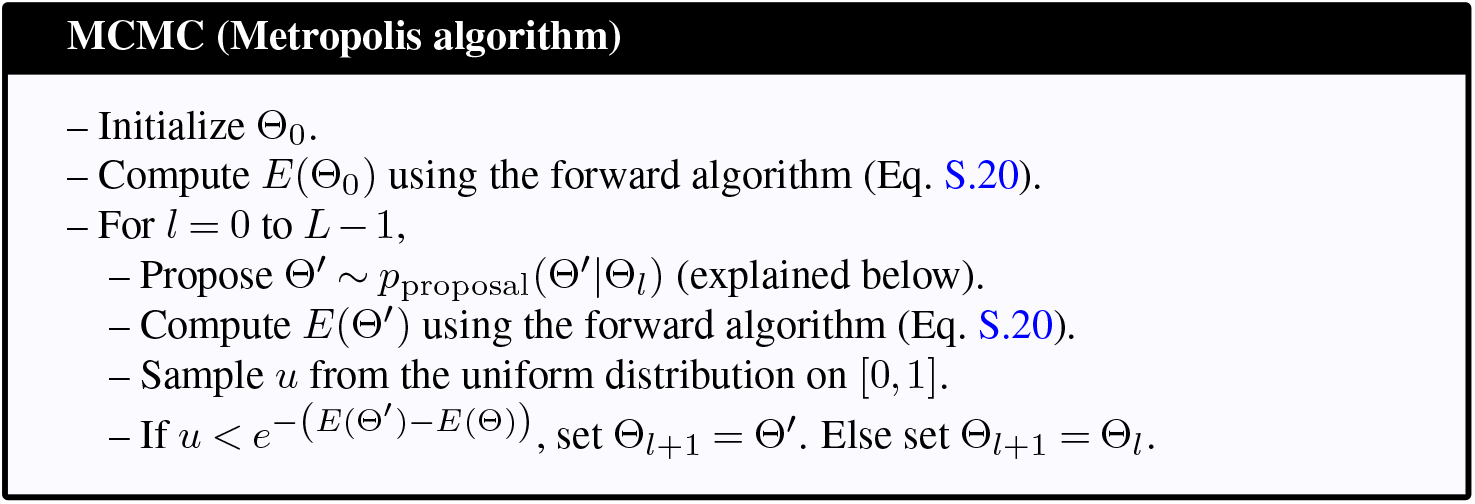

For sufficiently large Monte Calro steps *L* (e,g., *L* ∼ 10^6^), the sequence Θ_0_, Θ_1_, …, Θ_*L*_ approximates the posterior distribution *p*(Θ |Data).

For the proposal distribution *p*_proposal_, we employ the followings:

- For *N*_eff,i_, sample *h* ∼ 𝒩 (0, *ϵ*_*N*_), where *ϵ*_*N*_ is a positive constant. If *N*_eff,i_ + *h* ≥ 0, set 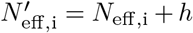. Else set 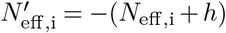.
- For *c*_*i*_ ∈ Θ, sample *h* ∼ 𝒩 (0, *ϵ*_*c*_), where *ϵ*_*c*_ is a positive constant. If *c*_*i*_ + *h* ≥ 1, set 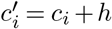. Else set 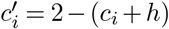.
- For the importation-rate matrix **A**_*ij*_, randomly choose a pair (*i, j*) and sample *ϵ* ∈ [−**A**_*ii*_, **A**_*ij*_] from the uniform distribution. Set 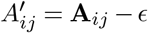 and 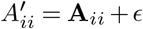.

*Remark:* While one may use other forms of *p*_proposal_(Θ^*′*^|Θ), it must satisfy two critical conditions:

1. The proposed Θ^*′*^ must be within the meaningful parameter region. For example, 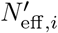 must be positive, and 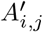 must lie on the simplex.
2. *p*_proposal_(Θ^*′*^| Θ) must be symmetric, i.e., *p*_proposal_(Θ^*′*^| Θ) = *p*_proposal_(Θ|Θ^*′*^). Otherwise, the Metropolis-Hastings algorithm should be used instead of the Metropolis algorithm.

Note that in our proposal distribution for *N*_e,*i*_ and *c*_*i*_, any proposed value that exceeds the boundary of the parameter space is reflected along the boundary. This prescription ensures the symmetric condition near the boundary. For **A**_*ij*_, our proposal distribution also satisfies the above two conditions (see ref. (55)).

##### S.2.3 Computing the maximum likelihood estimation through an EM algorithm

We use an EM algorithm to search for the maximum likelihood estimate of the parameters Θ. Suppose that the model parameter is Θ_old_ = (**A, *N***_e_)_old_ at a stage of the algorithm. By using Θ^old^, for each lineage, we first compute the filtered trajectory ***µ***_*t*_ and the covariance ***V***_*t*_ using the forward algorithm. Then, we compute the smoothed trajectory 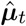 and the variance 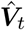 by solving the backward equation:

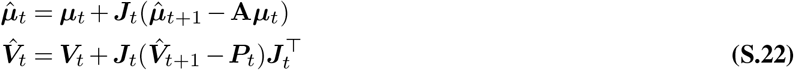

where ***P***_*t*_ = **A*V***_*t*_**A**^⊤^ + **Γ**_*t*_ and ***J***_*t*_ = ***V***_*t*_**A**^⊤^(***P***_*t*_)^−1^. Solving the backward algorithm provides the following quantities, which will be required below^1^;

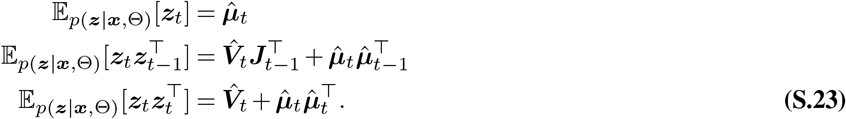

In the EM algorithm, we need to minimize the expectation *Q* of the complete-data likelihood *p*(***z, x***, Θ) (Here, ***z*** and ***x*** collectively denote the frequencies ***z***^*ν*^ and ***x***^*ν*^ of all *N*_lin_ lineages). It can be shown that the terms of *Q* that are dependent on **A**_*ij*_ and *N*_e,*i*_ are given by (see ref. (54))

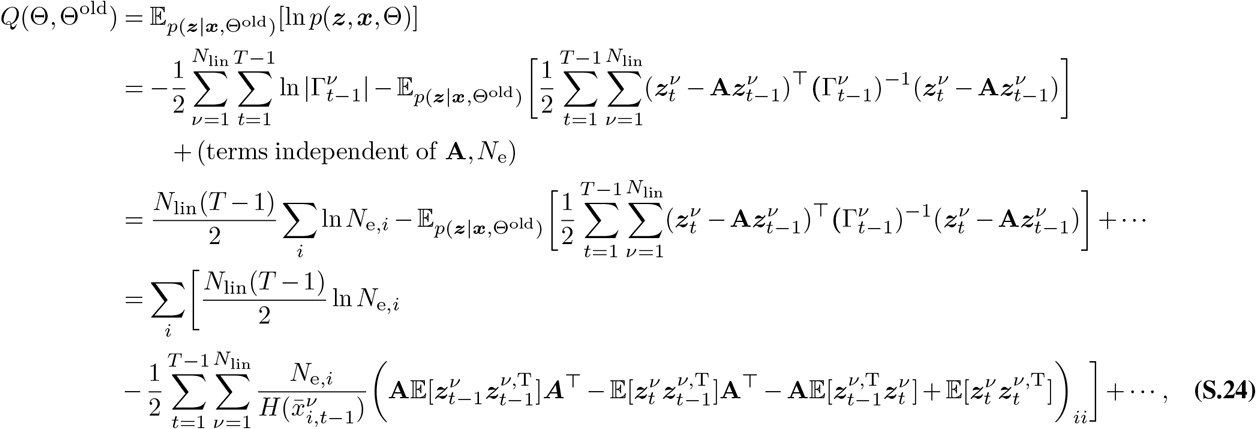

where (…)_*ii*_ in the last line represents the *ii* component of the matrix inside the parentheses. The notation |*M*| denotes the determinant of a matrix *M*.

The new matrix **A** is obtained by minimizing the quantities in the second line of Eq. (S.24). Since **A**_*ij*_ and 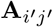 are decoupled if *i*≠ *i*^*′*^, each row of **A**_new_ can be determined separately. Specifically, the *i*-th row 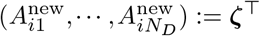 is obtained by solving

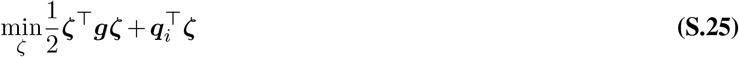

under the constraints, Σ_*j*_ *ζ*_*j*_ = 1 and *ζ*_*j*_ *>* 0. The matrix ***g*** and the vector ***q***_*i*_ are given by

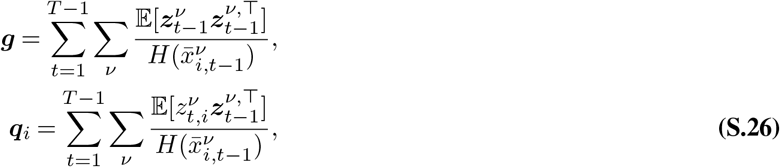

which can be evaluated by using Eq. (S.23). We determine each row of **A**^new^ by solving the constrained quadratic programming Eq. S.25 using the Python package CVXOPT.

After determining **A**_new_, the new effective population size 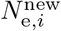 is determined by differentiating Eq. (S.24) with respect to *N*_e,*i*_. The result is

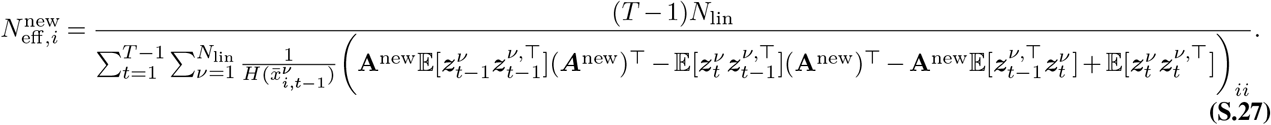

*Q* also has the terms that are dependent on the parameter describing the deviation from uniform sampling, *c*_*i*_;

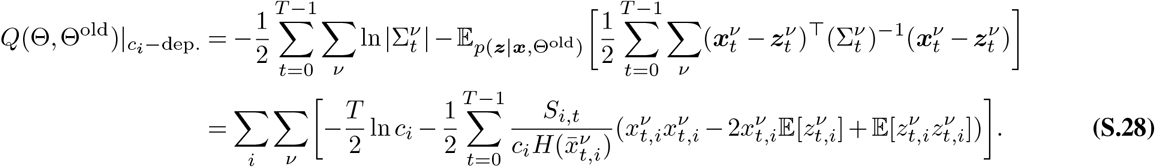

The new value of the measurement noise overdispersion *c*^new^ is determined by minimizing the above expression with respect to *c*_*i*_ (subjected to ≥ 1):

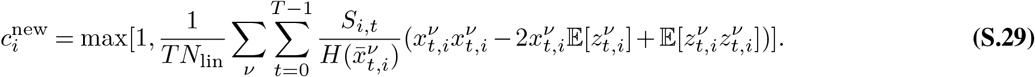

In the EM algorithm, we initialized the parameters, Θ = Θ_0_, and iteratively updated them to Θ_new_, by solving Eq. S.25 and evaluating Eqns. S.27 and S.29, until the likelihood stabilizes/converges.

###### Regularization

To stabilize the inference for deme-resolved analysis (higher than at region level), we added a Ridge-like regularization term 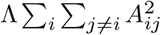, to *Q*. Here, Λ is a regularization parameter. The value of Λ is determined via cross-validation by dividing the set of allele-frequency (or lineage-frequency) trajectories into training and test data of equal size.

###### Bootstrapping

To evaluate the uncertainty in the MLE, we performed bootstrapping by randomly sampling lineages with replacement to generate multiple new sets, each containing the same number of lineages as the original dataset. We then applied the EM algorithm to each of these generated sets.

##### S.2.4 Computational tests

- 3-deme system We simulated frequency time-series data using the 3 × 3 matrix shown in Fig. 2. The effective population size and sampling rate were set to values similar to those inferred for London and its neighboring regions during the Delta wave. The results for the simulated data, inferred using the HMM-MCMC, HMM-EM methods, and the least squares (LS) method (see Eq. 4), are compared in Figs. S7A-D. The LS method, which ignores fluctuations due to genetic drift and sampling noise, tends to overestimate the interaction strengths (Fig. S7C). Note that the results from the LS method are not displayed in Figs. S7C and D because the LS method cannot infer *N*_e,*i*_ and *c*_*i*_.
- 50-deme system We simulated frequency time-series data using the 50 × 50 matrix shown in Fig. S8A, which was constructed by assuming five strongly interacting blocks, each consisting of 10 demes. The effective population size *N*_e,*i*_ and *c*_*i*_ were set to *N*_e,*i*_ = 1200 and *c*_*i*_ = 1.0, respectively. The sampling rate was assumed to be *S*_*i*_ = 500 for all demes. These values were chosen to mimic the actual situation during the Delta wave in England. From the simulated data, we inferred the parameters using the LS and HMM-EM methods. Fig. S8B compares the matrix elements of the true **A**_*ij*_ with those of the inferred **A**_*ij*_. The LS method tends to overestimate interactions, especially for small couplings. This bias is further illustrated in Fig. S8C, which shows the histogram of the inferred matrix elements corresponding to the zero elements in the true matrix.

The bias in the LS method can be intuitively understood by considering that in a large noise limit, the LS solution converges to a homogeneous matrix 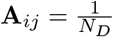. Thus, using the LS method, small couplings (roughly,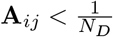) tend to be overestimated, while large couplings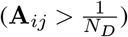 tend to be underestimated.

**Fig. S7.**
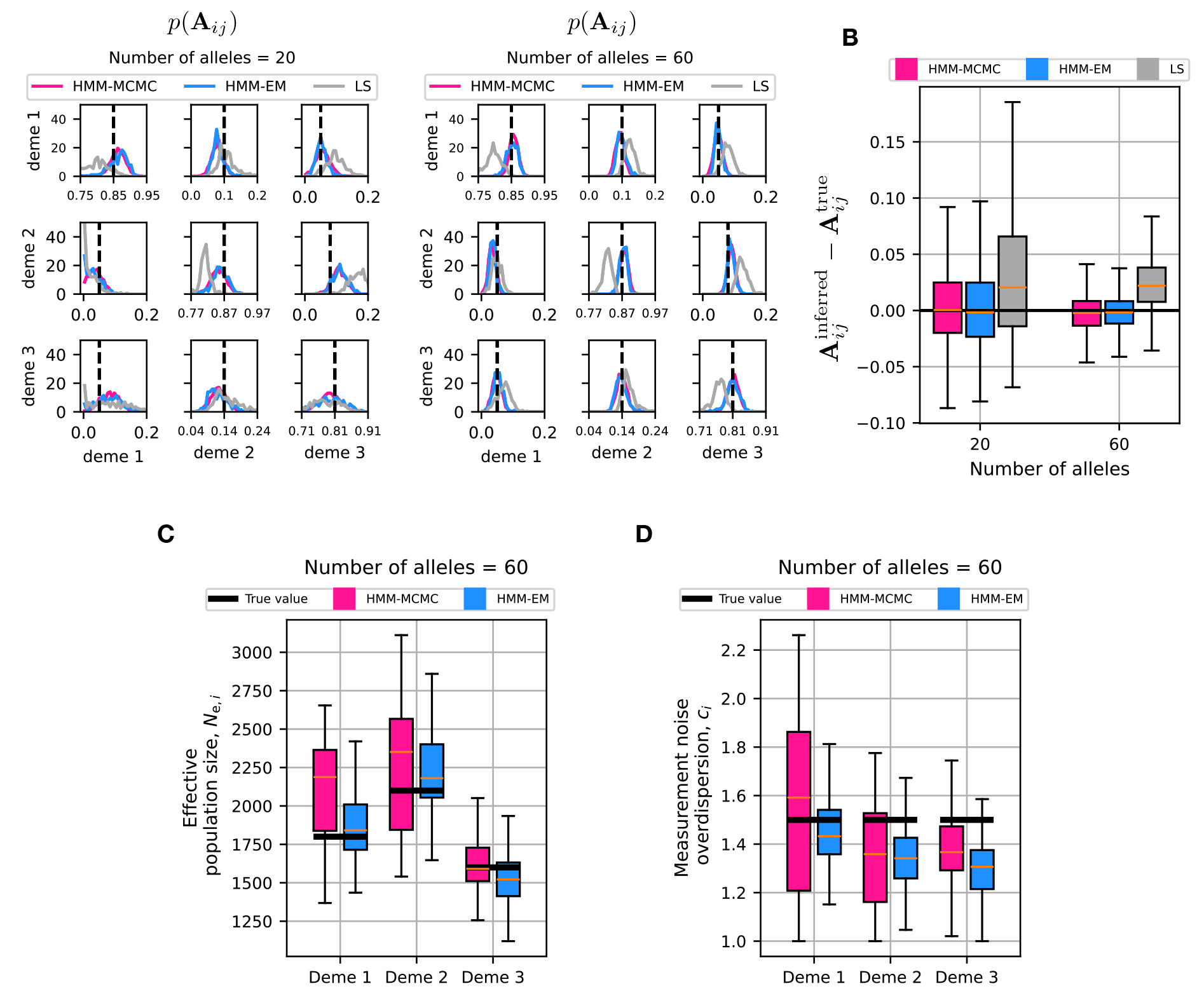
Supplementary figures to Fig. 2. The Wright-Fisher model on three demes is simulated, where the importation-rate matrix **A** and the effective population sizes are those shown in Fig. 2A. The measurement noise overdispersion parameter is set to *c*_*i*_ = 1.5. (**A**) The distribution of **A** inferred by the HMM-MCMC, HMM-EM, and LS methods, from 20 alleles (left) and 60 alleles (right). For the HMM-MCMC method, it represents the posterior distribution, while for the HMM-EM and LS methods, it represents the bootstrap distribution. (**B**) The error, 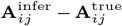, in the inference of interaction strengths for the HMM-MCMC, HMM-EM, and LS methods, where all pairs *i, j* are aggregated. (**C**) Boxplots showing the effective population sizes, inferred using the HMM-MCMC method from 60 alleles. The true values are indicated by thick horizontal lines. (**D**) Boxplots showing the measurement noise overdispersion, *c*_*i*_, inferred using the HMM-MCMC method from 60 alleles. The true values (*c*_*i*_ = 1.5) are indicated by thick horizontal lines.

**Fig. S8.**
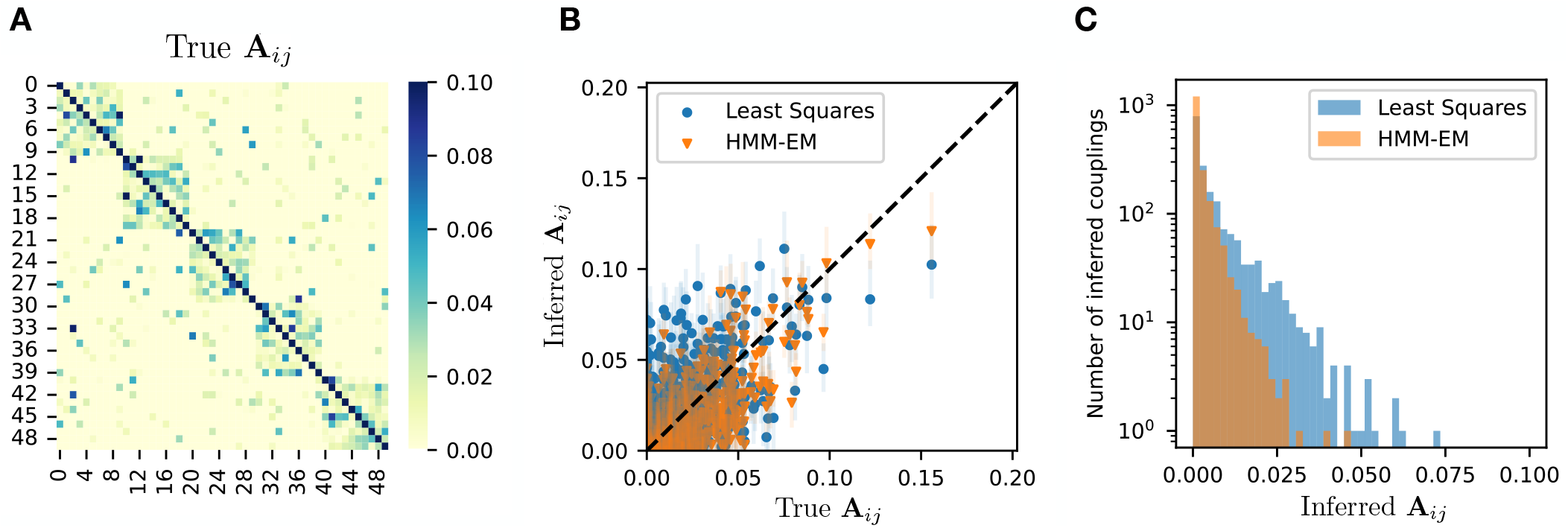
(**A**) Heatmap showing the true matrix. All matrix elements within the five diagonal blocks (each consisting of 10 demes) are assumed to be nonzero, while the elements of the off-diagonal blocks are nonzero with probability of 0.1. The nonzero elements are randomly sampled from the exponential distribution with a mean of 0.02. **B**. Element-wise comparison between the true matrix and the inferred matrix obtained using the least squares and HMM-EM methods. (**C**) Histogram of the inferred matrix elements corresponding to zeros in the true matrix.

#### S.3 Reproductive values in England

In the main text, we have shown that the per-capita reproductive values *π*_*i*_*/I*_*i*_ vary across regions (Fig. 7C). Additionally, the unnormalized “class” reproductive values *π*_*i*_ are spatially heterogeneous, see Fig. S9.

##### S.3.1 Ranking regions according to ratios of bidirectional importation rates

As discussed in the main text, reproductive values predict ratios of bidirectional importation rates when the principle of detailed balance, Eq. 2, holds. To verify this, we simultaneously reordered both rows and columns of the matrix **A**_*ij*_ to maximize the asymmetry, as measured by Σ*i > j***A**_*ij*_*/*(**A**_*ij*_ + **A**_*ji*_). This approach indeed arranges regions roughly in line with the values of *π*_*i*_ for both the Alpha and Delta waves, see Fig. S10. In particular, the two regions, EE and SW, which have lower *π*_*i*_ values, are ordered last in the reordered matrices.

**Fig. S9.**
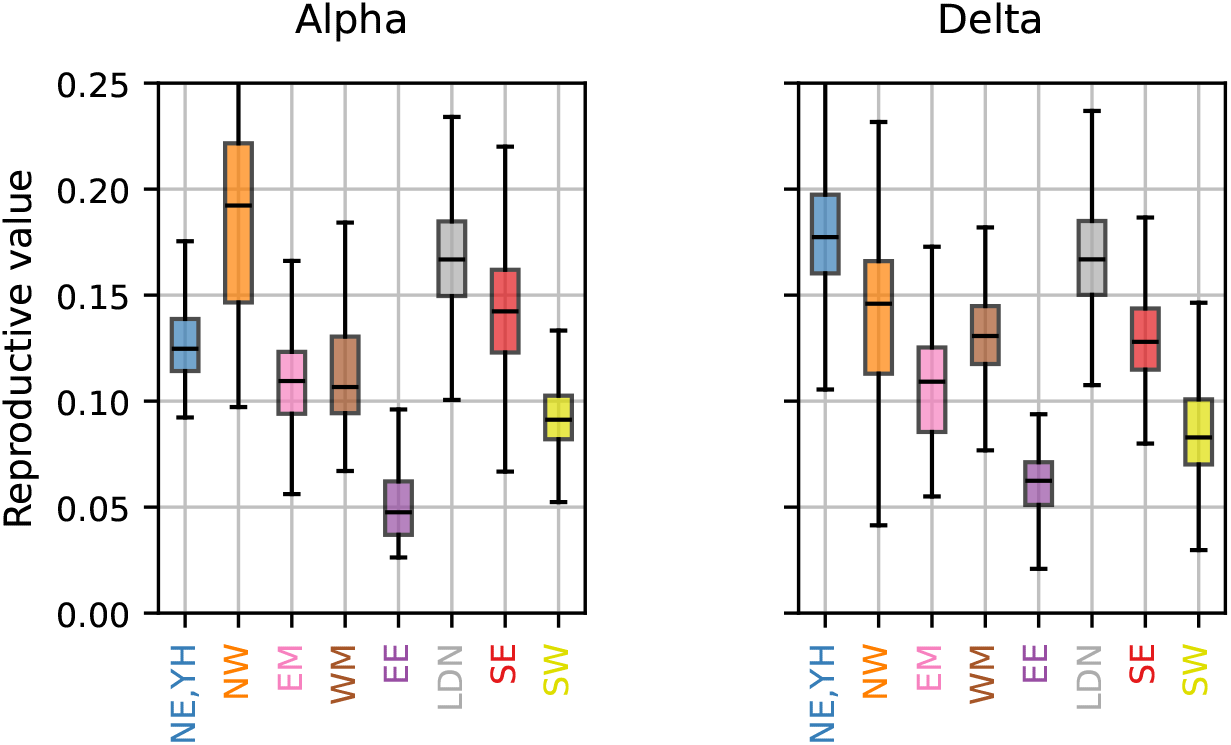
The inferred class reproductive values for each region during the Alpha wave (epiweeks 57-68) and Delta wave (epiweeks 82-95) in England.

**Fig. S10.**
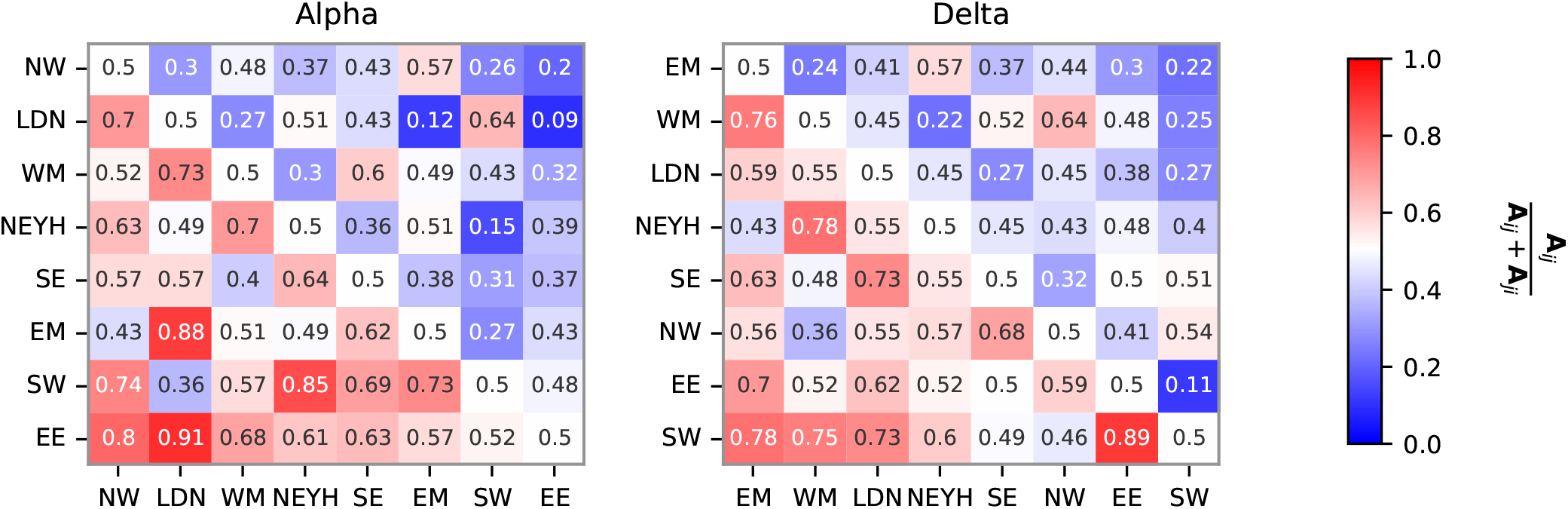
Using the inferred importation-rate matrix **A** for the Alpha and Delta variants in England and the number of infected *NI*_*i*_, as measured by the COVID-19 Infection Survey (37), the values of **A**_*ij*_ */*(**A**_*ij*_ + **A**_*ji*_) are represented by heat maps. The order of regions is arranged to maximize the asymmetry, Σ_*i>j*_**A**_*ij*_ */*(**A**_*ij*_ + **A**_*ji*_), is maximized.

##### S.3.2 Testing the effect of the delay in sequence reporting

One of the possible reasons for the observed heterogeneity in the reproductive value *π*_*i*_ is the difference in the population size. In fact, *π*_*i*_ correlates with the average number of infected individuals in each region during the wave; the Spearman correlation coefficients are *ρ* = 0.81 (p-value 0.015) for the Alpha wave and *ρ* = 0.88 (p-value 0.004) for the Delta wave, respectively. However, as presented in Fig. 7, the reproductive values are heterogeneous even after normalizing by the number of infected individuals.

Another possible explanation could be that the inferred heterogeneity in *π*_*i*_ results from artifacts caused by variations in the timing of disease reporting. For instance, assume two regions *i* and *j* have perfectly symmetrical cross-importation rates.

If region *i* reports cases substantially later than region *j*, its frequency trajectories will tend to follow region *j*, mimicking a causal influence of *j* on *i*. Consequently, the importation rate from *j* to *i*, **A**_*ij*_, is likely to be overestimated, which would then expected to decrease *π*_*i*_. A more quantitative argument based on perturbative analysis is provided in Sec. S.3.3.

While no systematic reporting time heterogeneity has been documented to our knowledge, to test the impact of a hypothetical reporting delay, we artificially advanced the allele counts data from EE by *τ*_shift_(= 1, 2) weeks compared to the other regions, mimicking the scenario where the other regions report sequences later by *τ*_shift_ weeks. We then re-inferred the importation-rate matrix (Fig. S11). As expected, the reproductive value of EE increases as *τ*_shift_ is increased, highlighting the importance of accurate reporting dates for precise inference.

**Fig. S11.**
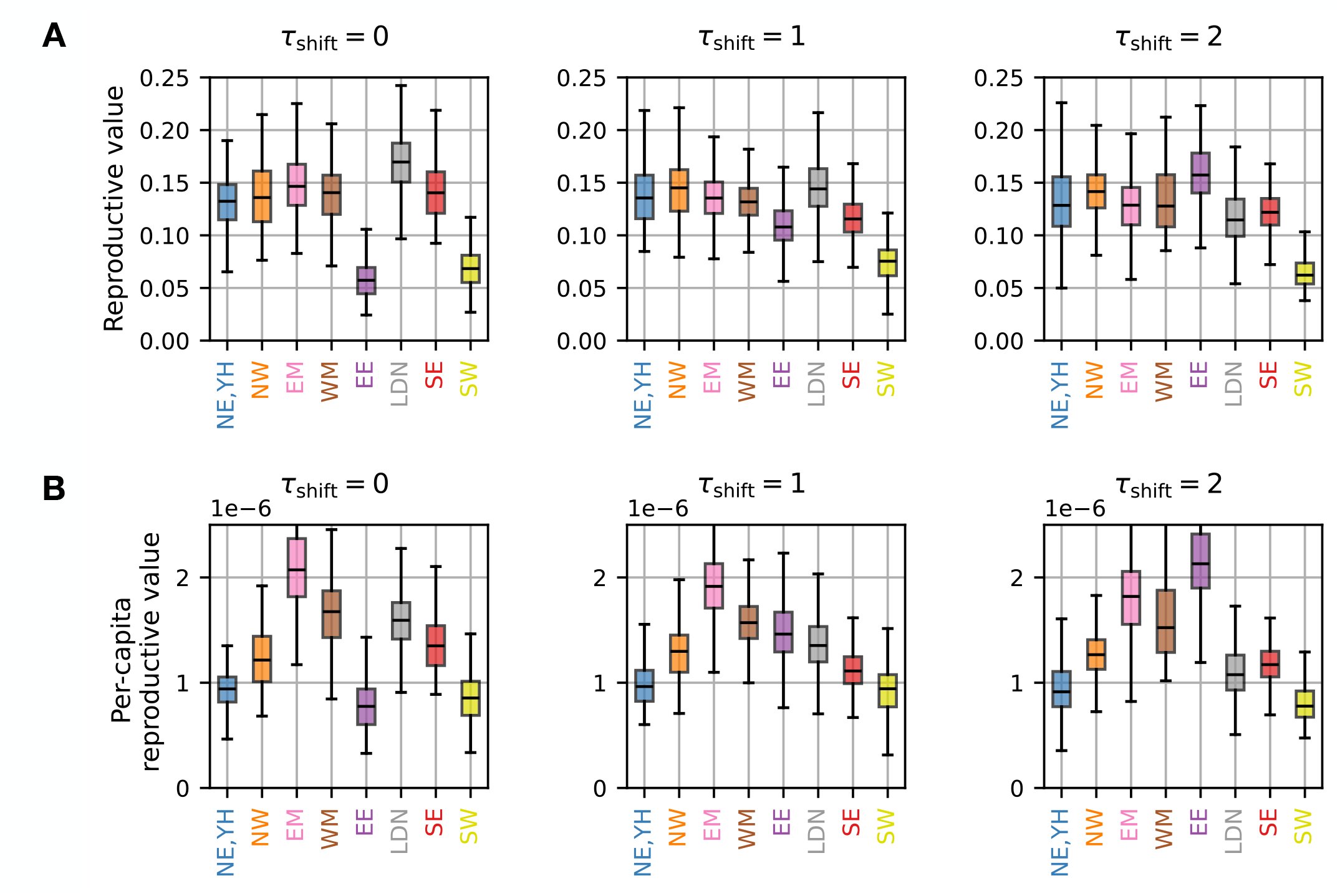
(**A**) The reproductive values inferred for the data during the Delta wave in England (epiweek 82+*τ*_shift_ to 93+*τ*_shift_ for EE and epiweek 84 to 94 for the other regions), where sequence reporting dates from EE are artificially advanced by *τ*_shift_ weeks. (**B**) The inferred per-capita reproductive values.

##### S.3.3 Perturbative analysis of the effect of delay in reporting

Consider a population consisting of two groups, each containing 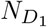 and 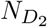 demes. Assume a consistent delay of one week in sequence reporting from the demes in the second group. Ignoring the genetic drift and measurement noise, the dynamics of the allele frequencies, 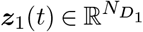 in the 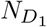 demes and 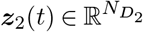 in the 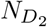 demes, are described by

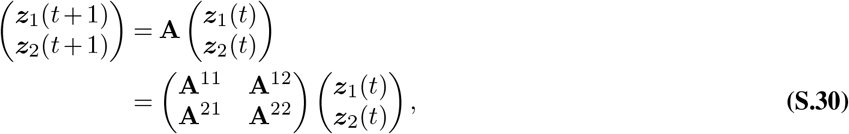

where **A**^*IJ*^ (*I, J* = 1, 2) represents the matrix of importation rates from demes in *J* to demes in *I*. The observed frequencies, 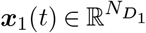 and 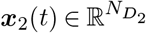, are given by

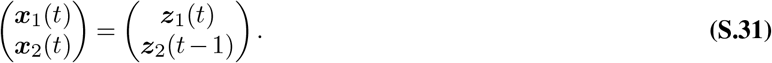

The dynamics of the observed frequencies are written as

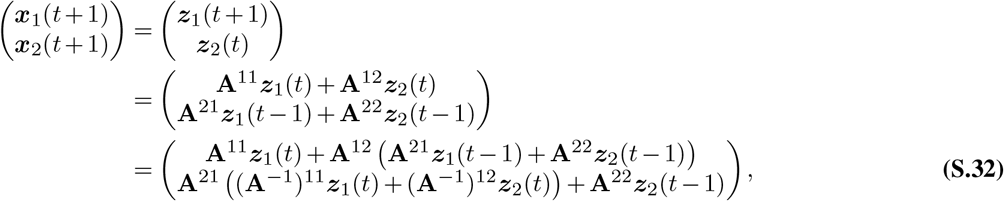

where 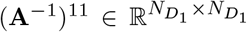 and 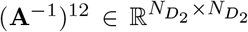 are the block matrices of **A**^−1^, defined by 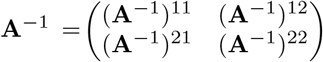. We assume that the interactions are weak, i.e. **A** = 𝒪 (*ϵ*) (ϵ ≪ 1) for *i* ≠ *j*, which implies ***z***_1_(*t* − 1) = ***z***_1_(*t*) +𝒪 (*ϵ*) and ***z***_2_(*t*) = ***z***_2_(*t* − 1) +O(*ϵ*). Under this approximation, Eq. (S.32) can be written as

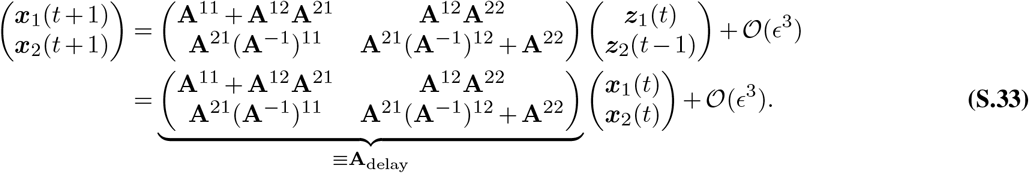

Hence, under the delay from the second group, the apparent dynamics are still linear but described by the modified matrix **A**_delay_. Compared to the true matrix 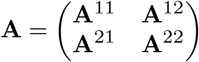, the delay fictitiously modifies the upper-right block (representing the influence of the second group on the first group) by introducing the factor **A**_22_ and the lower-left block (representing the influence of the first group on the second group) by introducing the factor (**A**^−1^)^11^.

To illustrate the delay effect, let us consider the simplest case where 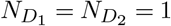, where **A**^11^, **A**^12^, **A**^21^, **A**^22^ are not matrices but positive scalars. The constraints **A**^11^ + **A**^12^ = 1 and **A**^21^ + **A**^22^ = 1, along with the positivity condition, imply that the delay-induced factors satisfy **A**^22^ *<* 1 and (**A**^−1^)^11^ *>* 1. Therefore, when the second group’s report is delayed, the influence of the second group on the first group becomes underestimated, whereas the influence of the first group on the second group becomes overestimated. This effect is demonstrated for cases where 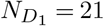 and 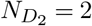 in Fig. S12.

**Fig. S12.**
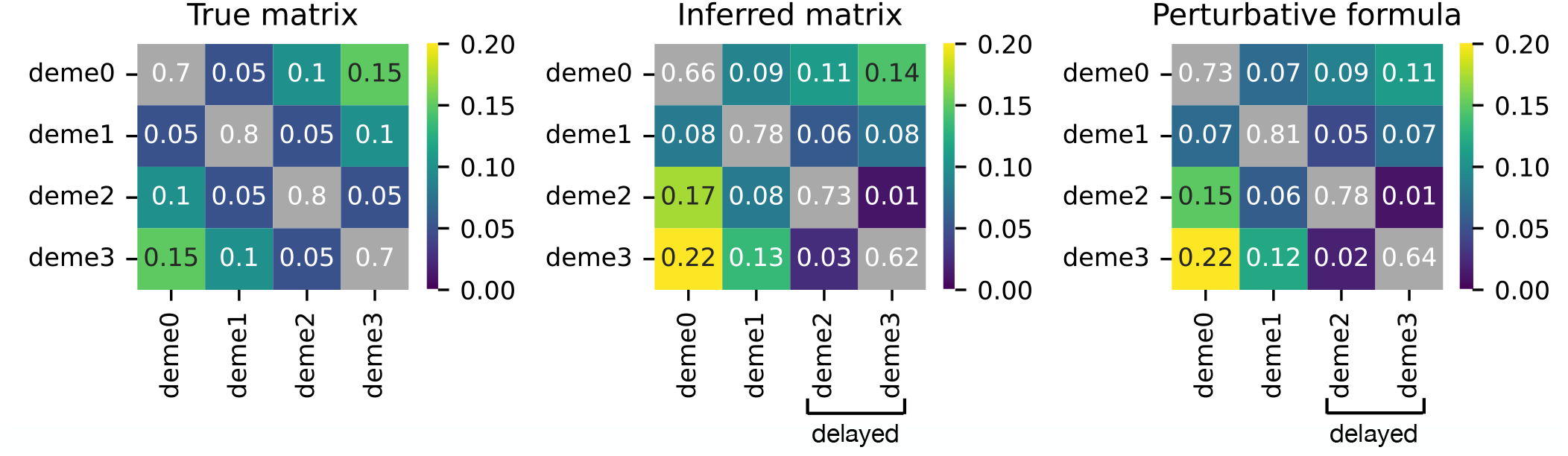
The Wright-Fisher simulation was performed with the 4*×* 4 importation-rate matrix shown in the leftmost heatmap. The effective population sizes and sampling rates were set to *N*_e,*i*_ = 10^4^ and *S*_*i*_ = 10^4^, respectively, for all demes. We examined a scenario in which there was a 1-week delay in sequence counts from deme 2 and deme 3. The matrices inferred from the HMM method and the perturbative formula in Eq. S.33 are presented in the middle and rightmost heatmaps, respectively. Consistent with the formula, the influences from demes 2 and 3 (corresponding to the third and fourth columns) are underestimated due to the delay, while those from demes 1 and 2 (corresponding to the first and second columns) are overestimated.

#### S.4 Spatial Resolution and Inference Reliability

In the deme-resolved results presented in Fig. 3, we divided England into 50 demes. In this section, we present two arguments supporting the idea that increasing the spatial resolution much beyond 50 demes is not feasible.

The first argument considers how **A**_*ij*_ should behave under changes in spatial resolution. Let *i* and *j* represent demes, and let *I* and *J* represent coarse-grained areas, referred to as regions. Suppose we have a deme-level matrix **A**_*ij*_. If all demes within the same region interact strongly, the region-level interaction **A**_*IJ*_ can be approximated by:

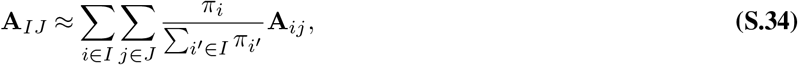

where *π*_*i*_ is the reproductive value(the normalized left eigenvector with eigenvalue 1 of the matrix **A**_*ij*_). This formula can be understood from a time-backward perspective: assuming demes within the same region equilibrate instantaneously, a lineage in region *I* is distributed across demes *i* ∈ region *I* with probability 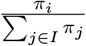. The lineage in deme *i* ∈ region *I* then moves backward in time to deme *j* with probability **A**_*ij*_. Summing over demes *i* ∈ region *I* and *j*∈ region *J* gives the probability that a lineage in region *I* moves backward in time to region *J*, which corresponds to **A**_*IJ*_ in Eq. (S.34).

In Fig. S13A and B, we show the deme-resolved matrix with *N*_*D*_ = 50 for the Delta wave in England and its coarse-grained counterpart. As shown in Fig. S13C, the coarse-grained matrix closely matches the one directly inferred from the region-level allele frequency data. Fig. S13D shows the correlation between the two matrices as a function of the number of demes *N*_*D*_. The decline in correlation above *N*_*D*_ ≈ 70 indicates a loss of inference reliability at higher spatial resolutions.

**Fig. S13.**
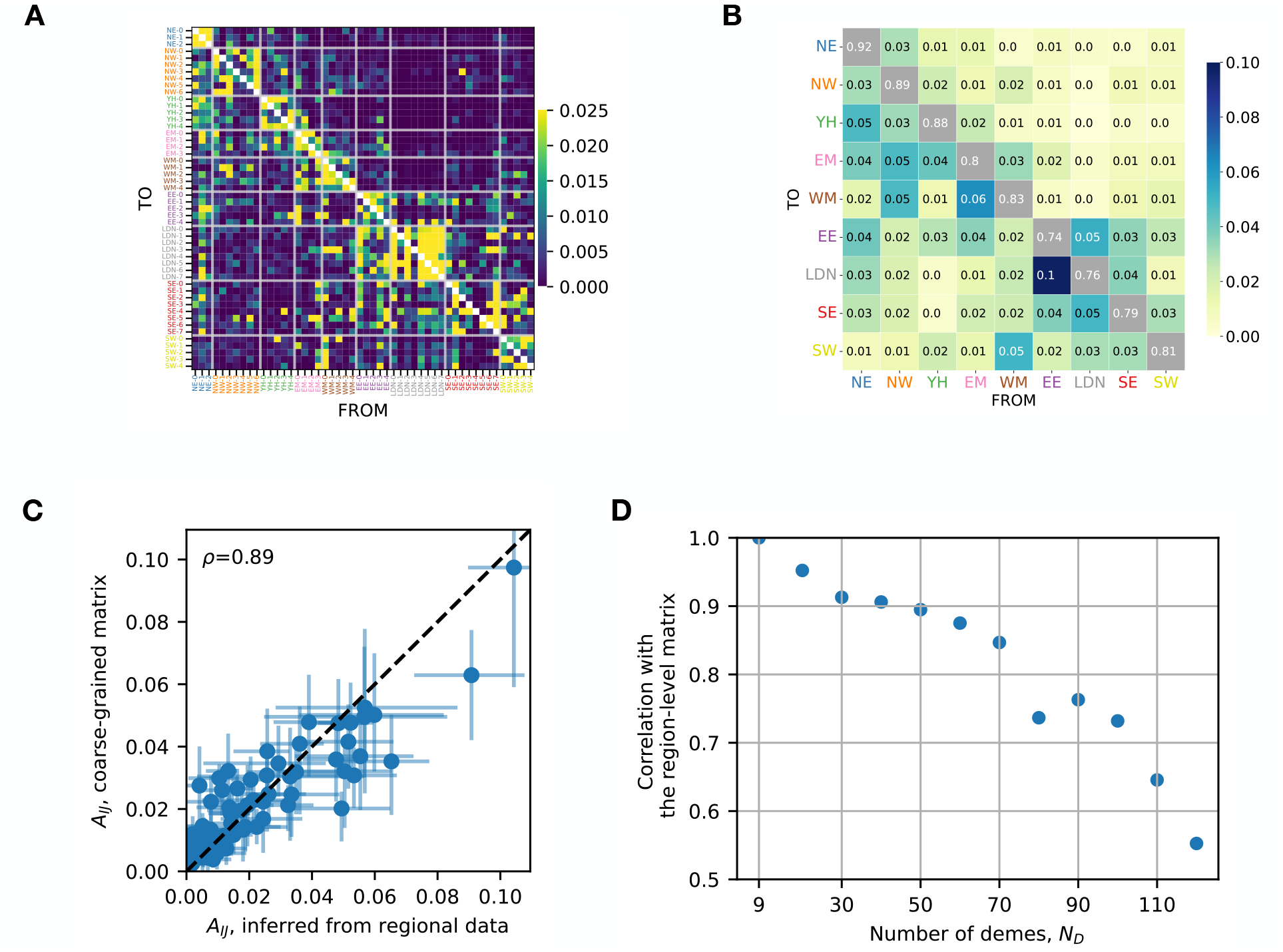
(**A**) Heatmap showing the importation-rate matrix **A**, with *N*_*D*_ = 50, during the Delta wave in England.(**B**) The coarse-grained matrix obtained by applying Eq. Eq. (S.34) to the 50 *×* 50 matrix in Fig. **A**. (**C**) Element-wise comparison between the coarse-grained matrix and the matrix directly inferred from the region-level mutation data. The Pearson correlation coefficient is 0.89. (**D**) Plot showing the Pearson correlation between the 9 *×* 9 coarse-grained matrix, obtained from *N*_*D*_ *×N*_*D*_ deme-resolved matrix, and the 9 *×* 9 matrix directly inferred from the region-level mutation data, as a function of the number of demes *N*_*D*_. For each spatial resolution *N*_*D*_, the matrix elements between the two matrices are compared as in Fig. **C**.

Another indirect argument comes from the relaxation times of the eigenmodes. Fig. S14A displays the inferred importation-rate matrix for the Delta wave at the level of upper tier local authorities (UTLA), which is the finest spatial resolution available from our data. Fig. S14B shows the relaxation time, ln −|*λ*_*i*_|, for each eigenmode (| *λ*_1_| ≤ |*λ*_2_ | ≤ …). While the relaxation time around *i* = 50 is approximately 2 weeks, it decreases to less than 1 week for *i >* 60. Given that the unit time of our analysis is 1 week, modes with relaxation times shorter than 1 week cannot be reliably inferred. This observation suggests that significantly increasing the spatial resolution beyond 50 demes is not practical.

#### S.5 Epidemiological interpretation of A

To aid in interpreting our importation-rate matrix **A**, we derive its expressions using specific epidemiological models.

**Fig. S14.**
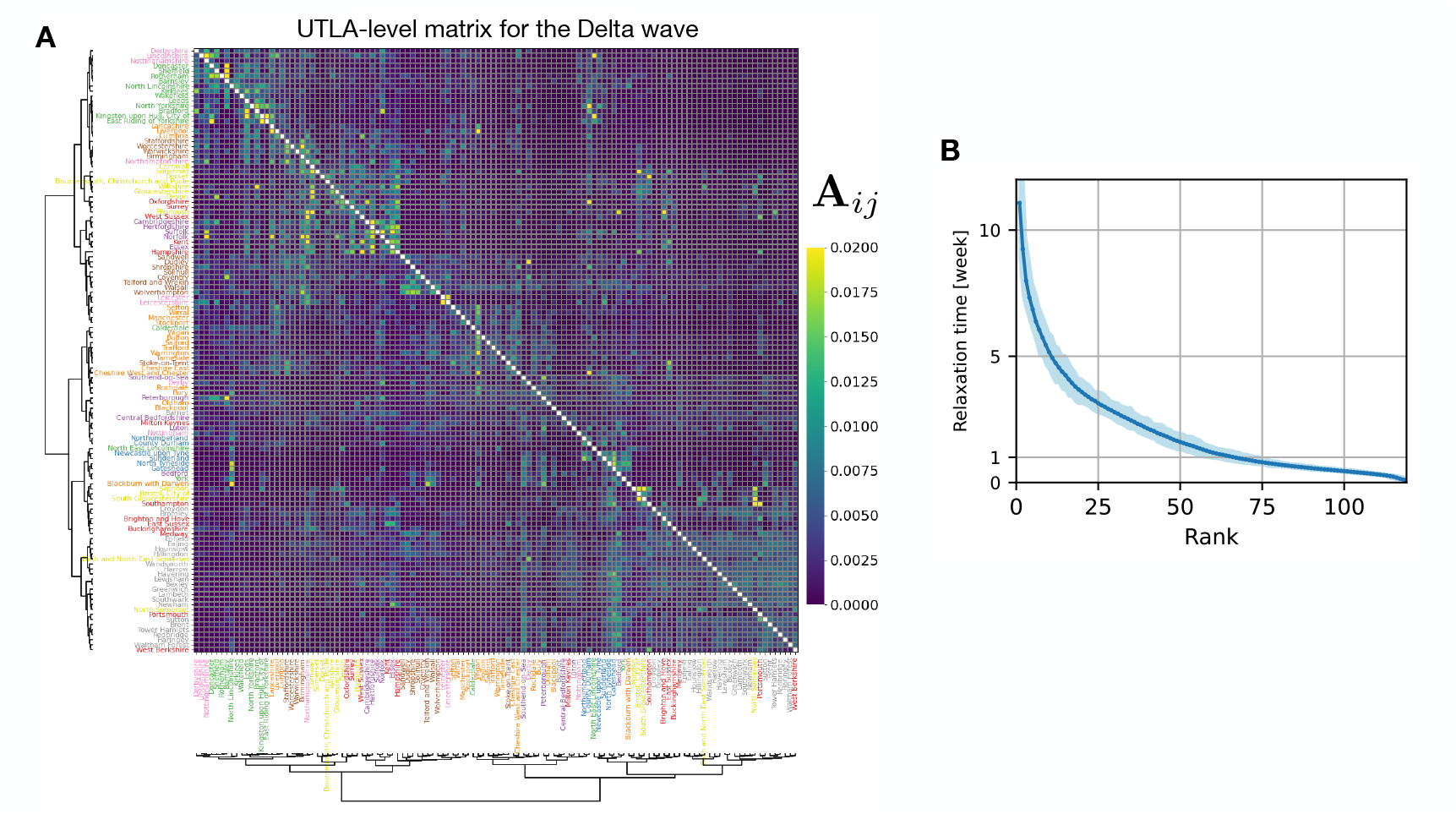
(**A**) The importation-rate matrix among 120 UTLAs in England, inferred for the Delta wave (Jun 20, 2021 - Sep 25, 2021) with the HMM-EM method (without Ridge regularization). The top 120 UTLAs with the most sequences are used in this inference. The 120 UTLAs are ordered by performing hierarchical clustering with the Jensen-Shannon distance between rows. (**B**) The relaxation time for each mode of the importation-rate matrix. The shaded region shows the 95% confidence interval, computed using the bootstrapping method.

We begin by extending the multi-patch SIR model (11, 53, 56, 57) to include distinct multiple viral lineages. The extended model is represented by the following set of equations:

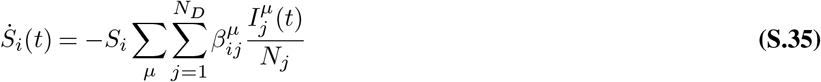

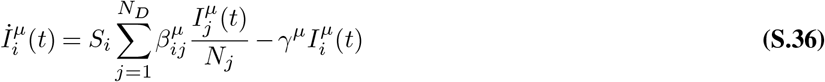

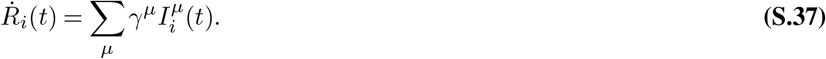

Here, 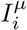 denotes the number of individuals infected by lineage *µ* in deme *i, N*_*i*_ is the population size, *N*_*i*_ = *S*_*i*_(*t*)+*I*_*i*_(*t*)+*R*_*i*_(*t*), and 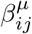 is the transmission rate of lineage *µ* from an infected individual in deme *j* to susceptible individuals in deme *i. γ*^*µ*^ represents a lineage-specific recovery rate.

In terms of the total number of infected individuals 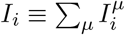 in deme *i* and the frequency 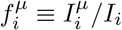 of lineage *µ*, Eq. (S.36) is expressed as

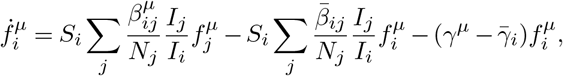

where 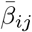 and 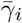 are lineage-averaged rates defined as

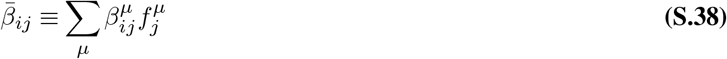

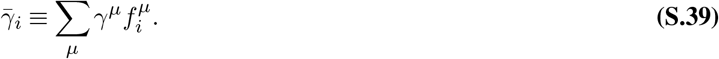

In terms of the fractions 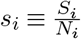 and 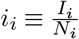, we have

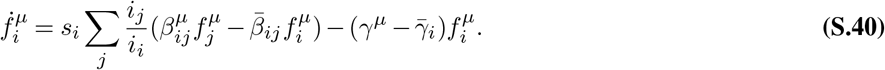

##### S.5.1 Neutral case

Suppose that all lineages have the same transmission rate and recovery rate, 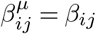 and *γ*^*µ*^ = *γ*. Then, Eq. (S.40) reduces to

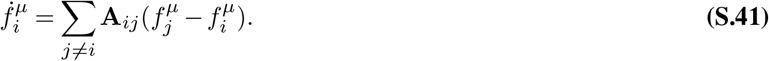

Where

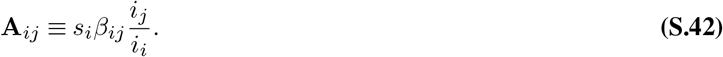

If we assume that *s*_*i*_ ≈1, which is a reasonable approximation in realistic situations, and that the infected fraction is spatially homogeneous (*i*_*i*_ ≈ *i*_*j*_), as was the case during the plateau period of the Delta wave in England (Fig. S26), then **A**_*ij*_ can be approximated as **A**_*ij*_ ≈ *β*_*ij*_.

By discretizing time using one week as the unit of time,

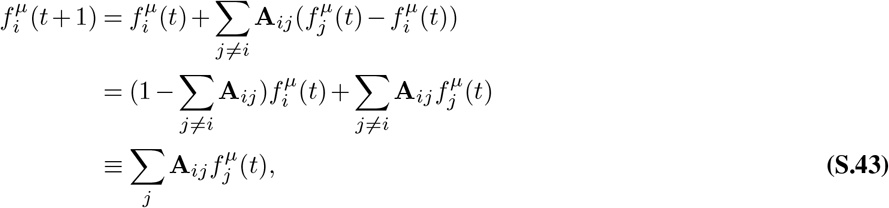

where **A**_*ii*_ ≡ 1 − Σ_*j i*_ **A**_*ij*_ (or equivalently, Σ_*j*_ **A**_*ij*_ = 1). We note that the diagonal elements **A**_*ii*_ in the time-discrete dynamics are determined from the normalization, and the transmission rate within a deme, *β*_*ii*_, does not enter the neutral dynamics.

In ref. (53), *β*_*ij*_ is modeled in terms of human mobility as

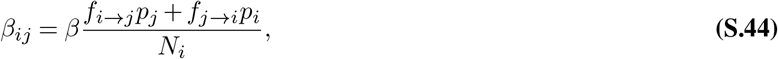

where *β* is the transmission rate when a susceptible individual has contact with an infected, *p*_*i*_ is the contact probability in deme *i*, and *f*_*i*→*j*_ is the mobility flux from *i* to *j*. The first term in the numerator corresponds to the case that a susceptible person in deme *i* visits deme *j* and gets infected by a residence of deme *j*, while the second term corresponds to the case that an infected person in deme *j* visits deme *i* and infects a resident of deme *i*. Note that, in ref. (53), *p*_*i*_ is assumed to be the same for all demes. Using Eq. (S.44), **A**_*ij*_ in Eq. (S.42) can be expressed in terms of the mobility flux as

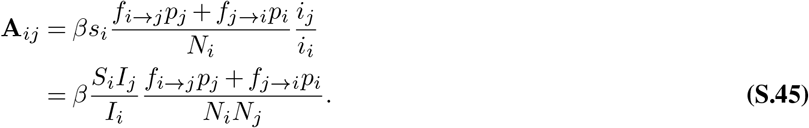

The expression in Eq. (S.45) implies:

1. The combination 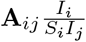 is symmetric in *i* and *j*.
2. The asymmetry in the couplings between *i* and is given by 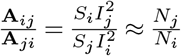, where the densities *s*_*i*_ and *i*_*i*_ are assumed to be approximately the same for all demes. This expression of the asymmetry means that a deme with a larger population size is expected to have a greater impact on the other.
3. **A**_*ij*_ in Eq. (S.45) satisfies the detailed balance condition. Specifically, the reproductive value(the left null vector of **A**_*ij*_) is given by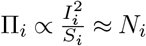.

##### S.5.2 Non-neutral variant

Consider the case of two non-neutral lineages, specifically the Alpha and Delta variants (*µ* = *α, δ*). We express 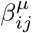 and *γ*^*µ*^ for these variants as follows: 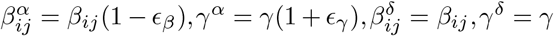 and denote the Delta’s frequency as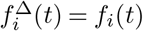.

Using these expressions, Eq. (S.40) for *µ* = *δ* becomes:

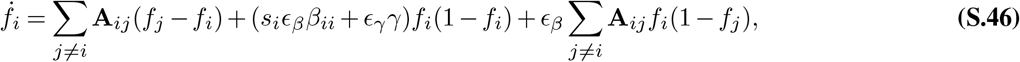

where 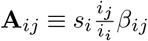 denotes the importation-rate matrix for the Delta variant. When the Delta variant is rare, *f*_*i*_ ≪ 1, we can drop the terms quadratic in frequencies:

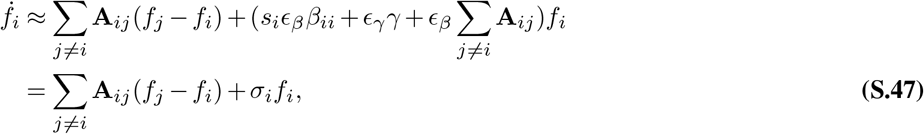

where *σ*_*i*_ ≡ *s*_*i*_*ϵ*_*β*_*β*_*ii*_ + *ϵ*_*γ*_*γ* + *ϵ*_*β*_ Σ_*i*≠*j*_ = **A**_*ij*_. In the main text, the subscript of *σ*_*i*_ is dropped, assuming that *s*_*i*_*ϵ*_*β*_*β*_*ii*_ and Σ_*j≠i*_ **A**_*ij*_(= 1 − **A**_*ii*_) are not significantly different across populations.

##### S.5.3 Other epidemiological scenarios

While we verified the linear dynamics *f*_*i*_(*t* + 1) = _*j*_ **A**_*ij*_*f*_*j*_(*t*) in Eq. S.43 assuming the standard transmission function *βSI/N*, it is worth noting that this result holds independently of specific transmission functions employed (see ref. (58) for a variety of transmission functions). For instance, if the transmission function takes the form 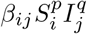 with some exponents *p, q >* 0, Eq. S.36 would be replaced by:

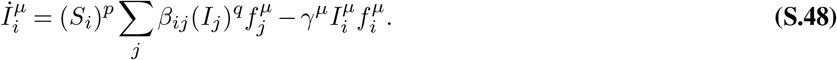

Note that the transmission term is linear in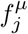, which guarantees that the sum over lineages yields the total transmission rate. More generally, Eq. S.36 can be generalized to:

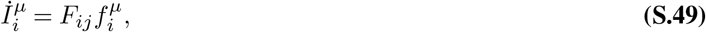

where *F*_*ij*_ is any lineage-independent function that may depend on the total numbers of susceptible and infected individuals, as well as any other lineage-independent quantities such as population densities and geographic distances. It can then be shown that the neutral dynamics of a lineage frequency is given by Eq. S.41 with:

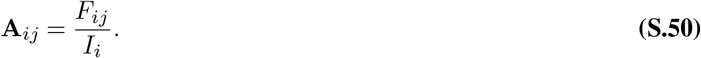

Furthermore, the linear dynamics *f*_*i*_(*t* + 1) = Σ_*j*_ **A**_*ij*_*f*_*j*_(*t*) can also be justified in another important class of epidemiological models, the SEIR model, which has been widely applied to SARS-CoV-2 (refs. (11, 59)). A multi-patch extension of the SEIR model is described by the following equations:

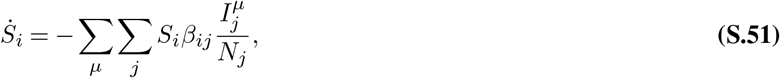

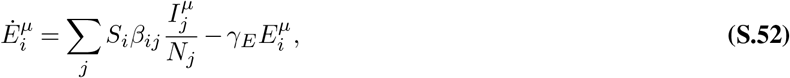

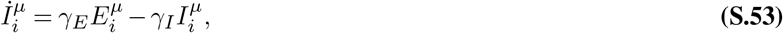

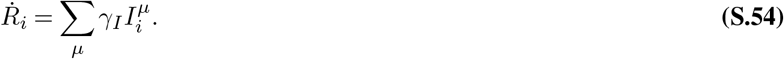

Here, 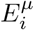 represents the number of asymptomatic individuals in deme *i* who have been infected by lineage *µ. γ*_*E*_ is the rate at which an exposed individual becomes infectious (the inverse of the average latent time), and *γ*_*I*_ is the rate at which an infectious individual recovers. The total population size, *N*_*i*_ = *S*_*i*_ + *I*_*i*_ + *E*_*i*_ + *R*_*i*_, is constant in each deme, where 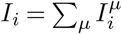 and 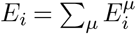, respectively, represent the total infected and exposed populations in deme *i* across all lineages.

We can show that the lineage frequencies, 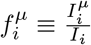 and 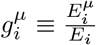, among exposed and infected individuals, obey the following equations:

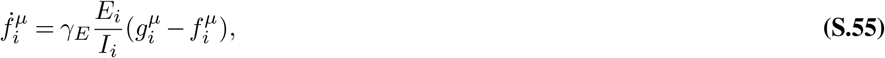

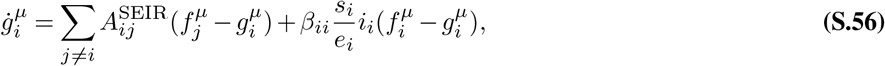

with

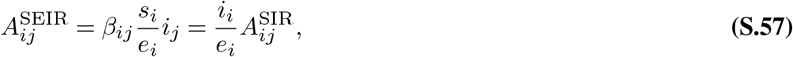

where 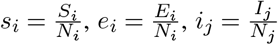 are the fractions of these epidemiological classes, and 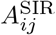 is the matrix in Eq. S.42. The matrix 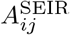 differs from 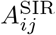 by the factor 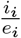. This factor is largely determined by the ratio 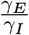 (see Eq. S.53) and takes similar values across demes.

By subtracting Eqs. S.55 and S.56, we obtain

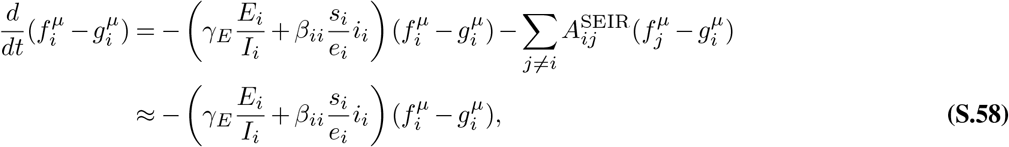

where we dropped the off-diagonal couplings, which are usually smaller than the within-deme transmission, 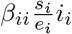. The coefficients 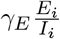 and 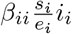 in Eq. S.58 can be roughly approximated as 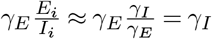 and 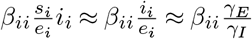. Evaluating these with realistic parameter values (for instance, *β*_*ii*_ = 0.4 day^−1^, *γ*_*E*_ = (3.0 day)^−1^, *γ*_*I*_ = (5.5 day)^−1^ (ref. (59)), we observe that the difference 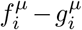 relaxes to 0 within 1 week. Thus, we may practically identify 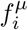 and 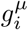. Consequently, from Eq. S.56, with this identification, we find that in the SEIR model, the dynamics of the lineage frequency 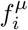 is described by

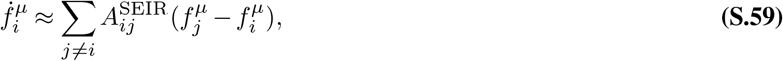

where, as argued above, 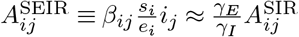.

#### S.6 The jump-size distribution calculated from the importation-rate matrix A

In Fig. 3C, we computed the probability distribution of per-individual jump distances, assuming that a jump from deme *i* to deme *j* occurs with a probability proportional to 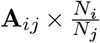. The rationale for this population-size-dependent rescaling is that the impact of a source location *j* on a target location *i* will be greater if the source population size *N*_*j*_ is larger and the target population size *N*_*i*_ is smaller. To offset these effects, we rescaled **A**_*ij*_ by the factor 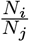, thereby obtaining the per-individual jump rate.

We can also justify the rescaling factor based on the SIR model. As described in SI Sec. S.5, when deriving Eq. S.44, two possibilities for an infection event are considered: (i) a susceptible person in deme *i* visits deme *j* and gets infected by a resident of deme *j*, and (ii) an infected person in deme *j* visits deme *i* and infects a resident of deme *i*. Assuming that the second contribution is dominant, we would obtain 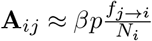 instead of Eq. S.45. Here, we assume *s*_*i*_ ≈ 1, *i*_*i*_ ≈ *i*_*j*_, and *p*_*i*_ = *p*. Consequently, the per-individual jump rate from location *j* to *i*, 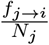, is proportional to 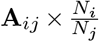, consistent with the above intuitive argument.

#### S.7 Distance between demes used for the hierarchical clustering of A_*ij*_ and multidimensional scaling analysis

For the deme-resolved coupling matrices **A** shown in Figs. 3 and 6, we ordered demes according to how similar their rows are. To this end, we performed hierarchical clustering using Ward’s method with the following metric *d*_*ij*_ between populations;

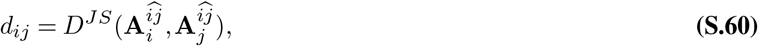

where *D*^*JS*^(***p, q***) is the square root of the Jensen-Shannon divergence between two probability distributions ***p*** and 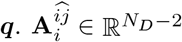 (resp. 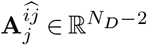) is the vector of the probability distribution obtained by removing the *i*-th and *j*-th elements from the *i*-th (resp. *j*-th) row of the importation-rate matrix. Explicitly, their *k*-th elements are given by

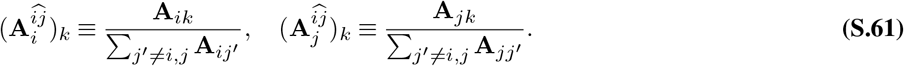

In the backward-in-time interpretation, the metric *d*_*ij*_ compares the probability distribution of the spatial locations of a lineage starting at population *i* and the one starting at population *j*, conditional on that they go to populations outside of *i* and *j*.

Figures S15A and B show the results of hierarchical clustering using Ward’s method, and the MDS analysis with the above metric for the matrix powers **A**_*n*_ (*n* = 1, 5, 10, 20), where **A** is the importation-rate matrix for the Delta wave in England (Fig. 3). Fig. S15C compares the Jensen-Shannon divergence to the physical distances between demes. We can see that the clustering of the 50 demes is relatively robust to the timescale *n* of interest. However, as *n* becomes large (compared to the timescale of relaxation, approximately *n* = 10 weeks), all rows of (**A**_*n*_)_*ij*_ approach the reproductive value***π*** (i.e., the left eigenvector of **A** corresponding to the eigenvalue 1), and the Jensen-Shannon divergence becomes less informative of physical distances. For example, for (**A**^20^)_*ij*_, demes in LDN, SE, and SW become highly clustered in the MDS plot (the bottom panel of Fig. S15B), indicating that the Jensen-Shannon divergence cannot effectively distinguish between physically close demes (the bottom panel of Fig. S15C).

**Fig. S15.**
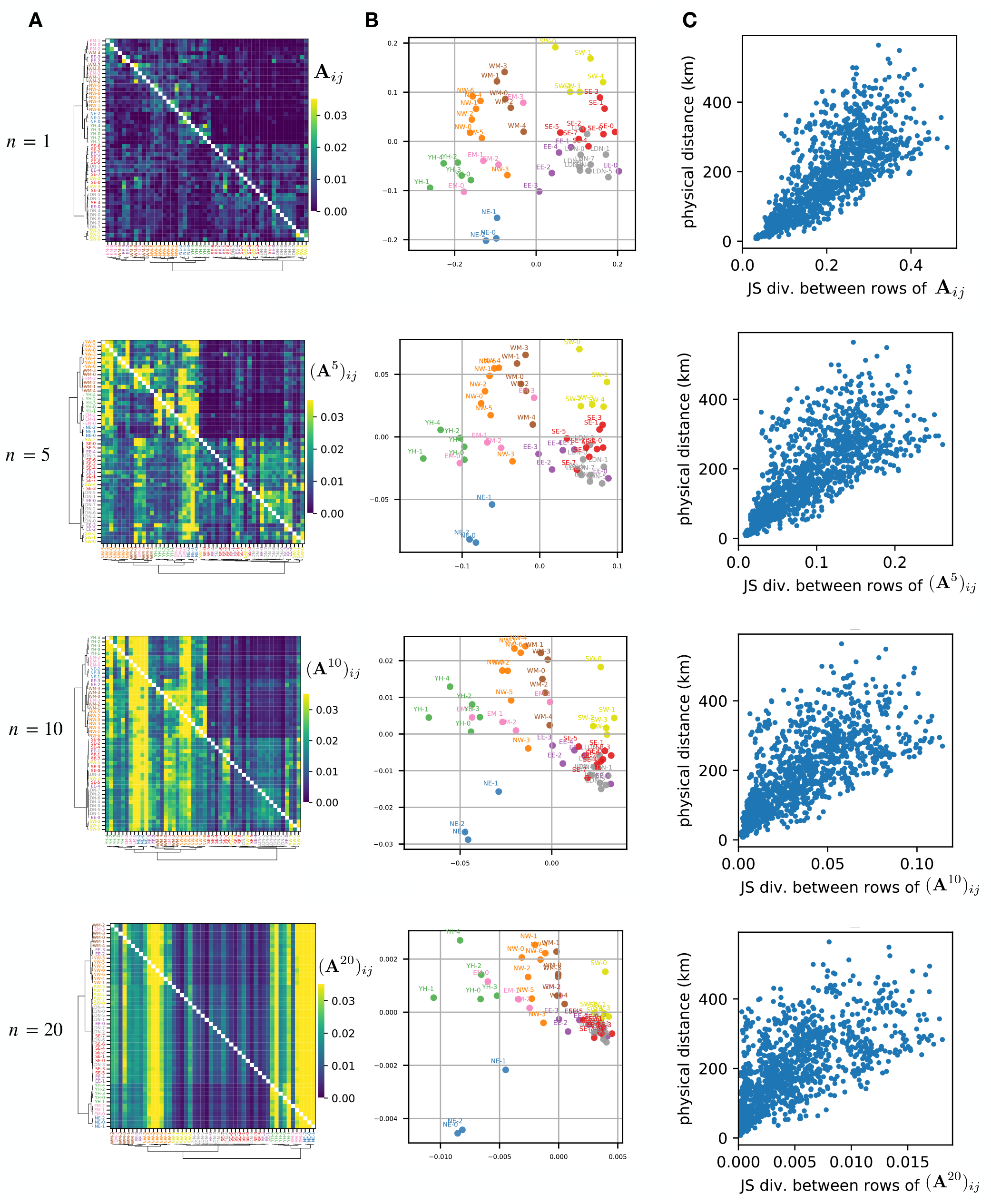
Clustering and distances based on the Jensen-Shannon divergence for the (*A*^*n*^) for *n* = 1, 5, 10, 20, where **A** is the 50 *×* 50 matrix for the Delta wave (weeks 78-91) in England. (**A**) Heatmaps of **A**^*n*^(*n* = 1, 5, 10, 20), where hierarchical clustering is performed using the Ward’s method with the Jensen-Shannon divergence between the rows of **A**^*n*^. (**B**) Multidimensional scaling plots. (**C**) Comparison between the Jensen-Shannon divergence and physical distance, with each dot representing these two distances for a specific pair of demes *i* and *j*.

#### S.8 Prediction of the spreading dynamics of the Delta variant shown in Fig. 8

*Non-neutral transmission dynamics:* We modeled the frequency dynamics of the Delta variant in England using the ordinary differential equation in the form of Eq. S.46:

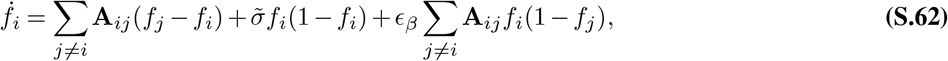

where **A**_*ij*_ represents the 50 × 50 importation-rate matrix presented in Fig. 3, and 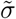 and *ϵ*_*β*_ are fit parameters.

*Determination of the parameter values:* To fix 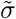 and *ϵ*, we simulated the above ODE and identified the optimal parameter values that minimize the discrepancies between our model’s predictions and the observed frequencies in a logit scale:

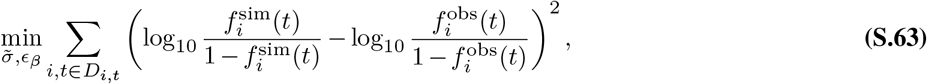

where 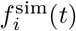 and 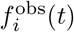 denote the simulated and observed frequencies, respectively, of the Delta variant. The data points 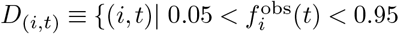 and (the number of sequences reported from location *i* in week *t*) *>* 10 are used for the fitting.

We examined the discrepancy between theoretical predictions and observed data, Eq. S.63, across a range of parameter values (Fig. S16). The discrepancy has a weak dependence on *ϵ*_*β*_, indicating that it cannot be reliably estimated solely from our fitting process. Therefore, we referred to the relative infectivity reported in ref. (35), which found that the Delta variant is 43–68% more transmissible than the Alpha variant, corresponding to *ϵ*_*β*_ values between 0.30 and 0.40 (see Eq. S.46 and the definition of *ϵ*_*β*_). Based on these, we constrained our optimization search to this realistic range, resulting in the optimal values, 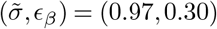. These parameter values are used in the simulations presented in the main text.

**Fig. S16.**
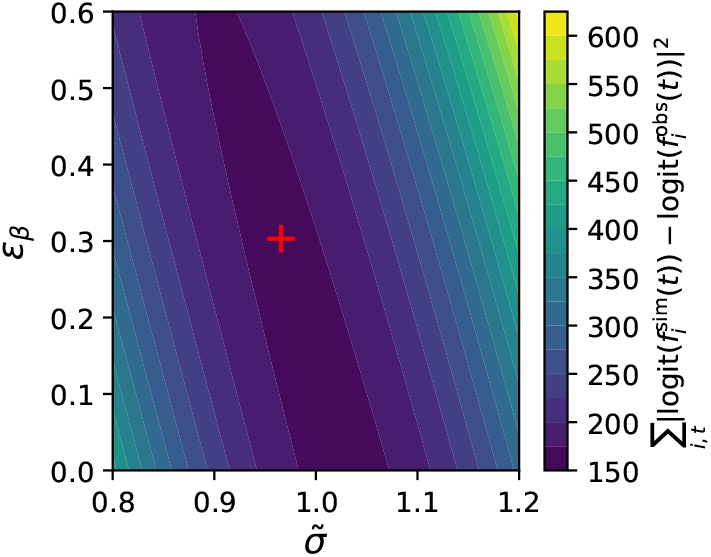
Heatmap showing the discrepancy between theoretical predictions and observed data, defined in Eq. S.63, as a function of 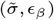. The parameter values, indicated by the red cross, are used for the results presented in the main text.

*Determination of the mid timepoint t*_1*/*2_: To compare the observed and simulated frequency trajectories, we calculated the mid timepoint *t*_1*/*2_ for each trajectory *f*_*i*_(*t*) by fitting it to a logistic curve, 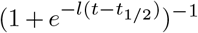, where *l* and *t*_1*/*2_ are fit parameters (see Fig. S17 for the actual trajectories and the fitting results).

**Fig. S17.**
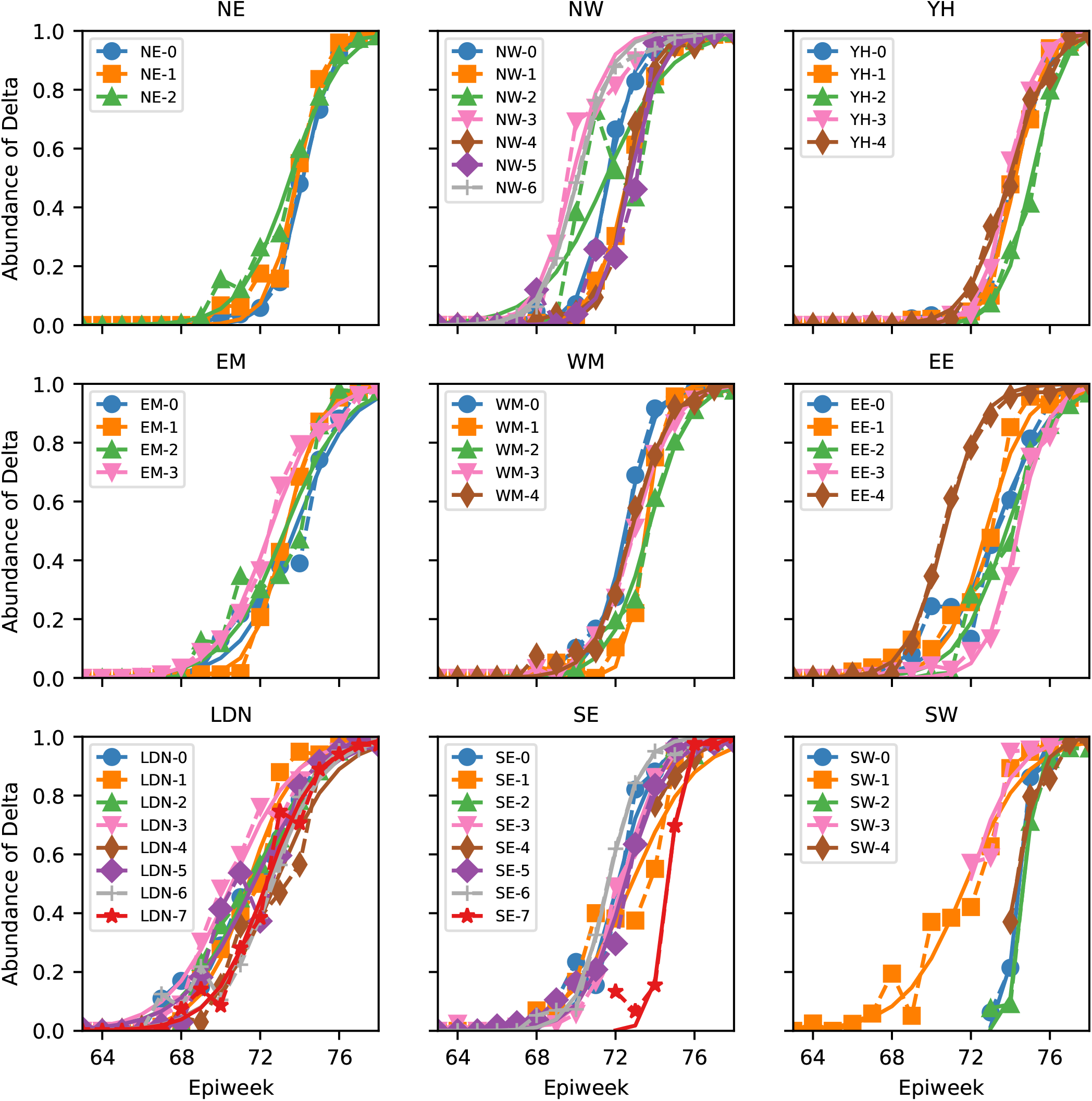
Procedure of determining *t*_1*/*2_ for the actual frequencies of the Delta variant in England. In each panel, the dashed lines represent the observed frequencies 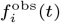 in demes within a region, while the solid lines represent fitted logistic curves 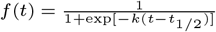, where *k* and *t*_1*/*2_ are fit parameters. For each deme, the time window with more than 10 sequences is used for the fitting. The inflection points *t*_1*/*2_ of these fitted logistic curves are used in Fig. 8 of the main text.

**Fig. S18.**
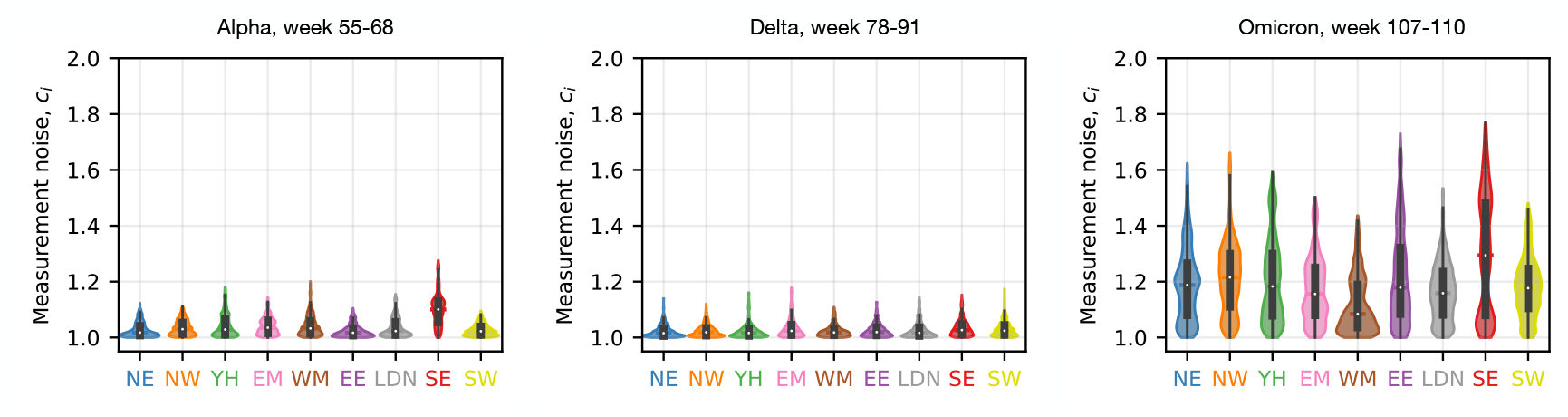
The posterior distributions of the measurement noise parameters *c*_*i*_ for the Alpha, Delta, and Omicron variants in England, which are inferred by applying the HMM-MCMC method to the region-level mutation data.

**Fig. S19.**
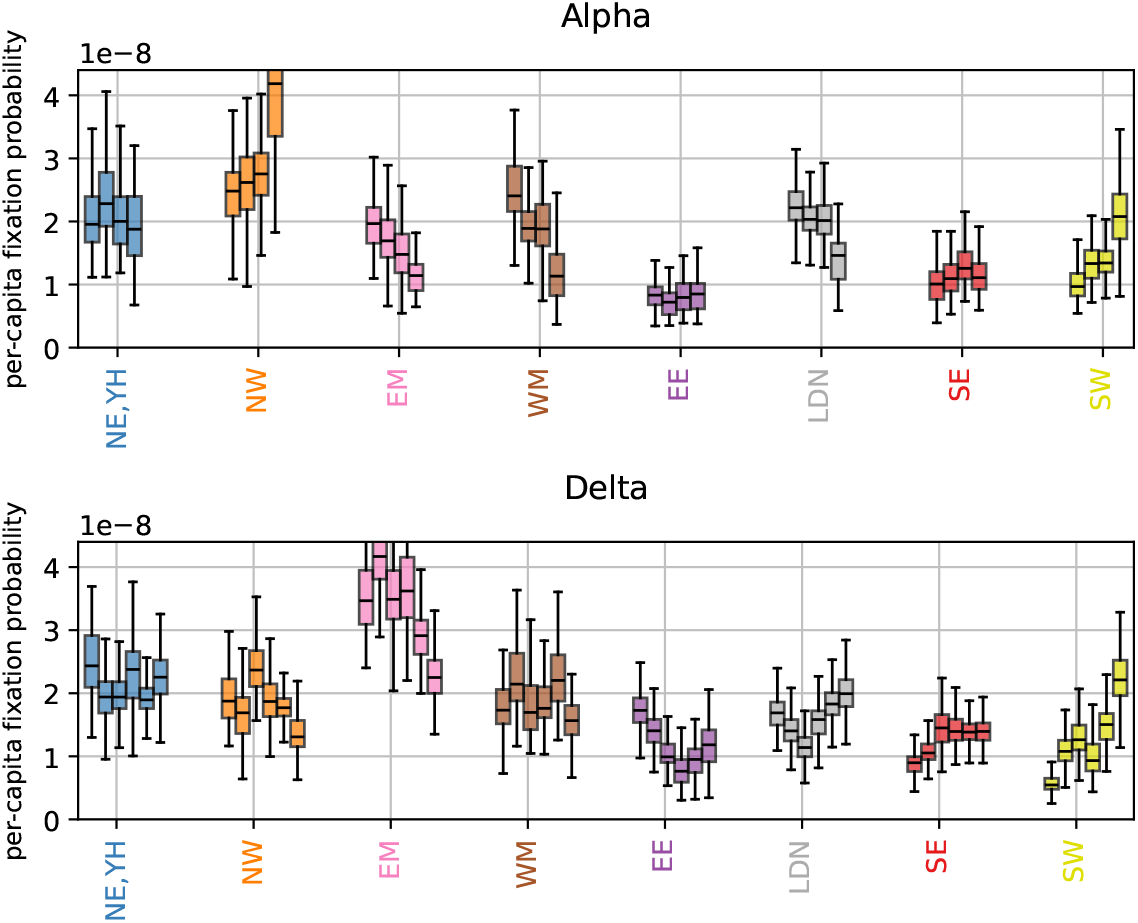
Supplementary figure to Fig. 7C. The reproductive value in England is inferred at the regional level, with NE and YH being treated as a single region. For the Alpha wave, the time windows used for the four box plots are epiweek 54-67, 55-68, 56-69, and 57-70, respectively, from left to right. For the Delta wave, the time windows used for the six box plots are epiweek 78-91, 79-92, 80-93, 81-94, 82-95, and 83-96, respectively, from left to right. For each variant, we aggregated the results for all the time windows, and the aggregated results are shown in Fig. 7C.

**Fig. S20.**
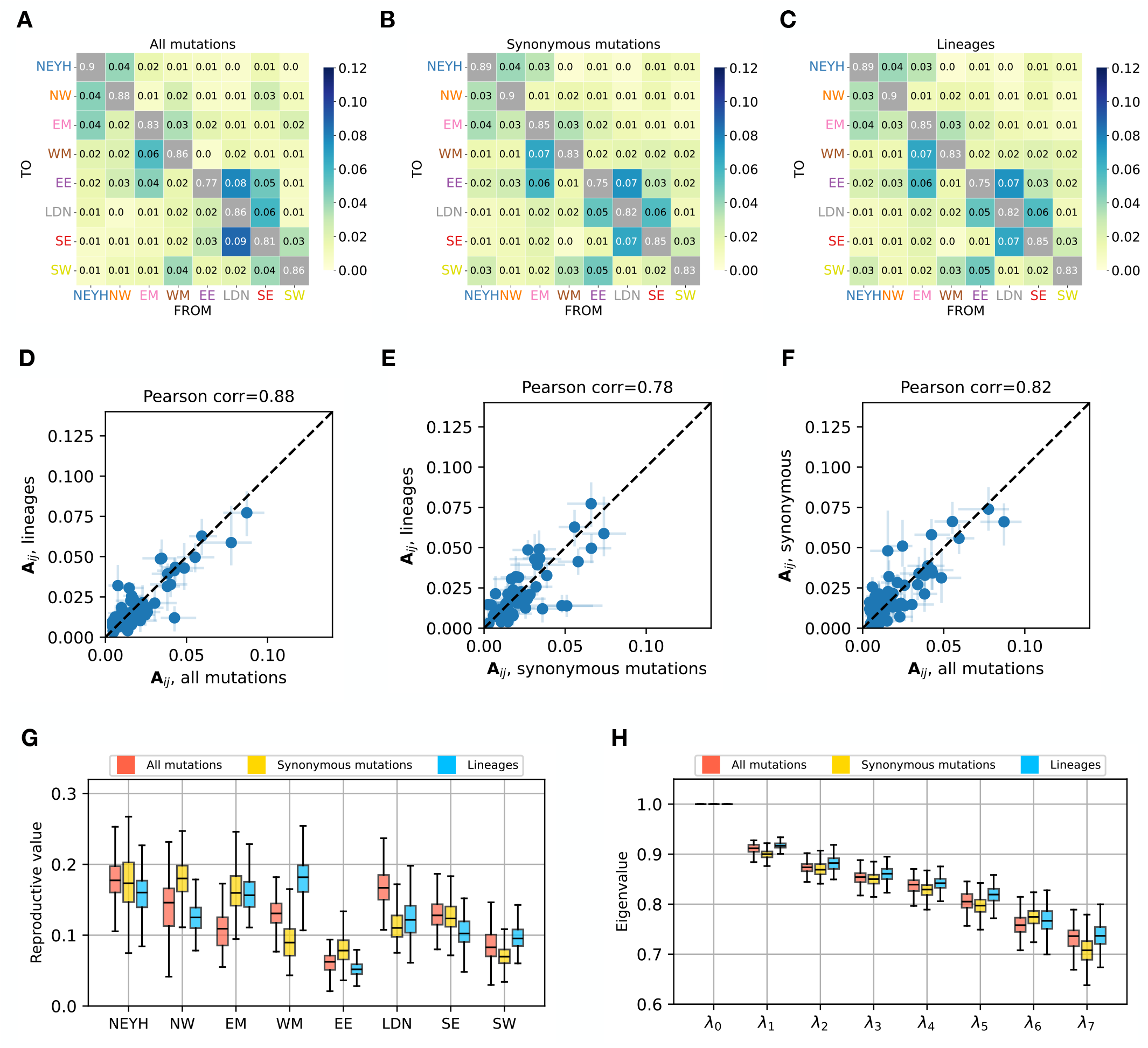
The heatmaps (**A-C**) display region-level inferred matrices for the Delta wave (epiweeks 78-91) in England, where YH and NE regions are combined, for different data types: (**A**) mutation data, incorporating both synonymous and nonsynonymous mutations; (**B**) synonymous mutation data; (**C**) lineage data. The mean values of parameters, computed using MCMC, are presented in these heatmaps. (**D-F**) provide element-wise comparisons of these matrices, with error bars indicating standard deviations. (**G**) Comparison of the reproductive values *π*_*i*_. (**H**) Comparison of the eigenvalues *λ*_*i*_ of the importation-rate matrix.

**Fig. S21.**
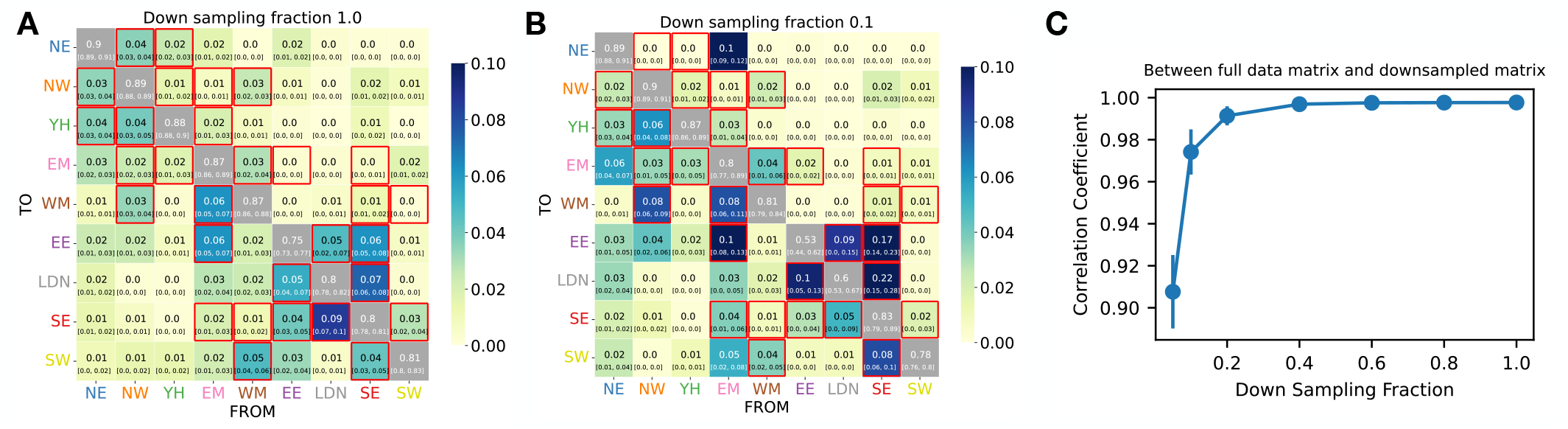
(**A**) The importation-rate matrix for the Delta wave in England, inferred from the full dataset. The matrix is inferred using the EM algorithm, and the error indicates the lower and upper quartiles obtained from the bootstrapping method. (**B**) The importation-rate matrix inferred from 10% of the data. (**C**) The Pearson correlation coefficient between the matrix elements of the full dataset and those of the downsampled matrix is computed for downsampling fractions of 5%, 10%, 20%, 40%, 60%, 80%, and 100% (i.e., the full dataset). The error bar indicates the standard deviation in the Pearson correlation coefficient obtained from the bootstrapping method. The matrix elements inferred from the downsampled data begin to deviate from those inferred from the full dataset when the fraction of downsampling is less than 10%.

**Fig. S22.**
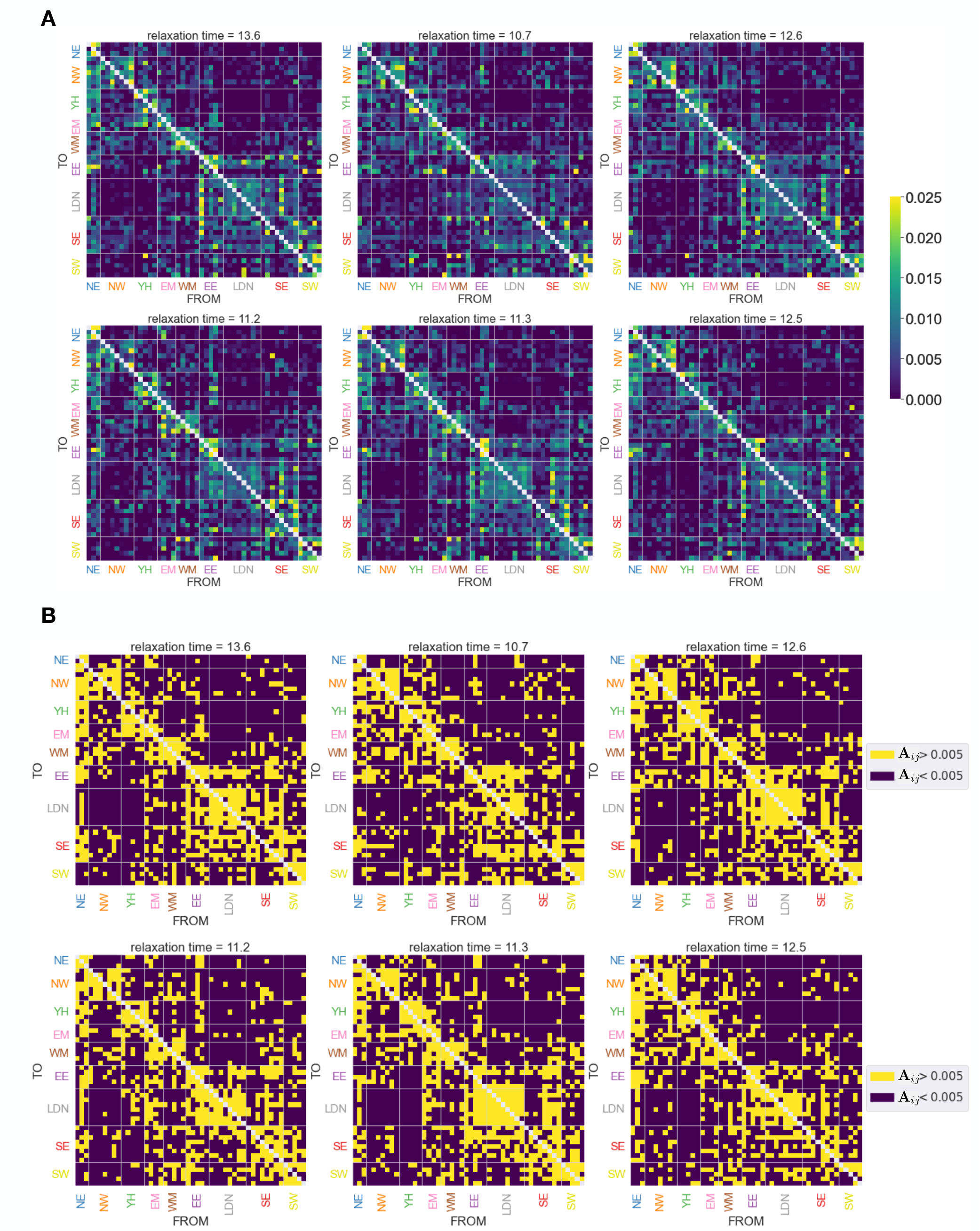
(**A**) Six examples of transmission-rate matrices inferred for the Delta wave in England, obtained through the bootstrapping method. For each matrix, the relaxation time computed from the matrix is written on the heatmap. (**B**) Each of the six matrices from Fig. **A** is binarized with a threshold of **A**_*ij*_ = 0.005.

**Fig. S23.**
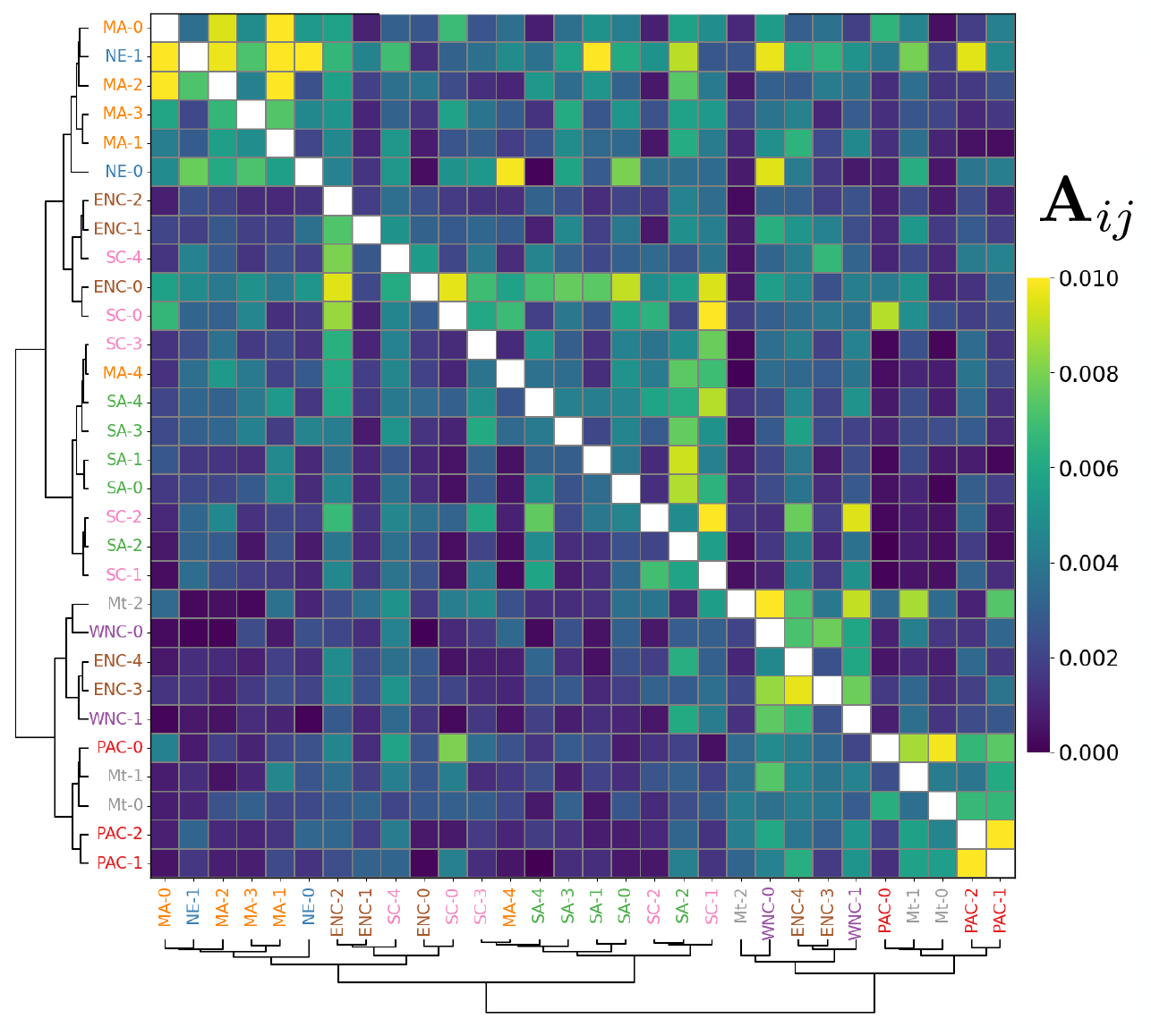
Heat map of the importation-rate matrix **A**_*ij*_ for 30 demes of the USA during the Delta wave (epiweek 82-96). The main infection pathways are illustrated in Fig. 6 of the main text.

**Fig. S24.**
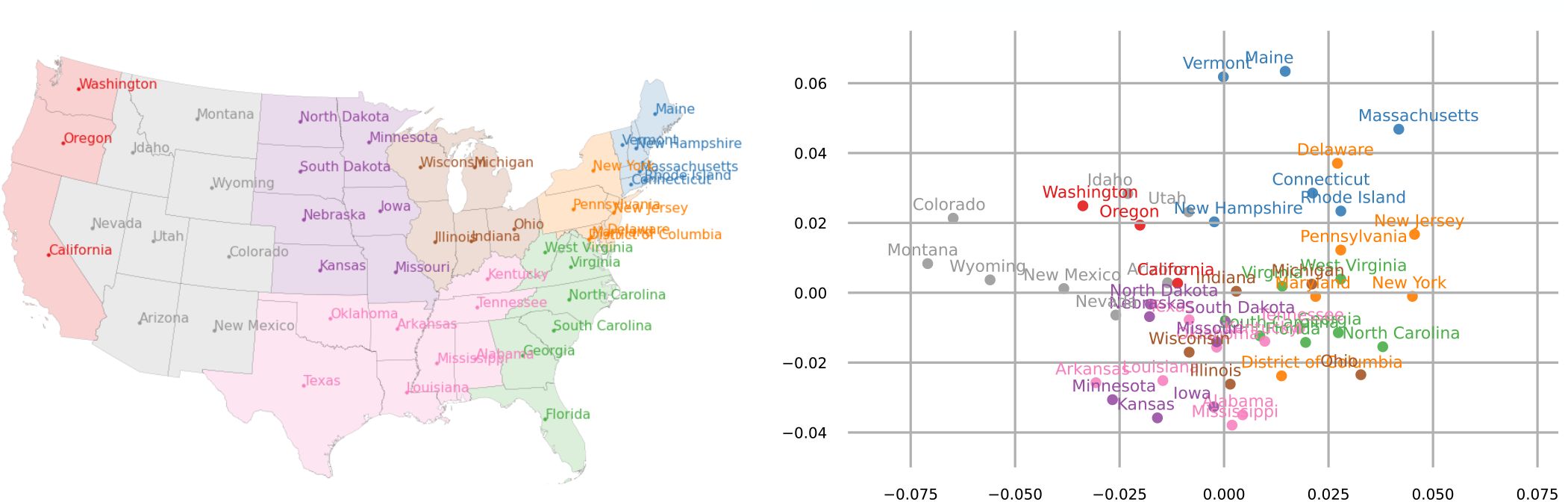
Comparison between the actual geography and the multidimensional scaling plot for the state-level importation-rate matrix of the Delta wave in the US.

**Fig. S25.**
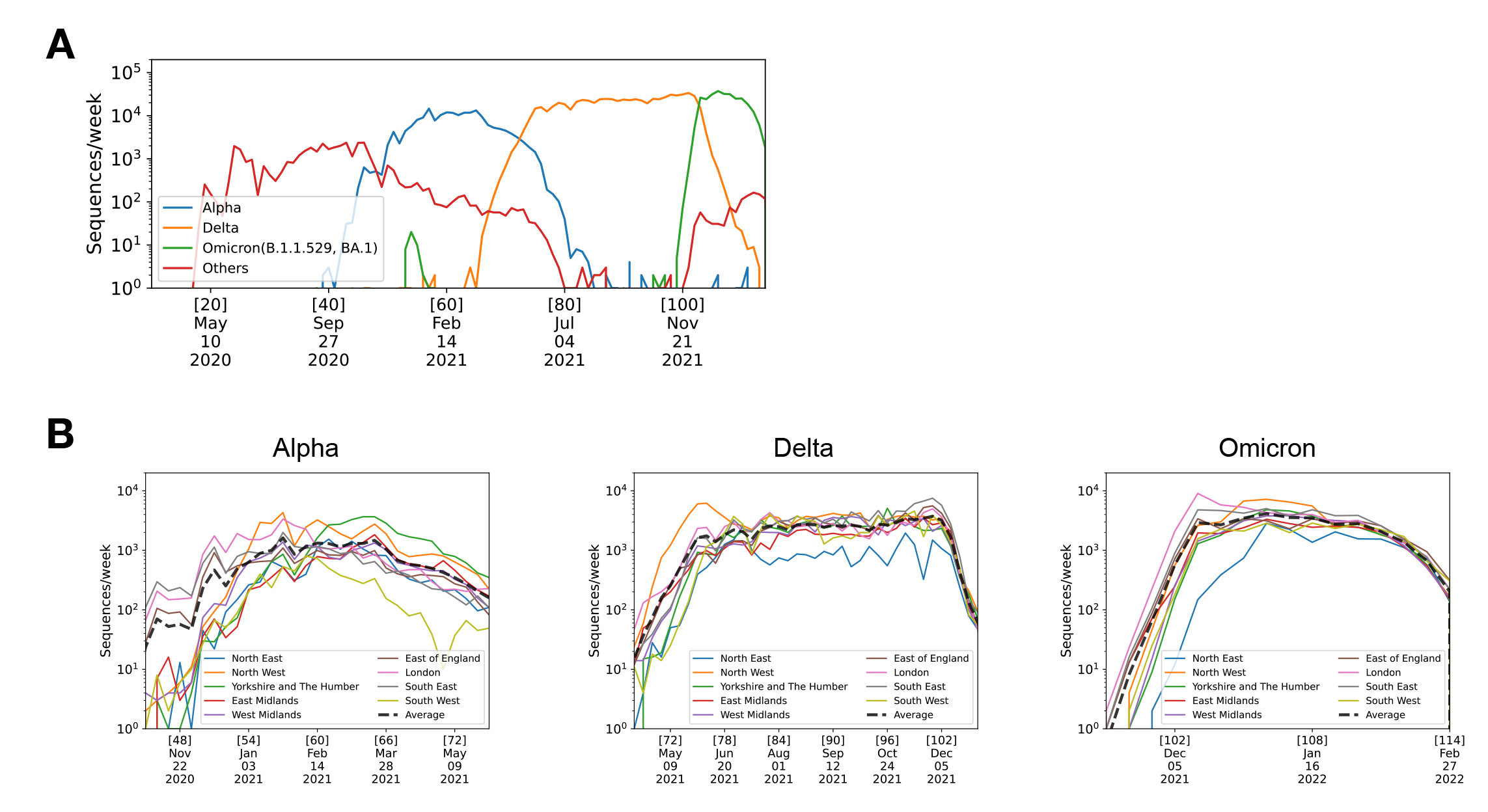
(**A**) The number of sequences per week for each variant in England. The numbers [*·*] shown above dates in the horizontal axis indicate epiweeks since Dec/29/2021. (**B**) The weekly number of sequences in each region of England for Alpha, Delta, and Omicron. The numbers [*·*] shown above dates in the horizontal axis indicate epiweeks since Dec/29/2021.

**Fig. S26.**
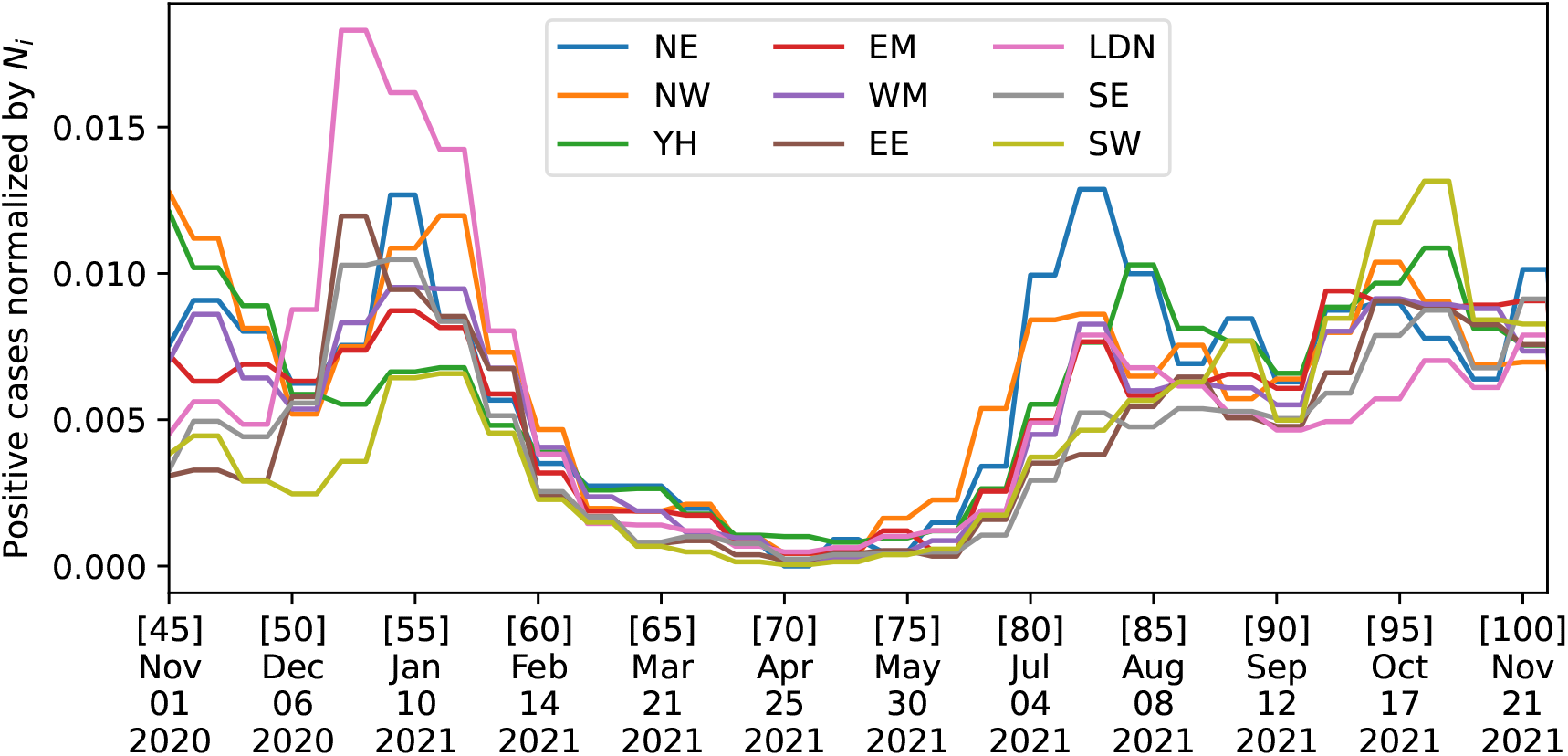
The number of weekly infected individuals (estimated by surveillance testing (37)) divided by the population size across over time in the regions of England.

#### S.9 Construction of demes

##### S.9.1 Construction of demes in England

For the deme-resolved analysis of England, presented in Fig. 3, we constructed 50 demes from the upper tier local authorities (UTLAs). This involved clustering within each of the 9 regions of England in the following manner:

1. First, we select the 150 largest UTLAs based on the number *s*_*u*_ of metadata sequences reported from each UTLA *u*.
2. In each of the 9 regions, we create a connected graph using Delaunay Triangulation, using the geographic locations of these UTLAs. Edges that cross over the ocean are removed (Fig. S27A).

To create 50 demes with similar sizes, we determine the number *n*_*i*_ of demes to be assigned to each of the 9 regions using a greedy algorithm:

1. As the initial state, we set *n*_*i*_ = 2 for all regions *i*.
2. Assuming that, at an intermediate step of the algorithm, the 9 regions respectively have *n*_*i*_ demes. We define the weight of each region as 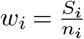, where 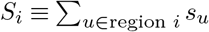 is the total number of sequences in region *i*. We then increase *n*_*i*_ by 1 for the region with the smallest weight.
3. This process is repeated until the sum of *n*_*i*_ across all regions reaches the target number of demes, 50. The resulting allocation of demes was *n*_*i*_ = 3, 7, 5, 4, 5, 5, 8, 8, 5 for *i* = NE, NW, YH, EM, WM, EE, LDN, SE, SW, respectively.

Finally, for each region *i*, we organize *n* demes (where the subscript *i* of *n*_*i*_ is omitted for notational simplicity) by grouping the UTLAs in the region into non-overlapping sets *U*_1_, …, *U*_*n*_:

1. Suppose that region *i* includes *M* UTLAs in the graph, labeled by *u*_*α*_ (*α* = 1,…, *M*). By arranging the UTLAs, we may assume that *s*_1_ ≥ *s*_2_ ≥ … ≥ *s*_*M*_, where *s*_*α*_ is the number of sequences reported from UTLA *u*_*α*_.
2. We initialize the sets *U*_*k*_ with the first *n* UTLAs, namely, the largest *n* UTLAs, as *U*_*k*_ = {*u*_*k*_} (*k* = 1,…, *n*).
3. For the remaining UTLAs *u*_*n*+1_, …, *u*_*M*_, we add each UTLA one by one to its nearest set on the graph constructed from the Delaunay Triangulation. Here, the distance between UTLA *u* and set *U*_*k*_ is defined as 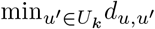, where 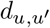 is the graph distance; for example, the distance is 1 for adjacent UTLAs.
4. If the nearest set for UTLA *u* is not unique, we add it to the set with the fewest sequences, to mitigate the imbalance in deme sizes.

The resulting demes are shown in Fig. S27B and summarized in Table 1.

**Fig. S27.**
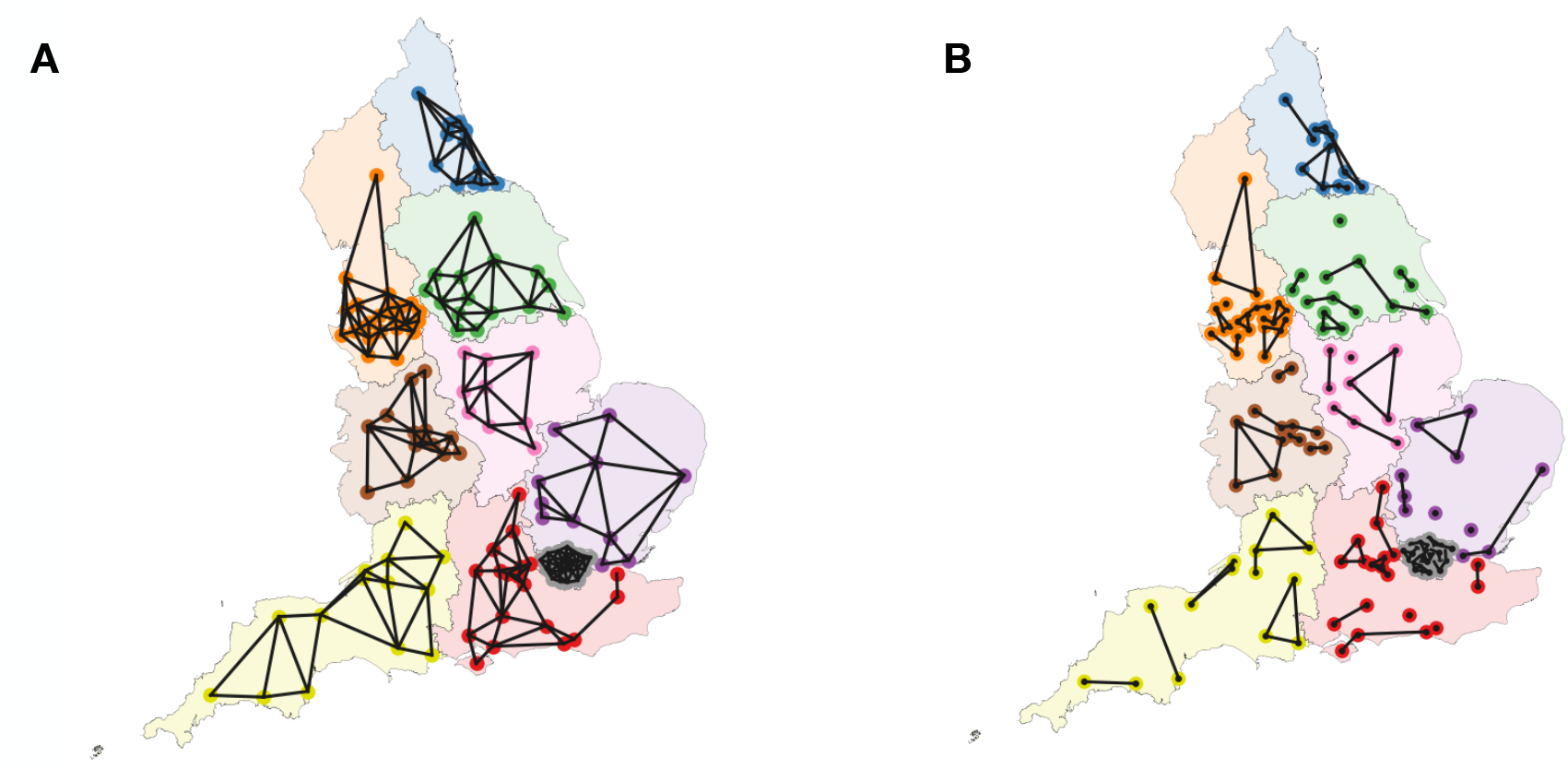
(**A**) The nodes represent the top 150 UTLAs with the highest number of sequences in the metadata. Within each region, a graph is constructed using Delaunay triangulation. (**B**) The UTLAs in each region are grouped into *n*_*i*_ demes, using the information of the number of sequences. The 50 demes constructed in this manner are shown as connected components.

**Table 1.**
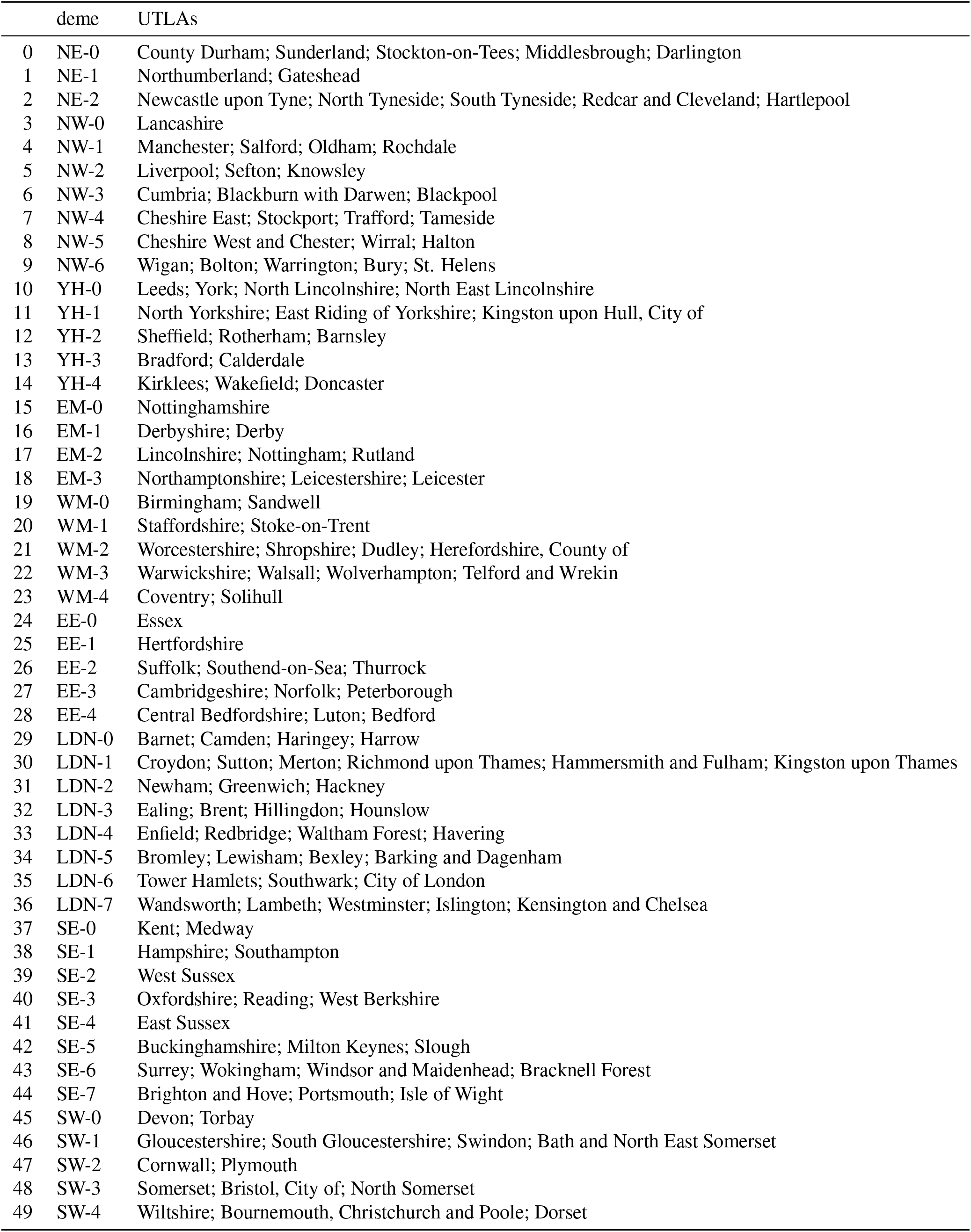
List of 50 demes in England.

|  | deme | UTLAs |
| --- | --- | --- |
| 0 | NE-0 | County Durham; Sunderland; Stockton-on-Tees; Middlesbrough; Darlington |
| 1 | NE-1 | Northumberland; Gateshead |
| 2 | NE-2 | Newcastle upon Tyne; North Tyneside; South Tyneside; Redcar and Cleveland; Hartlepool |
| 3 | NW-0 | Lancashire |
| 4 | NW-1 | Manchester; Salford; Oldham; Rochdale |
| 5 | NW-2 | Liverpool; Sefton; Knowsley |
| 6 | NW-3 | Cumbria; Blackburn with Darwen; Blackpool |
| 7 | NW-4 | Cheshire East; Stockport; Trafford; Tameside |
| 8 | NW-5 | Cheshire West and Chester; Wirral; Halton |
| 9 | NW-6 | Wigan; Bolton; Warrington; Bury; St. Helens |
| 10 | YH-0 | Leeds; York; North Lincolnshire; North East Lincolnshire |
| 11 | YH-1 | North Yorkshire; East Riding of Yorkshire; Kingston upon Hull, City of |
| 12 | YH-2 | Sheffield; Rotherham; Barnsley |
| 13 | YH-3 | Bradford; Calderdale |
| 14 | YH-4 | Kirklees; Wakefield; Doncaster |
| 15 | EM-0 | Nottinghamshire |
| 16 | EM-1 | Derbyshire; Derby |
| 17 | EM-2 | Lincolnshire; Nottingham; Rutland |
| 18 | EM-3 | Northamptonshire; Leicestershire; Leicester |
| 19 | WM-0 | Birmingham; Sandwell |
| 20 | WM-1 | Staffordshire; Stoke-on-Trent |
| 21 | WM-2 | Worcestershire; Shropshire; Dudley; Herefordshire, County of |
| 22 | WM-3 | Warwickshire; Walsall; Wolverhampton; Telford and Wrekin |
| 23 | WM-4 | Coventry; Solihull |
| 24 | EE-0 | Essex |
| 25 | EE-1 | Hertfordshire |
| 26 | EE-2 | Suffolk; Southend-on-Sea; Thurrock |
| 27 | EE-3 | Cambridgeshire; Norfolk; Peterborough |
| 28 | EE-4 | Central Bedfordshire; Luton; Bedford |
| 29 | LDN-0 | Barnet; Camden; Haringey; Harrow |
| 30 | LDN-1 | Croydon; Sutton; Merton; Richmond upon Thames; Hammersmith and Fulham; Kingston upon Thames |
| 31 | LDN-2 | Newham; Greenwich; Hackney |
| 32 | LDN-3 | Ealing; Brent; Hillingdon; Hounslow |
| 33 | LDN-4 | Enfield; Redbridge; Waltham Forest; Havering |
| 34 | LDN-5 | Bromley; Lewisham; Bexley; Barking and Dagenham |
| 35 | LDN-6 | Tower Hamlets; Southwark; City of London |
| 36 | LDN-7 | Wandsworth; Lambeth; Westminster; Islington; Kensington and Chelsea |
| 37 | SE-0 | Kent; Medway |
| 38 | SE-1 | Hampshire; Southampton |
| 39 | SE-2 | West Sussex |
| 40 | SE-3 | Oxfordshire; Reading; West Berkshire |
| 41 | SE-4 | East Sussex |
| 42 | SE-5 | Buckinghamshire; Milton Keynes; Slough |
| 43 | SE-6 | Surrey; Wokingham; Windsor and Maidenhead; Bracknell Forest |
| 44 | SE-7 | Brighton and Hove; Portsmouth; Isle of Wight |
| 45 | SW-0 | Devon; Torbay |
| 46 | SW-1 | Gloucestershire; South Gloucestershire; Swindon; Bath and North East Somerset |
| 47 | SW-2 | Cornwall; Plymouth |
| 48 | SW-3 | Somerset; Bristol, City of; North Somerset |
| 49 | SW-4 | Wiltshire; Bournemouth, Christchurch and Poole; Dorset |

##### S.9.2 Construction of demes in the US

In the US analysis, shown in Fig. 6, we constructed 30 demes using an approach similar to that used in the England analysis: Since the USA has no direct counterpart to the regions of England, we defined 8 regions in the USA for the purpose of the analysis: NE, MA, SA, SC, ENC, WNC, Mt, PAC, as colored in Fig. 6B. We then performed the same clustering as in the England analysis, making the following replacements: 150 UTLAs → 49 states (excluding Hawaii and Alaska) and 50 demes → 30 demes. In addition, instead of using Delaunay Triangulation, we constructed the graph of the US states based on whether they are geographically neighboring. The resulting demes are summarized in Table 2.

**Table 2.** List of 30 demes in the US.

|  | deme | States |
| --- | --- | --- |
| 0 | NE-0 | Massachusetts,Maine,New Hampshire,Vermont |
| 1 | NE-1 | Connecticut,Rhode Island |
| 2 | MA-0 | New York |
| 3 | MA-1 | Pennsylvania |
| 4 | MA-2 | New Jersey |
| 5 | MA-3 | Maryland,District of Columbia |
| 6 | MA-4 | Delaware |
| 7 | SA-0 | Florida |
| 8 | SA-1 | Georgia |
| 9 | SA-2 | North Carolina |
| 10 | SA-3 | Virginia,West Virginia |
| 11 | SA-4 | South Carolina |
| 12 | SC-0 | Texas,Oklahoma |
| 13 | SC-1 | Tennessee,Arkansas |
| 14 | SC-2 | Alabama |
| 15 | SC-3 | Louisiana,Mississippi |
| 16 | SC-4 | Kentucky |
| 17 | ENC-0 | Illinois |
| 18 | ENC-1 | Ohio |
| 19 | ENC-2 | Michigan |
| 20 | ENC-3 | Indiana |
| 21 | ENC-4 | Wisconsin |
| 22 | WNC-0 | Missouri,Kansas |
| 23 | WNC-1 | Minnesota,Iowa,Nebraska,South Dakota,North Dakota |
| 24 | Mt-0 | Arizona |
| 25 | Mt-1 | Colorado,New Mexico,Wyoming |
| 26 | Mt-2 | Utah,Nevada,Idaho,Montana |
| 27 | PAC-0 | California |
| 28 | PAC-1 | Washington |
| 29 | PAC-2 | Oregon |

#### S.10 Calendar Dates and Weeks Since December 29, 2019

| Epiweek | Date | Epiweek | Date | Epiweek | Date |
| --- | --- | --- | --- | --- | --- |
| 1 | Dec 29, 2019 | 38 | Sep 13, 2020 | 75 | May 30, 2021 |
| 2 | Jan 05, 2020 | 39 | Sep 20, 2020 | 76 | Jun 06, 2021 |
| 3 | Jan 12, 2020 | 40 | Sep 27, 2020 | 77 | Jun 13, 2021 |
| 4 | Jan 19, 2020 | 41 | Oct 04, 2020 | 78 | Jun 20, 2021 |
| 5 | Jan 26, 2020 | 42 | Oct 11, 2020 | 79 | Jun 27, 2021 |
| 6 | Feb 02, 2020 | 43 | Oct 18, 2020 | 80 | Jul 04, 2021 |
| 7 | Feb 09, 2020 | 44 | Oct 25, 2020 | 81 | Jul 11, 2021 |
| 8 | Feb 16, 2020 | 45 | Nov 01, 2020 | 82 | Jul 18, 2021 |
| 9 | Feb 23, 2020 | 46 | Nov 08, 2020 | 83 | Jul 25, 2021 |
| 10 | Mar 01, 2020 | 47 | Nov 15, 2020 | 84 | Aug 01, 2021 |
| 11 | Mar 08, 2020 | 48 | Nov 22, 2020 | 85 | Aug 08, 2021 |
| 12 | Mar 15, 2020 | 49 | Nov 29, 2020 | 86 | Aug 15, 2021 |
| 13 | Mar 22, 2020 | 50 | Dec 06, 2020 | 87 | Aug 22, 2021 |
| 14 | Mar 29, 2020 | 51 | Dec 13, 2020 | 88 | Aug 29, 2021 |
| 15 | Apr 05, 2020 | 52 | Dec 20, 2020 | 89 | Sep 05, 2021 |
| 16 | Apr 12, 2020 | 53 | Dec 27, 2020 | 90 | Sep 12, 2021 |
| 17 | Apr 19, 2020 | 54 | Jan 03, 2021 | 91 | Sep 19, 2021 |
| 18 | Apr 26, 2020 | 55 | Jan 10, 2021 | 92 | Sep 26, 2021 |
| 19 | May 03, 2020 | 56 | Jan 17, 2021 | 93 | Oct 03, 2021 |
| 20 | May 10, 2020 | 57 | Jan 24, 2021 | 94 | Oct 10, 2021 |
| 21 | May 17, 2020 | 58 | Jan 31, 2021 | 95 | Oct 17, 2021 |
| 22 | May 24, 2020 | 59 | Feb 07, 2021 | 96 | Oct 24, 2021 |
| 23 | May 31, 2020 | 60 | Feb 14, 2021 | 97 | Oct 31, 2021 |
| 24 | Jun 07, 2020 | 61 | Feb 21, 2021 | 98 | Nov 07, 2021 |
| 25 | Jun 14, 2020 | 62 | Feb 28, 2021 | 99 | Nov 14, 2021 |
| 26 | Jun 21, 2020 | 63 | Mar 07, 2021 | 100 | Nov 21, 2021 |
| 27 | Jun 28, 2020 | 64 | Mar 14, 2021 | 101 | Nov 28, 2021 |
| 28 | Jul 05, 2020 | 65 | Mar 21, 2021 | 102 | Dec 05, 2021 |
| 29 | Jul 12, 2020 | 66 | Mar 28, 2021 | 103 | Dec 12, 2021 |
| 30 | Jul 19, 2020 | 67 | Apr 04, 2021 | 104 | Dec 19, 2021 |
| 31 | Jul 26, 2020 | 68 | Apr 11, 2021 | 105 | Dec 26, 2021 |
| 32 | Aug 02, 2020 | 69 | Apr 18, 2021 | 106 | Jan 02, 2022 |
| 33 | Aug 09, 2020 | 70 | Apr 25, 2021 | 107 | Jan 09, 2022 |
| 34 | Aug 16, 2020 | 71 | May 02, 2021 | 108 | Jan 16, 2022 |
| 35 | Aug 23, 2020 | 72 | May 09, 2021 | 109 | Jan 23, 2022 |
| 36 | Aug 30, 2020 | 73 | May 16, 2021 | 110 | Jan 30, 2022 |
| 37 | Sep 06, 2020 | 74 | May 23, 2021 | 111 | Feb 06, 2022 |

## Footnotes

1 There is an erratum in the corresponding equation in ref. (54).

## Notes

### Competing Interest Statement

The authors have declared no competing interest.

### Funding Statement

Humboldt Professorship of the Alexander von Humboldt Foundation (to OH). JSPS KAKENHI (Grant Numbers JP22K03453 and JP22K06347) and the RIKEN iTHEMS Program (to TO). QY acknowledges support from the National Science Foundation Graduate Research Fellowship under Grant No. DGE 1106400. GI acknowledges support of a Humboldt Research Fellowship by the Humboldt Foundation. This research used resources of the National Energy Research Scientific Computing Center (NERSC), a U.S. Department of Energy Office of Science User Facility located at Lawrence Berkeley National Laboratory, operated under Contract No. DE- AC02-05CH11231 using NERSC BER-ERCAP0019907.

### Author Declarations

The study used (or will use) ONLY openly available human data that are accessibly through the GISAID platform and the COG-UK consortium.

